# Exome sequencing of 20,979 individuals with epilepsy reveals shared and distinct ultra-rare genetic risk across disorder subtypes

**DOI:** 10.1101/2023.02.22.23286310

**Authors:** Epi25 Collaborative, Siwei Chen, Benjamin M Neale, Samuel F Berkovic

## Abstract

Identifying genetic risk factors for highly heterogeneous disorders like epilepsy remains challenging. Here, we present the largest whole-exome sequencing study of epilepsy to date, with >54,000 human exomes, comprising 20,979 deeply phenotyped patients from multiple genetic ancestry groups with diverse epilepsy subtypes and 33,444 controls, to investigate rare variants that confer disease risk. These analyses implicate seven individual genes, three gene sets, and four copy number variants at exome-wide significance. Genes encoding ion channels show strong association with multiple epilepsy subtypes, including epileptic encephalopathies, generalized and focal epilepsies, while most other gene discoveries are subtype-specific, highlighting distinct genetic contributions to different epilepsies. Combining results from rare single nucleotide/short indel-, copy number-, and common variants, we offer an expanded view of the genetic architecture of epilepsy, with growing evidence of convergence among different genetic risk loci on the same genes. Top candidate genes are enriched for roles in synaptic transmission and neuronal excitability, particularly postnatally and in the neocortex. We also identify shared rare variant risk between epilepsy and other neurodevelopmental disorders. Our data can be accessed via an interactive browser, hopefully facilitating diagnostic efforts and accelerating the development of follow-up studies.

## Introduction

Epilepsy is a group of heterogeneous disorders, characterized by an enduring predisposition to generate epileptic seizures.^1^ Epilepsy has a prevalence of 4-10 per 1,000 individuals worldwide, making it one of the most common neurological conditions.^2^ The role of genetic contributions to epilepsy has been long recognized,^3,4^ yet delineating the full range of genetic effects on the epilepsies remains a core challenge.

Whole-exome sequencing (WES) has proven effective in gene discovery for Mendelian disorders, including familial and severe epilepsy syndromes. There has been an increasing number of genes implicated in the developmental and epileptic encephalopathies (DEEs, [MIM: 308350]), a severe group of epilepsies characterized by early-onset, intractable seizures and developmental delay. In contrast, genes discovered for the milder, more common forms of epilepsies – genetic generalized epilepsy (GGE [MIM: 600669]) and non-acquired focal epilepsy (NAFE [MIM: 604364, 245570]) characterized by generalized and focal seizures, respectively – remain scarce.^5^ Most discoveries have been based on familial cases with limited sample sizes, and/or hypothesis-driven approaches that are focused on one or a few predefined candidate genes.^6,7^ Hypothesis-free, large-scale WES analyses have only recently been enabled and expanded through global consortia efforts.^6,8–10^

In this study, we present the largest WES analysis of epilepsy to date, from the Epi25 Collaborative, a global collaboration committed to sequencing and deep-phenotyping 25K individuals with epilepsy. Our previous data collection and analysis of ∼17K and ∼29K individuals in case-control cohorts have revealed rare coding variants confer risk for all three major subtypes of non-lesional epilepsies (DEEs, GGE, and NAFE). Here, we expand the evaluation to ∼54K individuals, comprising 20,979 cases and 33,444 matched controls spanning six genetic ancestries, with improved power for detecting “ultra-rare” variant (URV) association. We apply a hypothesis-free approach to test association between URVs (single nucleotide variants [SNVs] and short insertions/deletions [indels]) and cases-controls status, at individual-gene, gene-set level and exome-wide, and pursue these analyses separately for each subtype of epilepsy. With the enlarged sample size, we discover exome-wide significant genes for different epilepsies, identifying both shared and distinct rare variant risk factors. Integrating these findings with associations implicated by copy number variants (CNVs) and genome-wide association study (GWAS), we identify convergence of different types of genetic risk factors in the same genes. Spatiotemporal brain transcriptome analysis of WES- and GWAS-implicated genes show consistent expression patterns highlighting the neocortex and the postnatal development period. More broadly, comparing results to other large-scale WES studies, we provide significant evidence for an overlapping rare variant risk between epilepsy and other neurodevelopmental disorders (NDDs). Together, our WES analysis at the unprecedented scale makes an important step forward in discovering rare variant risk underlying a spectrum of epilepsy syndromes and offers a valuable resource for generating hypotheses about syndrome-specific etiologies.

## Results

### Study overview

We performed WES and harmonized variant detection of an initial dataset of over 70,000 epilepsy-affected and control individuals recruited across 59 sites globally. After stringent quality control (QC; Methods), we included a total of 20,979 individuals with epilepsy and 33,444 controls without known neurological or neuropsychiatric conditions in our URV association analysis, roughly doubling the sample size in our last release of Epi25 WES study.^10^ The samples were predominantly of European genetic ancestries (76.6% non-Finnish and 2.7% Finnish), with smaller proportions of African (7.7%), East Asian (5.3%), South Asian (1.1%), and Admixed American (6.6%) genetic ancestries. Epilepsy cases were matched with controls of the same genetic ancestry and samples were pooled for a joint burden analysis (Supplementary Table 1-3 and Supplementary Figure 1-5).

In the primary analysis, we evaluated the excess of ultra-rare, deleterious SNVs and indels – protein-truncating/damaging missense (MPC^11^ score ≥2) variants observed at no more than five copies among the entire dataset – in individuals with epilepsy compared to controls, using a Firth logistic regression model with adjustment for sex and genetic ancestry (Methods). We performed the association analyses separately for each epilepsy subtype – including 1,938 individuals with a diagnosis of a DEE, 5,499 with GGE, and 9,219 with NAFE – as well as for all epilepsy-affected individuals combined (including an additional 4,323 with other epilepsy syndromes). Stringent Bonferroni correction was applied to adjust for 18,531 consensus coding sequence (CCDS) genes and 5,373 gene sets, each multiplied by eight case-control comparisons across four epilepsy groups and two variant classes. To ensure our model was well calibrated, we used ultra-rare synonymous variants as a negative control for all tests (Extended Data Fig. 1). In a similar fashion, we performed CNV discovery and burden analysis on the same dataset (Methods). Moreover, we performed secondary subgroup burden analyses, splitting the data by genetic ancestry and sex. We focused on the primary, most-powered analyses in our main result section while presenting the secondary analyses in Extended Data.

### Gene-based burden identifies exome-wide significant genes

For gene discovery, we tested the burden of URVs in each protein-coding gene, across all three epilepsy subtypes and all-epilepsy combined (Supplementary Data 1). In the analysis of protein-truncating URVs in DEEs, we identified five genes at exome-wide significance (**Fig. 1a** and Table 1a; Methods): *NEXMIF* ([MIM: 300524], log[OR]=6.7, *P<*2.2×10^-16^), *SCN1A* ([MIM: 182389], log[OR]=4.1, *P=*6.3×10^-9^), *SYNGAP1* ([MIM:603384], log[OR]=4.2, *P=*5.9×10^-8^), *STX1B* ([MIM: 601485], log[OR]=4.5, *P=*2.3×10^-7^), and *WDR45* ([MIM: 300526], log[OR]=5.5, *P=*2.4×10^-7^). All five are well-established epilepsy genes, reviewed by the GMS Genetic Epilepsy Syndromes panel^12^ with diagnostic level of evidence. *NEXMIF* and *SCN1A* have been consistently the top genes in our prior Epi25 analyses;^9,10^ the other three genes for the first time surpassed the exome-wide significance threshold. The 6^th^ ranked gene – *ANKRD11* ([MIM: 611192]), which approaches exome-wide significance (log[OR]=3.9, *P=*1.2×10^-6^) – emerged as a novel candidate for DEEs.

**Fig. 1:**
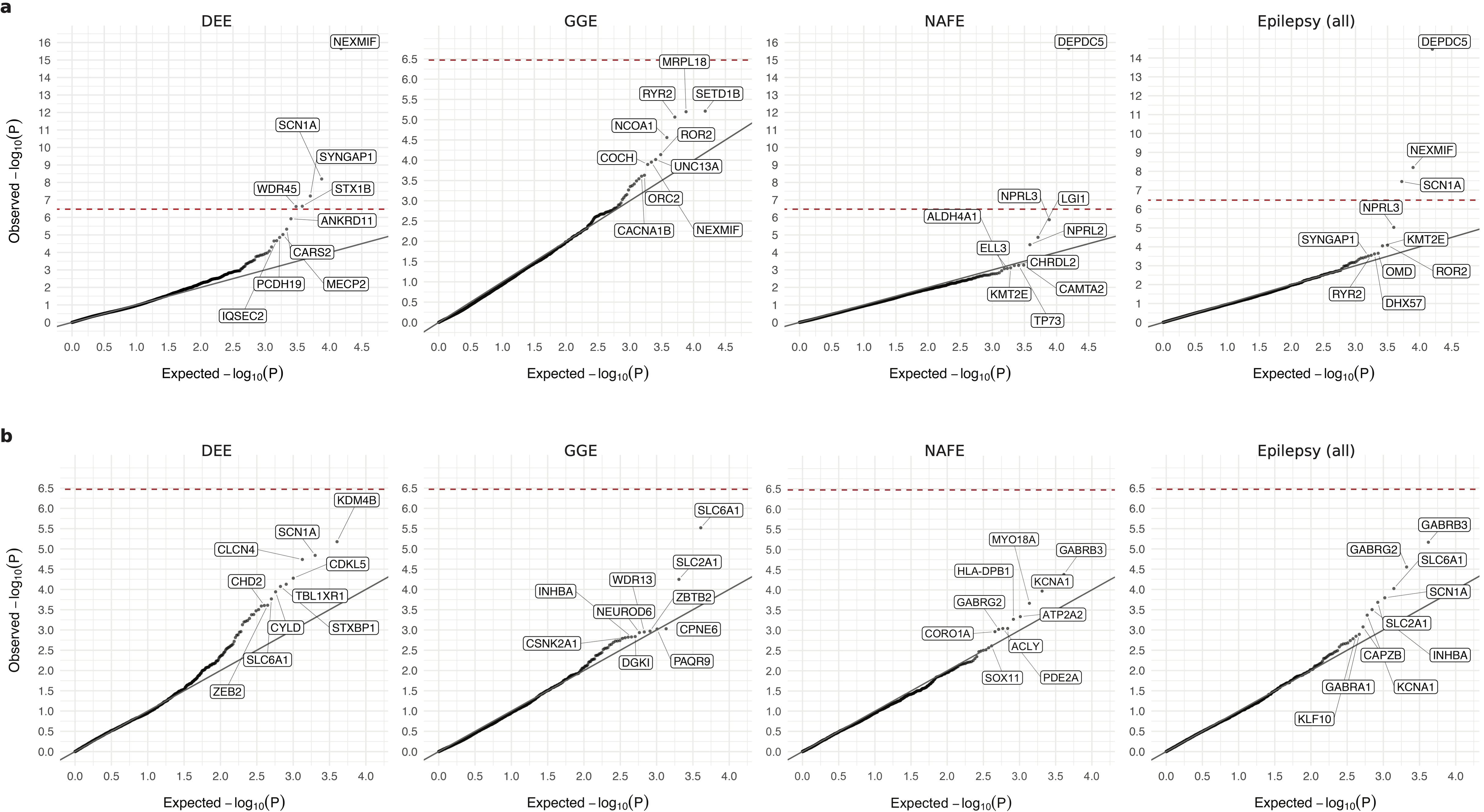
Results from gene-based burden analysis of URVs. **a**,**b**, Burden of protein-truncating (**a**) and damaging missense (**b**) URVs in each protein-coding gene with at least one epilepsy or control carrier. The observed −log_10_-transformed *P* values are plotted against the expectation given a uniform distribution. For each variant class, burden analyses are performed across four epilepsy groups – 1,938 DEEs, 5,499 GGE, 9,219 NAFE, and 20,979 epilepsy-affected individuals combined – versus 33,444 controls. *P* values are computed using a Firth logistic regression model testing the association between the case-control status and the number of URVs (two-sided); the red dashed line indicates exome-wide significance *P*=3.4×10^-7^ after Bonferroni correction (see Methods). Top ten genes with URV burden in epilepsy are labeled.

**Table 1:**
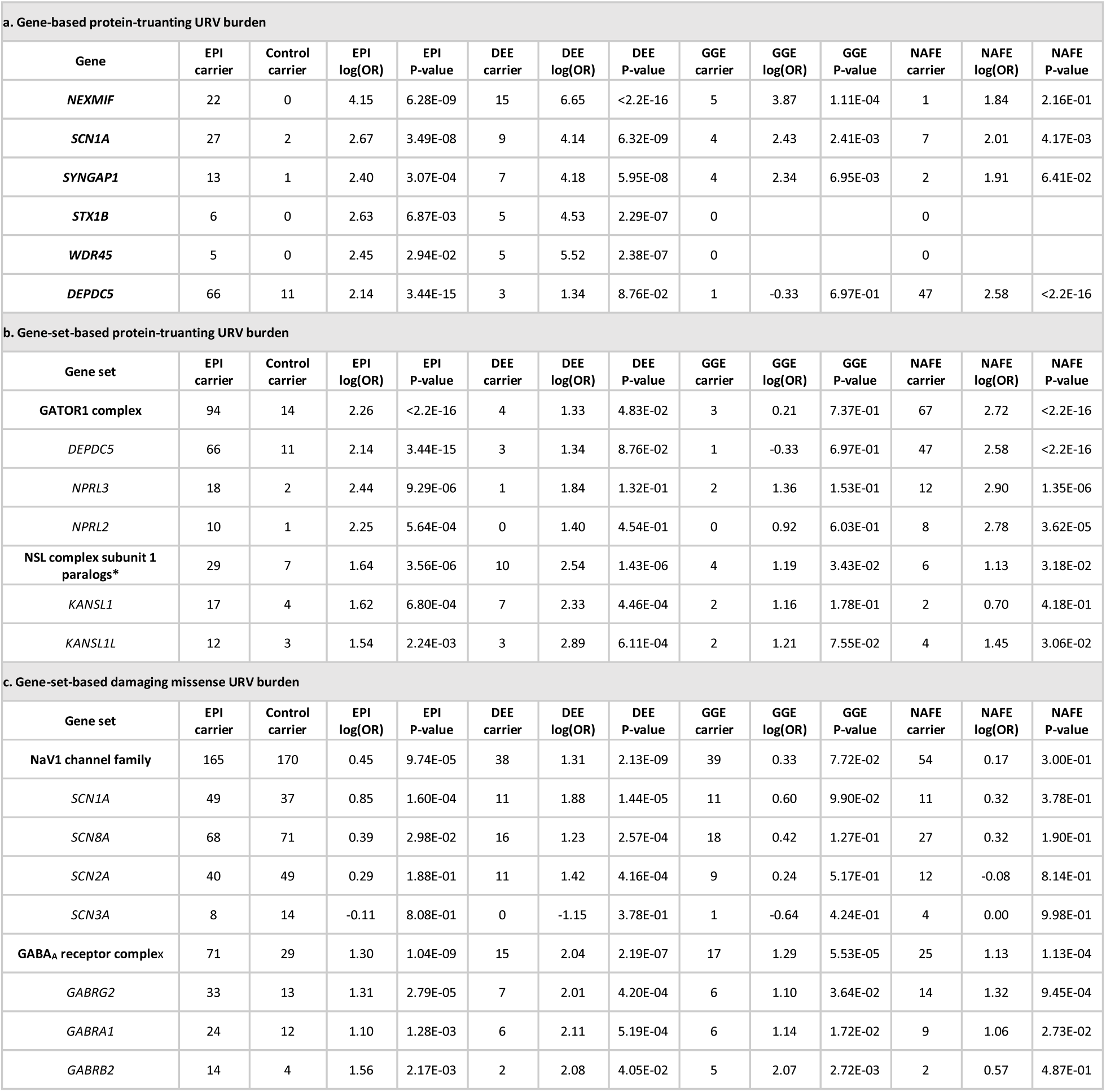
Genes and gene sets identified by exome-wide gene burden analyses. **a**, Genes with significant burden of protein-truncating URVs. **b**,**c**, Gene sets with significant burden of protein-truncating (**b**) and damaging missense (**c**) URVs. Genes and gene sets with exome-wide significance are shown (*P*-value <3.4E-7 for gene-based burden testing and *P*-value <1.2E-6 for gene-set-based burden testing); asterisk indicates a *P*-value very close to the exome-wide significance. *P*-values are reported as <2.2E-16 for extremely small values below the precision threshold of the statistical software. The number of mutation carriers, log odds ratio (OR), and *P*-value are listed for each burden test across four epilepsy groups.

Analysis of protein-truncating URVs in NAFE showed the strongest association evidence for *DEPDC5* ([MIM: 614191], log[OR]=2.6, *P<*2.2×10^-16^; **Fig. 1a** and Table 1a), which encodes part of the GATOR1 complex, a repressor of the mTORC1 pathway that has been prominently associated with focal epilepsies.^7^ The other two components of the GATOR1 complex, *NPRL3* (MIM: 600928) and *NPRL2* (MIM: 607072), were also among the top associations (*NPRL3*: log[OR]=2.9, *P=*1.4×10^-6^, *NPRL2*: log[OR]=2.8, *P=*3.6×10^-5^), and they together manifested the strongest burden in the subsequent gene-set-based analysis. Notably, *DEPDC5* was the only exome-wide significant hit in the earlier Epi4K WES study of familial NAFE cases;^8^ the expanded inclusion of non-familial cases in our cohort implicates *DEPDC5* in both familial and non-familial cases (Supplementary Data 2), reinforcing the notion that sporadic and familial forms of epilepsy have shared genetic risk.

No genes surpassed the exome-wide significance threshold in GGE. Three genes remained significant when we combined all epilepsy subtypes (**Fig. 1a**): *DEPDC5* (log[OR]=2.1, *P=*3.4×10^-15^), *NEXMIF* (log[OR]=4.1, *P=*6.3×10^-9^), and *SCN1A* (log[OR]=2.7, *P=*3.5×10^-8^). The signals of enrichment became slightly attenuated compared to the epilepsy subtype-specific analysis, which may reflect the genetic and etiological heterogeneity of different epilepsies.

In comparison to protein-truncating URVs, burden analysis of damaging missense URVs did not identify individual genes at exome-wide significance. Nevertheless, the top associations captured known epilepsy genes – notably the *SLC6A1* (MIM: 137165) and *GABRB3* (MIM: 137192) genes, both involved in the GABAergic pathway^9^ and showing enrichment across multiple epilepsy subtypes (**Fig. 1b**). Most of the previously implicated variants in these two genes were also missense,^13,14^ and our study discovered an additional 24 and 26 damaging missense URVs in *SLC6A1* and *GABRB3*, respectively, increasing the existing candidates by ∼50% (Supplementary Data 3). Another top hit – *KDM4B* (MIM: 609765) – was found specifically associated with DEEs, which has not been previously reported.

### Gene-set-based burden facilitates biological interpretation

To further investigate biologically relevant pathways associated with epilepsy, we performed burden tests at a gene-set level. Different from our prior Epi25 analyses, which focused on a few prioritized gene sets,^15^ we systematically tested collections of gene entities that belong to a gene family^16^ or encode a protein complex^17^ (Supplementary Data 4; Methods), in search for novel associations.

The most pronounced signal, as described in the gene-based burden of protein-truncating URVs, was from the GATOR1 complex in NAFE (log[OR]=2.7, *P<*2.2×10^-16^; **Fig. 2a** and Table 1b). We identified a total of 56 distinct protein-truncating URVs in GATOR1 (Supplementary Data 5), among which 45 appeared novel according to the most recent study of epilepsy-related GATOR1 variants by Baldassari et al.^7^ In contrast to Baldassari et al, where most (>70%) GATOR1 protein-truncating variant carriers were familial, only 20% of the carriers in our study cohort had a known family history of epilepsy. Both familial and non-familial cases showed significant burden of GATOR1 protein-truncating URVs (Supplementary Data 2), highlighting the role of GATOR1 genes in the etiology of focal epilepsy.

**Fig. 2:**
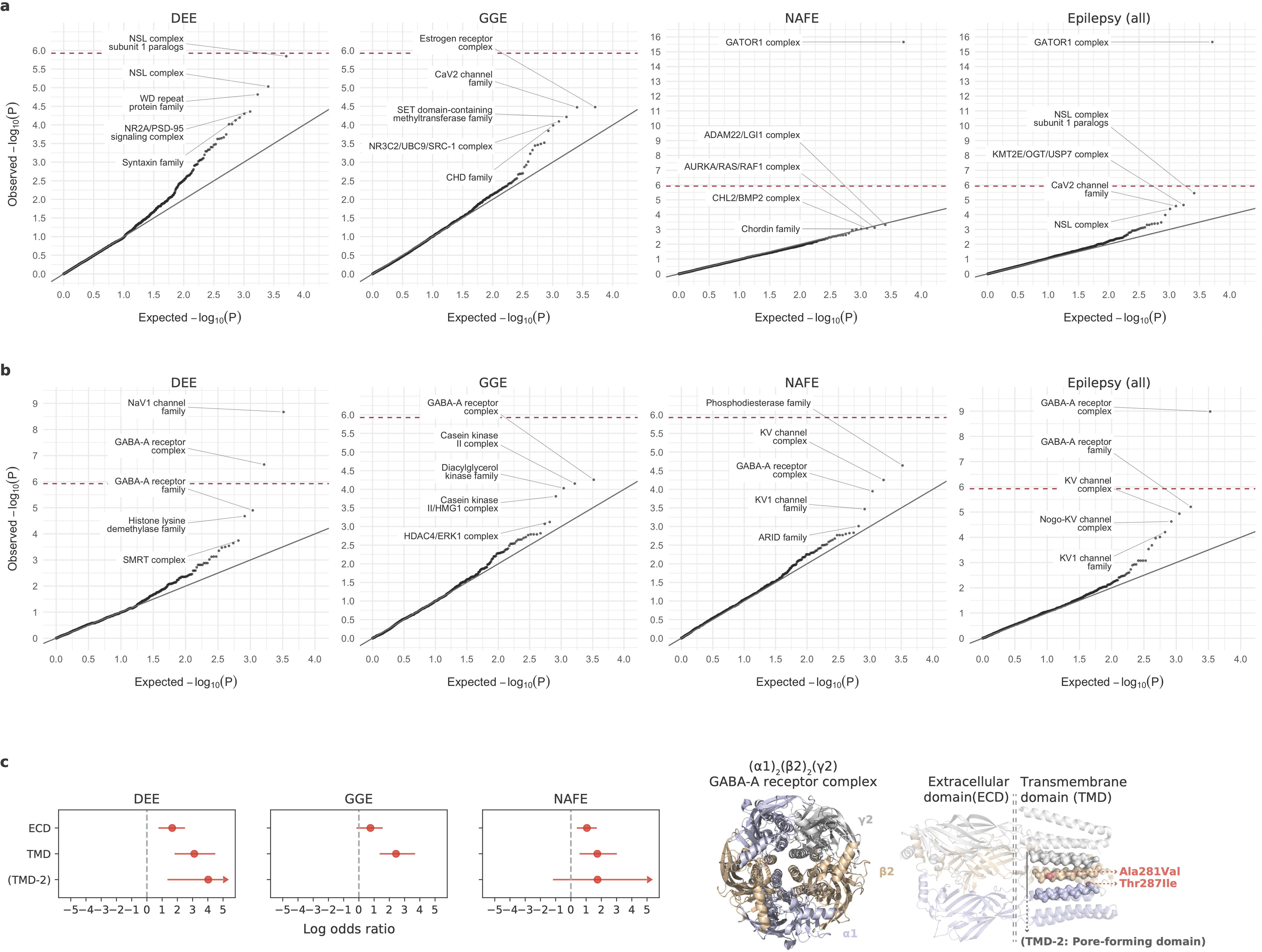
Results from gene-set-based burden analysis of URVs. **a**,**b**, Burden of protein-truncating (**a**) and damaging missense (**b**) URVs in each gene set (gene family/protein complex) with at least one epilepsy or control carrier. The observed −log_10_-transformed *P* values are plotted against the expectation given a uniform distribution. For each variant class, burden analyses are performed across four epilepsy groups – 1,938 DEEs, 5,499 GGE, 9,219 NAFE, and 20,979 epilepsy-affected individuals combined – versus 33,444 controls. *P* values are computed using a Firth logistic regression model testing the association between the case-control status and the number of URVs (two-sided); the red dashed line indicates exome-wide significance *P*=1.2×10^-6^ after Bonferroni correction (see Methods). Top five gene sets with URV burden in epilepsy are labeled. **c**, Burden of damaging missense URVs in the (α1)_2_(β2)_2_(γ2) GABA_A_ receptor complex with respect to its structural domain. Left, forest plots showing the stronger enrichment of damaging missense URVs in the transmembrane domain (TMD) than the extracellular domain (ECD), and the unique signal from DEEs in the second TMD (TMD-2) that forms the ion channel pore. The dot represents the log odds ratio and the error bars represent the 95% confidence intervals of the point estimates. For presentation purposes, error bars that exceed a log odds ratio of 5 are capped, indicated by arrows at the end of the error bars (see Supplementary Data 6 for exact values). Right, a co-crystal structure (PDB ID: 6X3Z) showing the pentameric subunits of the receptor and highlighting the two protein-truncating URVs from DEEs located in the pore-forming domain.

Several strong signals emerged in the analysis of damaging missense URVs, led by well-established ion channel protein complexes and gene families (**Fig. 2b** and Table 1c). The top association was the GABA_A_ receptor complex, encoded by *GABRA1* (MIM: 137160), *GABRB2* (MIM: 600232), and *GABRG2* (MIM: 137164; [α1]_2_[β2]_2_[γ2], the most abundantly expressed isoform in the brain),^18^ which controls the majority of inhibitory signaling in the central nervous system. The complex showed extensive enrichment across all epilepsy subtypes, recapturing the pervasive role of GABA_A_ receptors across the spectrum of severities in epilepsy.^19^ Further dissecting the signals with respect to the structural domain of the complex, we observed stronger URV burden in the transmembrane domains than the extracellular domain, particularly for DEEs and GGE; and DEEs exhibited a unique signal in the second transmembrane -helix lining the ion channel pore of the receptor^20^ (**Fig. 2c**; Supplementary Data 6). These patterns collectively point to an association of damaging missense URVs in the pore-forming domain with a more severe form of epilepsy.

Potential novel associations were found in two gene sets: the NSL complex (with protein-truncating URVs in *KANSL1* [MIM: 612452], *KANSL2* [MIM: 615488], and *PHF20* [MIM: 610335]) and the phosphodiesterase (PDE) gene family (with damaging missense URVs in *PDE2A* [MIM: 602658] and *PDE10A* [MIM: 610652]), associated with DEEs and NAFE, respectively. Despite the sparsity of URVs, our results broaden the potential allelic spectrum of variants that may confer risk for different subtypes of epilepsies.

### Protein structural analysis characterizes missense URVs

The strong burden of damaging missense, but not protein-truncating, URVs in genes encoding ion channels suggests a pathophysiological mechanism of protein alteration rather than haploinsufficiency. Given the specialized structure of ion channels, we further characterized missense URVs at a protein structure level. We collected experimentally resolved 3D structures of ion channel proteins, and applied Rosetta^21^ to assess the Gibbs free energy changes (ΔΔG, or ddG in abbreviation) of protein folding upon a particular missense URV; a decrease in Gibbs free energy of unfolding, i.e., a positive ddG value suggests a destabilizing effect of the variant on protein and a negative value suggests a stabilizing effect. We computed ddG for a total of 1,782 missense URVs across 16 ion channel protein complexes (Supplementary Data 7; Methods).

There was a positive correlation between ddG and MPC (ρ=0.15, *P=*8.3×10^-11^; **Fig. 3a**), nonetheless, incorporating ddG further stratified epilepsy association signals (**Fig. 3b**; Supplementary Data 8). Significant enrichment was found for both destabilizing (ddG ≥1 kcal/mol) and stabilizing (ddG ≤-1 kcal/mol) URVs, suggesting a diverging molecular mechanism for these missense URVs. Dissecting the signals by protein structural domains, we found divergent distributions for destabilizing and stabilizing missense URVs, with the former enriched in the ECD of the complex and the latter in the TMD (**Fig. 3c**; Supplementary Data 9). These results may provide testable hypotheses about how ion channel dysfunction could produce a broad range of epilepsy syndromes.

**Fig. 3:**
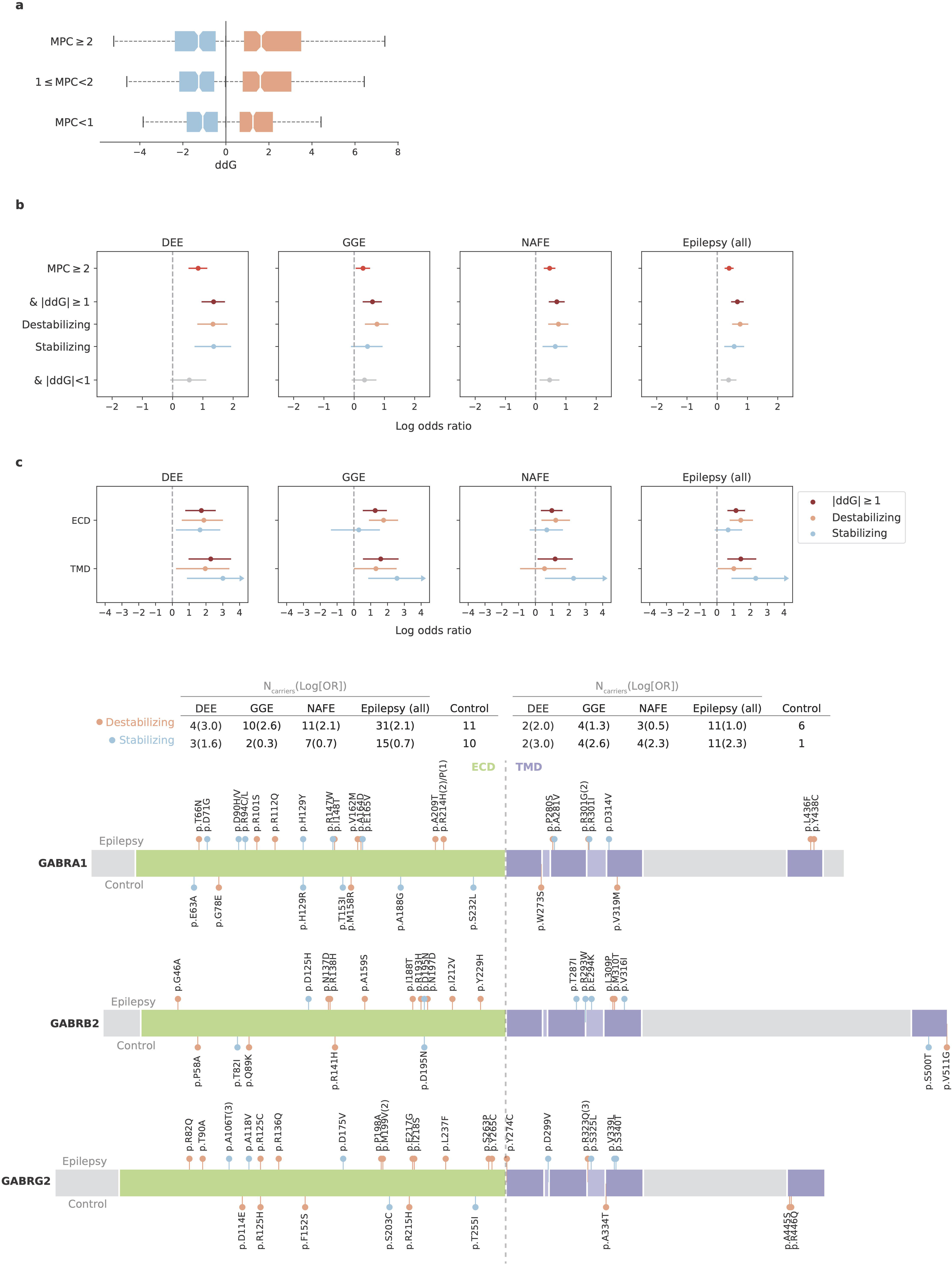
Protein structural analysis of missense URVs in ion channel genes. **a**, Correlation between ddG and MPC in measuring the deleteriousness of missense URVs. A higher absolute ddG value suggests a more deleterious effect on protein stability; positive (orange) and negative (blue) values suggest destabilizing and stabilizing effects, respectively. Box plots show the distribution of ddG values across different MPC ranges (blue boxes: N=232, 272, and 242 for MPC<1, 1≤MPC<2, and MPC≥2, respectively; orange boxes: N=327, 397, and 342 or MPC<1, 1≤MPC<2, and MPC≥2, respectively). The center line represents the median (50th percentile) and the bounds of the box indicate the 25th and 75th percentiles, with the whiskers extending to the minimum and maximum values within 1.5 times the interquartile range from the lower and upper quartiles, respectively. **b**, Burden of damaging missense URVs stratified by ddG. Stronger enrichment is observed when applying |ddG|≥1 to further prioritize damaging missense URVs with MPC≥2. **c**, Burden and distribution of destabilizing (ddG≥1) and stabilizing (ddG≤-1) missense URVs on the (α1)_2_(β2)_2_(γ2) GABA_A_ receptor complex with respect to its structural domain. Top, forest plots showing the stronger enrichment of destabilizing missense URVs (orange) in the extracellular domain (ECD) and stabilizing missense URVs (blue) in the transmembrane domain (TMD). Bottom, schematic plots displaying the distribution of destabilizing and stabilizing missense URVs on GABAA receptor proteins. URVs found in epilepsy cases are plotted above the protein and those from controls are plotted below the protein. The number of epilepsy and control carriers are listed in the table above. In **b** and **c**, burden analyses are performed across four epilepsy groups – 1,938 DEEs, 5,499 GGE, 9,219 NAFE, and 20,979 epilepsy-affected individuals combined – versus 33,444 controls. The dot represents the log odds ratio and the error bars represent the 95% confidence intervals of the point estimates.

### CNV deletion burden converges with protein-truncating URVs

In parallel with SNVs and indels, we performed variant calling of CNVs on the same dataset (Methods). After sample QC, we examined the burden of rare CNVs in 18,963 epilepsy cases – including 1,743 DEEs, 4,980 GGE, and 8,425 NAFE – versus 29,804 controls (∼90% of initial; Methods). We first tested a curated set of 79 CNVs previously associated with NDDs, known as “genomic disorders” (GD).^22^ Four GD loci were significantly enriched in epilepsy cases: three from the common complex forms of epilepsies (16p13.11 deletion, 15q13.2-q13.3 deletion, and 17q12 duplication) and one from DEEs (15q11.2-q13.1 duplication). All four loci have been prominent in previous reports as predisposing to a diverse range of epilepsy syndromes.^23–26^

Moreover, we evaluated CNV burden at the individual gene level; a gene was considered affected by a CNV if 10% of its coding exons were deleted or 75% were duplicated. The most significant signal was from CNV deletions in the *NPRL3* gene, with 11 deletions found in NAFE cases versus 0 in controls (log[OR]=4.1, *P=*9.4×10^-7^; Supplementary Data 10). Notably, *NPRL3* was also one of the top hits implicated by protein-truncating URVs in NAFE, and jointly analyzing the two identified *NPRL3* a new exome-wide significant gene (log[OR]=3.8, *P=*8.1×10^-12^; **Fig. 4a**). Among the top ten genes with protein-truncating URV burden across four epilepsy groups, about one-third (14/40) were found affected by a CNV deletion, and the vast majority (11/14) showed enrichment in epilepsy cases (**Fig. 4b**). These included *DEPDC5*, which together with *NPRL3* reinforces a haploinsufficiency mechanism for GATOR1-related focal epilepsies (**Fig. 4c**). Strengthened burden was also found for potential novel genes – e.g., *CARS2* (MIM: 612800) in DEEs and *NCOA1* (MIM: 602691) in GGE, both with supporting evidence from previous studies.^27–29^ Analysis of CNV duplications did not show any individual genes close to exome-wide significance (Supplementary Data 10).

**Fig. 4:**
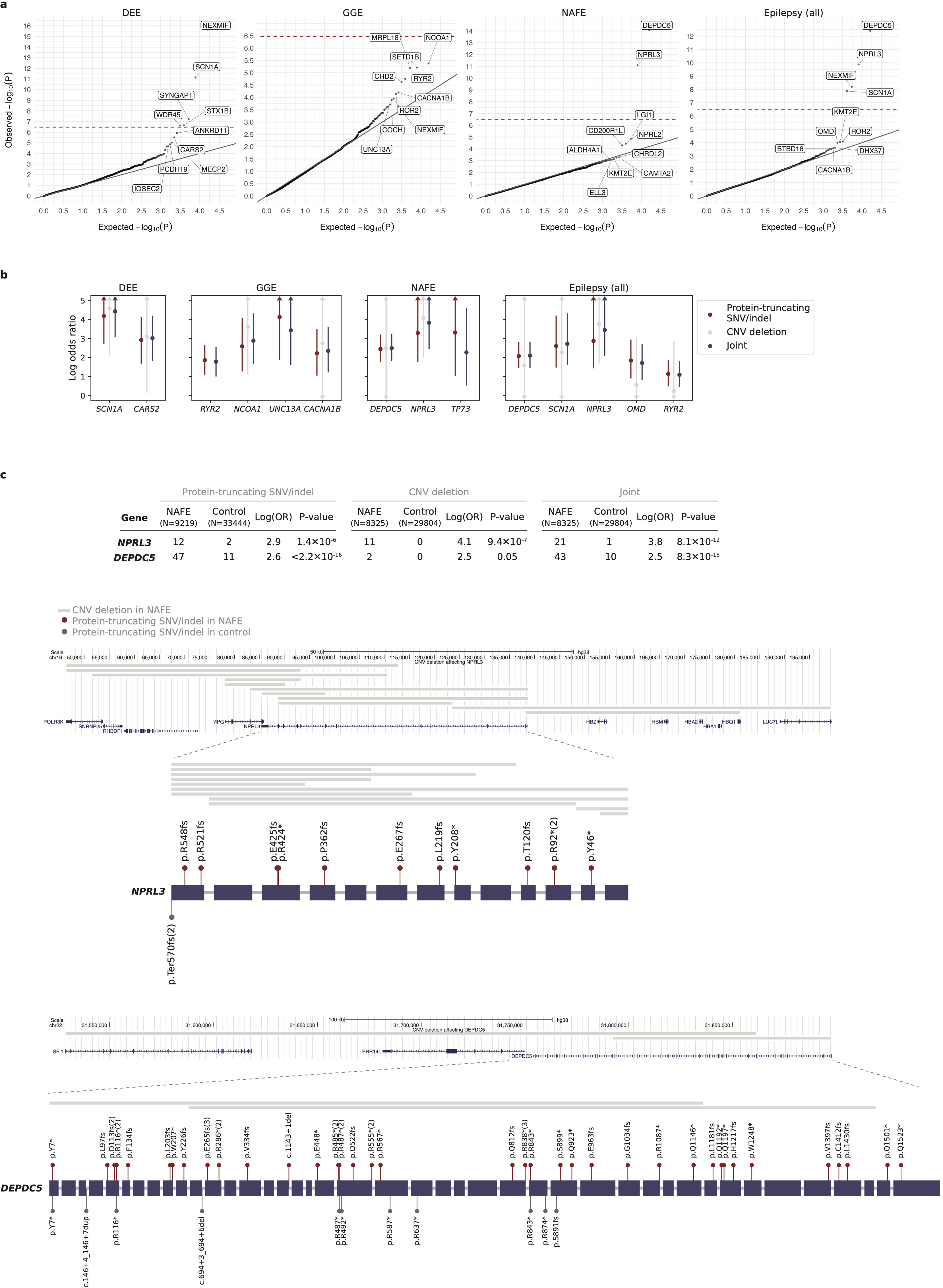
Convergence of CNV deletions and protein-truncating URVs in gene-based burden. **a**, Joint burden of CNV deletions and protein-truncating URVs in each protein-coding gene with at least one epilepsy or control carrier. The observed −log_10_-transformed *P* values are plotted against the expectation given a uniform distribution. Joint burden analyses are performed on the subset of samples that passed CNV calling QC (see Methods), across four epilepsy groups – 1,743 DEEs, 4,980 GGE, 8,425 NAFE, and 18,963 epilepsy-affected individuals combined – versus 29,804controls; for genes that do not have a CNV deletion called, results from the burden analysis of protein-truncating URVs on the full sample set are shown. *P* values are computed using a Firth logistic regression model testing the association between the case-control status and the number of URVs (two-sided); the red dashed line indicates exome-wide significance *P*=3.4×10^-7^ after Bonferroni correction (see Methods). Top ten genes with variant burden in epilepsy are labeled. **b**, Joint burden of CNV deletions and protein-truncating URVs in the top ten genes ranked by protein-truncating URV burden. Only genes affected by both variant types with enrichment in epilepsy (log[OR]>0) are show. For comparison, the burden of protein-truncating URVs (SNVs/indels; red), CNV deletions (gray), and the joint (purple) are analyzed on the same sample subset as described in **a**. The dot represents the log odds ratio and the error bars represent the 95% confidence intervals of the point estimates. For presentation purposes, error bars that exceed a log odds ratio of 5 are capped, indicated by arrows at the end of the error bars (see Supplementary Data 10 for exact values). **c**, Genomic location and distribution of CNV deletions and protein-truncating URVs with respect to the *NPRL3* and *DEPDC5* genes. Variants found in epilepsy cases (red) are plotted above the schematic gene plots and those from controls (gray) are plotted below the gene. The number of epilepsy and control carriers are listed in the table above. *P* values are computed using a Firth logistic regression model testing the association between the case-control status and the number of URVs (two-sided).

### An expanded view of epilepsy genetic architecture

Similar to other common NDDs, the common forms of epilepsy – GGE and NAFE – have both common and rare genetic risk factors. In partnership with the International League Against Epilepsy (ILAE) Consortium on Complex Epilepsies, we recently performed the largest GWAS meta-analysis of over 29K individuals with common epilepsies,^30^ which revealed 26 genome-wide significant common risk loci (MAF>1%). Together with the rare variant findings in this study, they constitute an expanded view of the genetic architecture of epilepsy (**Fig. 5a**). As natural selection purges deleterious variants from human populations, rarer variants exhibited larger disease effects: protein-truncating URVs (MAF<0.005%) had the largest effect sizes (median OR=79), pinpointing specific genes at exome-wide significance; rare CNVs (MAF<0.1%), known for genomic disorders, also showed large effects in increasing epilepsy risk (median OR=26); and common variants each individually had a small contribution (median OR<1.1).

**Fig. 5:**
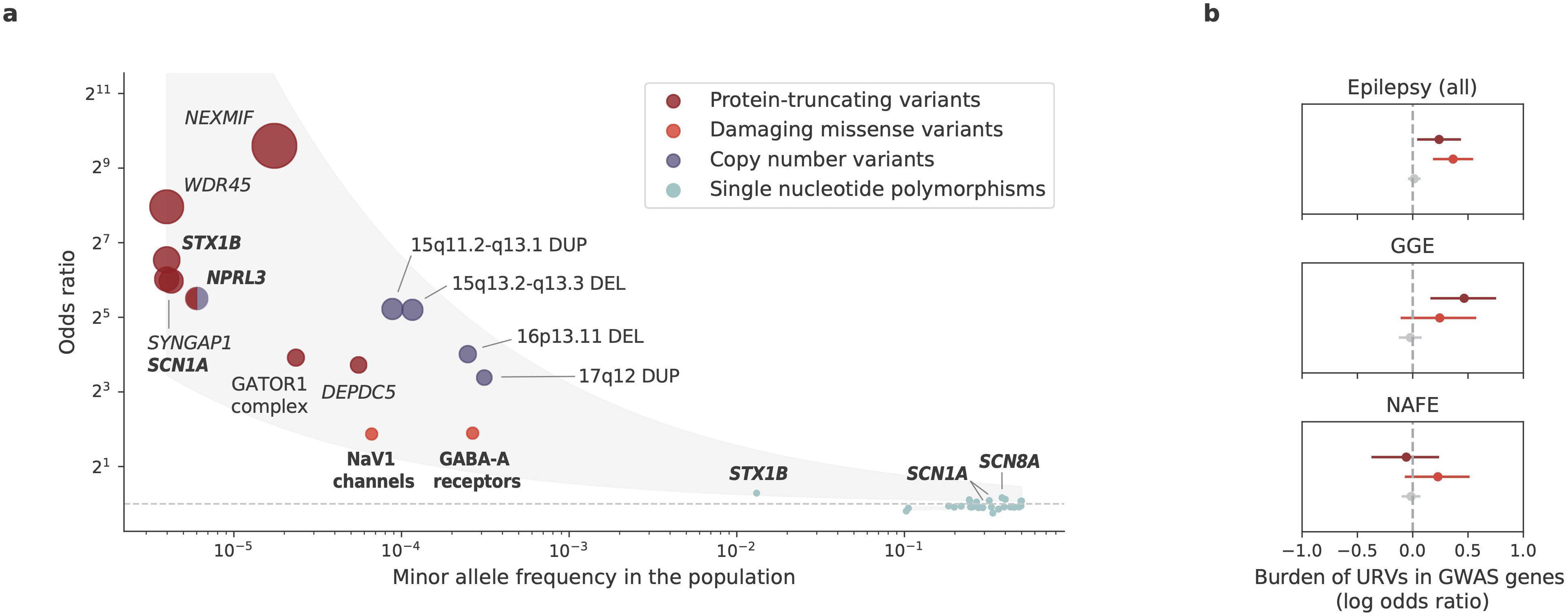
Epilepsy genetic architecture from large-scale genetic association studies. **a**, An allelic spectrum of epilepsy genetic risk loci. Significant risk loci identified by large-scale WES and GWA studies are shown. The odds ratio of each risk loci (y-axis) is plotted against the minor allele frequency in the general population (gnomAD non-neuro subset, x-axis); for individual genes, the cumulative allele frequency (CAF) is computed, and for gene sets, the CAF is averaged over gene members. The color and size of each dot represent the variant class and effect size (odds ratio) of the genetic association. Bold indicates convergent findings between different variant classes. The shaded area represents the upper and lower 95% confidence intervals of the point estimates, fitted by exponential curves. **b**, Burden of URVs in genes implicated by GWAS loci. Significant enrichment is observed for URVs from epilepsy-affected individuals in 29 GWAS genes (upper: 20,979 cases versus 33,444 controls), URVs from GGE in the 23 GGE-specific GWAS genes (middle: 5,499 GGE versus 33,444 controls), but not for URVs from NAFE in GGE GWAS genes (bottom: 9,219 NAFE versus 33,444 controls); and significance was only seen for protein-truncating (red) and damaging missense (orange) URVs but not for synonymous URVs (gray). The dot represents the log odds ratio and the error bars represent the 95% confidence intervals of the point estimates.

With new discoveries expanding the allelic spectrum, we found emerging evidence for convergence between different classes of genetic risk variants. Besides the *NPRL3* gene highlighted by the joint CNV+SNV burden analysis, three (out of 29) genes – *SCN1A*, *SCN8A*, and *STX1B* – prioritized from the GWAS loci overlapped with genes enriched with URVs, contributing to an overall URV burden in GWAS genes (**Fig. 5b**). Further delineating the analysis to epilepsy subtypes, we observed significant enrichment for URVs from GGE in the GGE-specific GWAS genes, whereas none for URVs from NAFE (**Fig. 5b**; Supplementary Data 11). This result suggests that the convergence of common and rare variant risk tends to be epilepsy subtype-specific.

At the individual gene level, 13 of the 23 GGE GWAS risk genes showed an excess of protein-truncating URVs (Supplementary Data 11). The lead gene was *RYR2* (MIM: 180902), in which 14 protein-truncating URVs were observed in our GGE cohort (log[OR]=1.8, *P=*8.6×10^-6^), with the GWAS hit residing in the intronic region (rs876793). Mutations in *RYR2* have been well-known in the etiology of arrhythmogenic disorders,^31^ while more recent studies reported that the same mutation can cause GGE independent of arrhythmias.^32,33^ Our finding adds weight to the hypothesis that *RYR2* mutations likely constitute a neuro-cardiac calcium channelopathy,^32,33^ where mutant receptors may induce either arrhythmias or GGE depending on their selective expression in the heart or in the brain.

### Functional roles of candidate genes in neural circuitry

To gain more functional insights into the identified genetic associations, we surveyed the spatiotemporal expression of top candidate genes in the human brain. We first focused on 13 genes – seven with individually exome-wide significance (*NEXMIF*, *SCN1A*, *SYNGAP1*, *STXBP1*, *WDR45*, *DEPDC5*, *NPRL3*) and six from members of significant gene families or protein complexes (*NPRL2*, *SCN2A*, *SCN8A*, *GABRA1*, *GABRB2*, *GABRG2*). We analyzed the expression of each gene across six brain structures and nine developmental periods, using the BrainSpan^34^ database (Methods). We found that the candidate genes were more highly expressed during the postnatal period than prenatally (*P*=7.0×10^-18^, Extended Data Fig. 2), and the highest expression was found in the neocortex (*P*=7.6×10^-6^, **Fig. 6a**; Methods). Consistent neocortical and developmental expression patterns were found for the 29 epilepsy risk genes discovered by GWAS^30^ (**Fig. 6a**).

**Fig. 6:**
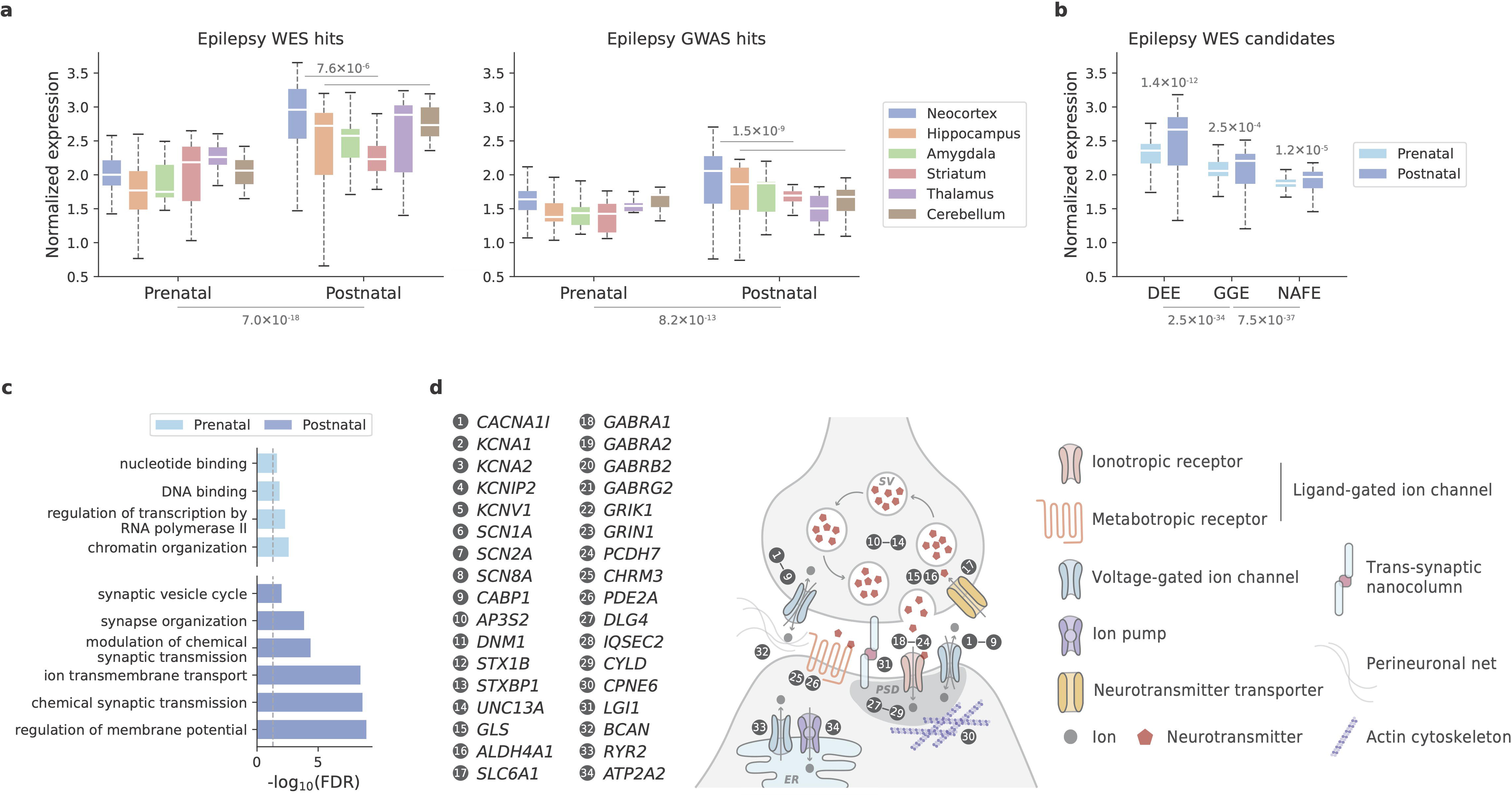
Functional analysis of candidate epilepsy genes. **a**,**b**, Spatiotemporal brain transcriptome analysis of exome/genome-wide significant genes identified in this WES study (N=13) or our recent GWA study (N=29) (**a**) and the top 20 genes enriched for deleterious URVs in each subtype of epilepsy (**b**). Candidate genes show the highest expression in the neocortex during postnatal periods. The expression values (log_2_[TPM+1]) are normalized to the mean for each BrainSpan sample and then averaged by each candidate gene set. Significance was evaluated by Wilcoxon signed rank test (N=162/200, 15/17, 14/19, 20/14, 13/16, and 13/21 for prenatal/postnatal neocortex, hippocampus, amygdala, striatum, thalamus, and cerebellum samples, respectively). Box plots indicate median, interquartile range (IQR) with whiskers adding IQR to the first and third quartiles. **c**, Gene Ontology terms enriched for candidate epilepsy genes with a prenatal- or postnatal-expression bias (N=43 and 50, respectively). Vertical dashed line indicates false discovery rate (FDR)=0.05; the full list of enriched terms is provided in Supplementary Data 12. **d**, A schematic diagram showing the distribution and function of 34 postnatally-biased genes on neuron structures. SV: synaptic vesicle, PSD: post-synaptic density, ER: endoplasmic reticulum.

Extending the comparison to the top 20 genes enriched for deleterious URVs recapitulated the postnatal expression pattern across different subtypes of epilepsy, and on average, genes implicated in the more severe subtype DEEs showed a higher expression level (**Fig. 6b**). Further quantifying the relative prenatal versus postnatal expression pattern^35^ for both WES- and GWAS-implicated genes, we classified 43 genes to have a prenatal and 50 genes with a postnatal expression preference (Supplementary Data 12). Gene Ontology enrichment analysis^36^ of these two gene sets identified two different functional categories: “gene transcription regulation” for the prenatal genes and “synaptic transmission & membrane excitability” for the postnatal genes (**Fig. 6c** and Supplementary Data 12). The latter showed overall stronger enrichment, in which 34 genes were mapped to a range of functional components of synaptic transmission and neuronal excitability (**Fig. 6d**; Methods).

For the prenatal genes with a transcriptional regulatory role, we further tested whether they regulate the postnatal genes and thus, functionally converge at neurotransmission. We constructed a gene regulatory network between these two gene sets and asked whether there are more transcription factor (TF)-target connections between them than random. Forty connections were found (Supplementary Data 12), which was significantly higher than that of a random gene set with similar brain expression profiles (Methods). The result lends support to the hypothesis about a convergent pathophysiological effect associated with these two different categories of genes in epilepsy. Meanwhile, the same TFs had a much larger number of targets beyond our candidate genes, which showed significant enrichment in a broader set of neurodevelopmental processes other than neurotransmission (Supplementary Data 12). Therefore, these results collectively implicate both developmental and functional changes of neural circuitry in the pathophysiology of epilepsy.

### Shared rare variant risk with neurodevelopmental disorders

Recent WES studies have revealed substantial rare variant risk for NDDs. Analysis of *de novo* mutations in severe developmental disorders (DDs) has discovered 285 genes at exome-wide significance,^37^ and rare variant associations in autism spectrum disorder (ASD)^22^ and schizophrenia (SCZ)^38^ have implicated 185 and 32 genes at a false discovery rate of 5%, respectively. To explore how these and our findings may point to common genetic etiologies, we examined the burden of URVs in these NDD genes (Supplementary Data 13). Significant enrichment was found for all three gene sets associated with DD, ASD, and SCZ (**Fig. 7a**), suggesting that there is shared genetic risk of rare variation among the broader spectrum of NDDs. Nine (out of 13) epilepsy significant genes overlapped with DD/ASD genes but none with SCZ (Extended Data Fig. 3 and Supplementary Data 13), suggesting a larger genetic overlapping between epilepsy and DD/ASD than SCZ, which is in line with the high comorbidity of DD/ASD and epilepsy, in particular DEEs. Given the known genetic overlapping between DD and ASD, we repeated the analyses on the subsets of mutually exclusive NDD genes (i.e., 196 DD-only, 99 ASD-only, and 22 SCZ-only genes, respectively). Although attenuated, there remained clear rare variant signals shared by epilepsy and other NDDs (Supplementary Data 13).

**Fig. 7:**
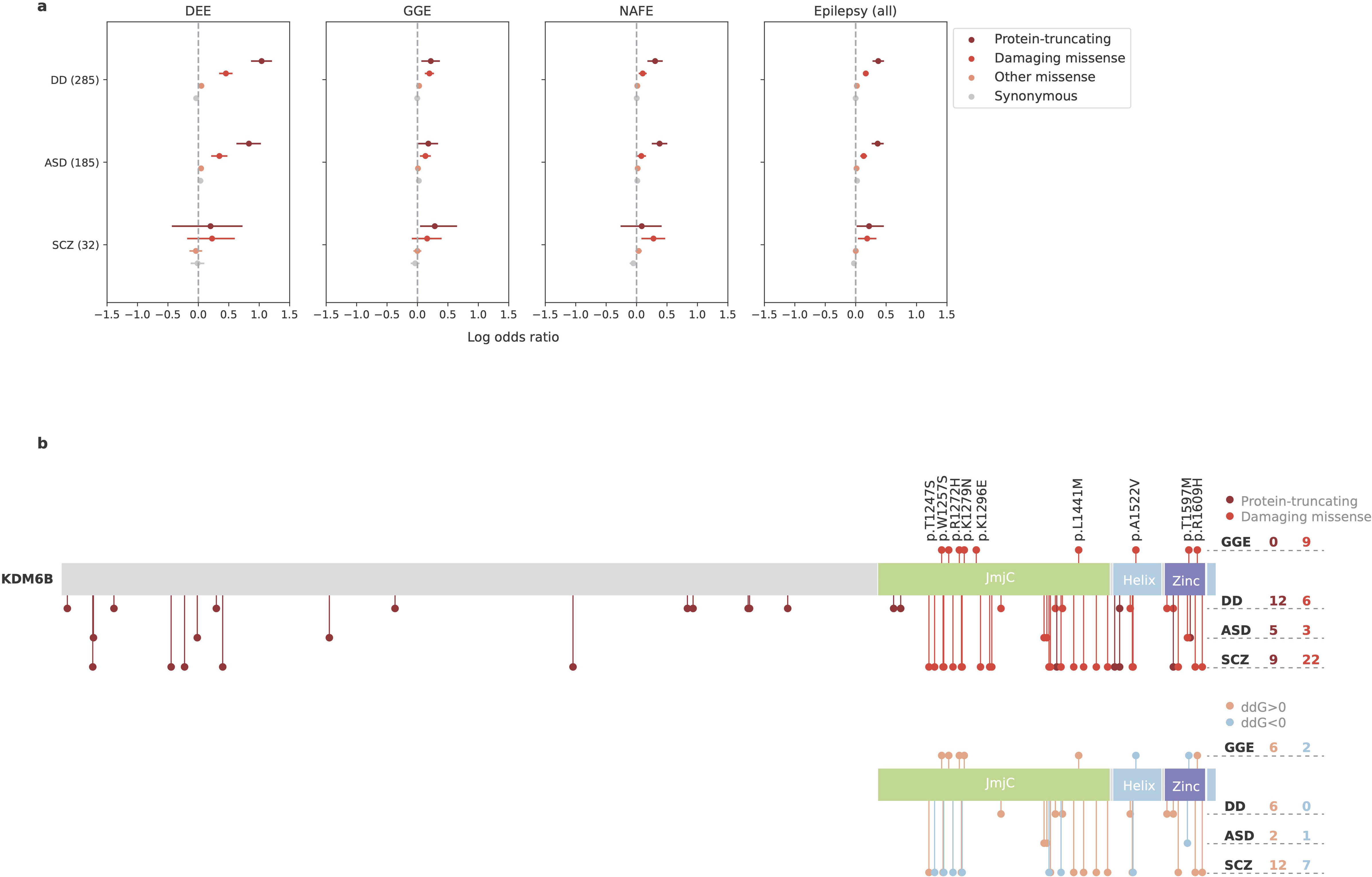
Shared rare variant risk between epilepsy and other NDDs. **a**, Burden of URVs in genes implicated by WES of severe developmental disorders (DD; N=285), autism spectrum disorder (ASD; N=185), and schizophrenia (SCZ; N=32). Burden analyses are performed across four variant classes and four epilepsy groups – 1,938 DEEs, 5,499 GGE, 9,219 NAFE, and 20,979 epilepsy-affected individuals combined – versus 33,444 controls. Overall, DD/ASD-associated genes show stronger enrichment of epilepsy URVs than SCZ. The dot represents the log odds ratio and the error bars represent the 95% confidence intervals of the point estimates. **b**, Distribution of rare variants from GGE and other NDDs on the KDM6B protein. Top, a schematic protein plot displaying the distribution of protein-truncating (darker red) and damaging missense (lighter red) variants on KDM6B. Bottom, a schematic protein plot displaying the distribution of damaging missense variants with a likely destabilizing (ddG>0; orange) and stabilizing (ddG<0; blue) effect on KDM6B. In both plots, variants found in GGE are plotted above the protein and those from other NDDs are plotted below the protein (in the order of DD, ASD, and SCZ as labeled); the number of variant carriers are listed accordingly on the right.

About one-third (136/409) NDD genes showed nominally significant enrichment of deleterious URVs in at least one epilepsy subtype (Supplementary Data 13). The vast majority (128/136=94.1%) were DD/ASD genes, and only one gene – *KDM6B* (MIM: 611577) – was shared by all three NDD gene sets. Notably, URVs in *KDM6B* associated with epilepsy were exclusively missense (MPC ≥2), whereas *KDM6B* variants implicated in DDs were predominately protein-truncating (**Fig. 7b**; Supplementary Data 13). All missense variants were clustered at the KDM6B catalytic domain (JmjC) and C-terminal helix/zinc motifs, which are important for enzyme-cofactor binding and protein stability.^39^ Protein structural analysis predicted that most of the damaging missense variants to have a destabilizing effect on the KDM6B protein (Supplementary Data 13), especially those in DDs, while diverging effects were observed for epilepsy and SCZ (**Fig. 7b**). These results suggest that, even converging in the same gene, rare variant risk may differ in its severity and/or the molecular mechanism that underlies specific phenotypes of NDDs.

### Shared rare variant risk across genetic ancestries and sexes

Finally, we repeated the burden analysis within subgroups of samples by genetic ancestry and sex. Six genetic ancestral groups were classified, comprising 76.6% non-Finnish European, 7.7% African, 6.6% Admixed American, 5.3% East Asian, 2.7% Finnish, and 1.1% South Asian samples (Extended Data Fig. 4a; Supplementary Data 14). The non-European subgroups remained underpowered to detect exome-wide significant genes, yet showed enrichment of URVs in top genes identified in the full analysis (Extended Data Fig. 4b). Significant enrichment was found in the sets of established epilepsy genes and genes that are intolerant to genetic variation, as well as in intolerant genes that are not currently linked to epilepsy (Extended Data Fig. 4c; Supplementary Data 14). These results suggest that while there likely exist shared genetic risk underlying epilepsy across different genetic ancestries, more remains to be discovered, especially for non-European ancestral groups.

Similar patterns were observed in sex-specific burden analyses. Both female and male subgroups showed significant URV burden in known epilepsy genes, with the female subgroup exhibiting stronger enrichment in X-linked genes (Extended Data Fig. 4d; Supplementary Data 15). At the individual gene level, given the similar sample size between sexes, we directly compared their burden *P* values and found three exome-wide significant genes with sex-biased URV burden: *NEXMIF* and *SCN1A* in female and *NPRL3* in male (Extended Data Fig. 4e; Supplementary Data 15). The top hit *NEXMIF* is an X-linked gene, the loss-of-function of which causes NEXMIF encephalopathy and has been shown to have markedly different manifestations between females and males^40^. Different molecular mechanisms could explain the phenotypic variability^40,41^; albeit intriguing based on the X-chromosome inactivation theory, they require further investigation with functional studies.

## Discussion

In the largest WES study of epilepsies to date, we characterize the contribution of ultra-rare genetic variation to epilepsy risk. Here, we have aggregated deeply-phenotyped epilepsy collections from across the world, sequenced them and have harmonized variant detection and quality control to power analysis and interpretation of the genetic data for etiological and clinical implications.

Our exome-wide burden analyses redemonstrated the role of known epilepsy genes with improved power and discovered potential novel rare risk variants for different subtypes of epilepsies. Most associations were identified in a particular epilepsy subtype, implicating distinct genetic etiologies underlying different epilepsies. Protein-truncating URVs exhibited the strongest signal, with six individual genes surpassing the stringent exome-wide significance threshold: five genes (*NEXMIF*, *SCN1A*, *SYNGAP1*, *STX1B*, and *WDR45*) were associated with the severe group of DEEs, while the most significant gene, *DEPDC5,* was found in NAFE. In comparison to protein-truncating URVs, analysis of damaging missense URVs remained underpowered to identify individual genes at exome-wide significance. Yet, strong associations emerged when we aggregated sets of genes that share common functions. The top associations were predominantly genes encoding ion channel complexes, such as Nav/Kv channels and GABA_A_ receptors. Notably, these gene sets did not show significant enrichment of protein-truncating URVs, suggesting more diverse molecular mechanisms than haploinsufficiency. Protein structural analysis of missense URVs in these genes suggested diverging effects on ion channel protein stability. In this study, we deliberately separated the analysis of protein-truncating and damaging missense URVs, while assuming a protein-truncating-like effect for all damaging missense URVs identified no additional significant genes but weakened our analytical power (Extended Data Fig. 5).

Potential novel associations were identified in several genes and gene sets. The top candidates were predominately implicated in DEEs, including protein-truncating URVs in *ANKRD11* and NSL complex and damaging missense URVs in *KDM4B*. The *ANKRD11* gene is a known causal gene for the KBG syndrome,^42^ a rare genetic disorder characterized by a range of developmental and neurological abnormalities including epilepsy.^43^ The NSL complex genes play an important role in regulating core transcriptional and signaling networks,^44^ mutations in which have been associated with NDDs.^45,46^ The *KDM4B* gene encodes a demethylase enzyme that regulates gene expression essential for brain development,^47^ and rare variants in KDM4B have been implicated in global developmental delay.^48^ Collectively, these genes have an already established role in NDDs that present shared clinical characteristics with DEEs. Another new candidate, the PDE gene family, was found associated with NAFE. PDE enzymes catalyze the hydrolysis of cAMP and cGMP, two key second messengers modulating a variety of neuronal pathways.^49,50^ Lastly, a noteworthy finding was the *RYR2* gene associated with GGE, which was identified by combining rare and common genetic variation, representing an example of convergent epilepsy generic risk across the allele frequency spectrum.

Besides nominating new genes, identifying new variants in known epilepsy genes will also facilitate the characterization of specific mechanisms. Over the past five-year efforts from Epi25 WES, there has been a steady increase in the number of deleterious URVs discovered in epilepsy panel genes (Extended Data Fig. 6a); almost all (130/134) genes with a known monogenic cause have been identified with at least one deleterious URVs (Extended Data Fig. 6b), providing a valuable resource for downstream functional analysis. While the number of damaging missense URVs increases at a higher rate than protein-truncating URVs, the number of additional genes identified with a missense URV grows more slowly (Extended Data Fig. 6b). This pattern reflects an accumulation of missense URVs in the same set of genes, highlighting the need of effective approaches to characterize the specific functional consequences of these missense variants.

Compared to our prior URV results,^10^ the top genes that maintained or obtained stronger association in this enlarged study are known epilepsy genes (Extended Data Fig. 6c). This trend demonstrates a high replicability of existing gene findings, and likewise, calls for larger sample sizes to confirm the present results. Moreover, since our last Epi25 study, we began to include diverse genetic ancestry samples in our primary analysis (Extended Data Fig. 6d), and in the present study, we performed the first genetic ancestry-specific subgroup analyses (Extended Data Fig. 4). There was an overall trend of shared rare variant risk across ancestry groups, yet delineating differences in individual genes still awaits larger sample sizes. The power of identifying significant genes depends on both our ability to detect ultra-rare variants and the effect sizes of these variants in increasing epilepsy risk. At the current sample size, we have ∼80% power to detect strong gene burden with a large effect size of OR>8 (Extended Data Fig. 6e; Methods). Larger sample sizes are required for detecting smaller effect sizes. With the current case:control ratio, we will need ∼58K cases to achieve 80% power for detecting moderate gene burden (OR ≥3) and ∼462K cases for weak gene burden with an OR=1.5. Meanwhile, increasing the control-to-case ratio will also enhance power. With the same number of cases, doubling the controls will yield up to a 20% power increase and quadrupling the controls will provide us with nearly 100% power to detect strong gene burden (Extended Data Fig. 6f; Methods). Existing epilepsy panel genes are largely dominated by DEE genes, and these genes exhibited substantially stronger burden for URVs from DEEs than the other two epilepsy subtypes (Extended Data Fig. 6g). Excluding epilepsy panel genes, we found significant residual burden of URVs in genes that are intolerant to genetic variation (Extended Data Fig. 6g). Collectively, these findings suggest that the discovery of rare variant risk for epilepsy is far from saturation.

A promising strategy to accelerate gene discovery is to integrate URVs and other genetic variants. As presented in this study, the timeliness of our WES and GWAS efforts have resulted in the discovery of a wide allelic spectrum of epilepsy genetic risk factors, comprising different types of genetic variation, varying in size and/or frequency, each contributing to uncovering part of the complex genetic architecture of epilepsy. Currently missing pieces include balanced structural variants (e.g., inversions and translocations not detected in our CNV calls) and mitochondrial variants. Both have been recognized as source of epilepsy genetic risk^51,52^, while larger-scale studies are required for characterizing genome-wide patterns of these variants. Given the growing evidence that different genetic risk factors converge at least partially in the same genes, we believe that an extended model that jointly analyzes these variants would likely provide the most informative results beyond any single approach. Overall, the ongoing sequencing and genotyping efforts, together with the ever-increasing scale of genetic association studies, will continue to expand and/or refine our understanding of the genetic architecture of epilepsy, delineate specific underlying pathophysiological processes, and hopefully enable a move towards more targeted treatment approaches.

## Data Availability

We provide summary-level data at the variant and gene level in an online browser for visualization and download (https://epi25.broadinstitute.org/). There are no restrictions on the aggregated data released on the browser. Full results from the exome-wide burden analysis are also available in Supplementary Datasets 1 and 4. WES data from Epi25 cohorts are available via the NHGRI controlled-access AnVIL platform (https://anvilproject.org/; dbGaP accession phs001489). Data availability of non-Epi25 control cohorts is provided in the supplementary materials.

## Acknowledgements

We thank the Epi25 principal investigators, local staff overseeing individual cohorts, and all of the individuals with epilepsy and their families who participated in Epi25 for their commitment to this international collaboration. This work is part of the Centers for Common Disease Genomics (CCDG) program, funded by the National Human Genome Research Institute (NHGRI) and the National Heart, Lung, and Blood Institute (NHLBI). CCDG-funded Epi25 research activities at the Broad Institute, including genomic data generation in the Broad Genomics Platform, were supported by NHGRI grant UM1 HG008895 (PIs: E.S.L, S.B.G, M.J.D, and S.K.). The Genome Sequencing Program efforts were also supported by NHGRI grant 5U01HG009088. A supplemental grant for Epi25 phenotyping was supported by “Epi25 Clinical Phenotyping R03,” National Institutes of Health R03NS108145 (PIs: D.H.L. and S.F.B.). Additional support for analysis was provided by NINDS grant R01NS106104 (PI: C.C.). The content is solely the responsibility of the authors and does not necessarily represent the official views of the National Institutes of Health. We also thank the Stanley Center for Psychiatric Research at the Broad Institute for supporting the genomic data generation. Additional funding sources and acknowledgment of individual cohorts are listed in the supplemental materials.

## Author Contributions

All authors contributed to patient phenotyping data and sample collection or to analyses. Roles in specific committees of the project are listed in the supplementary materials.

---

**Extended Data Fig. 1:**
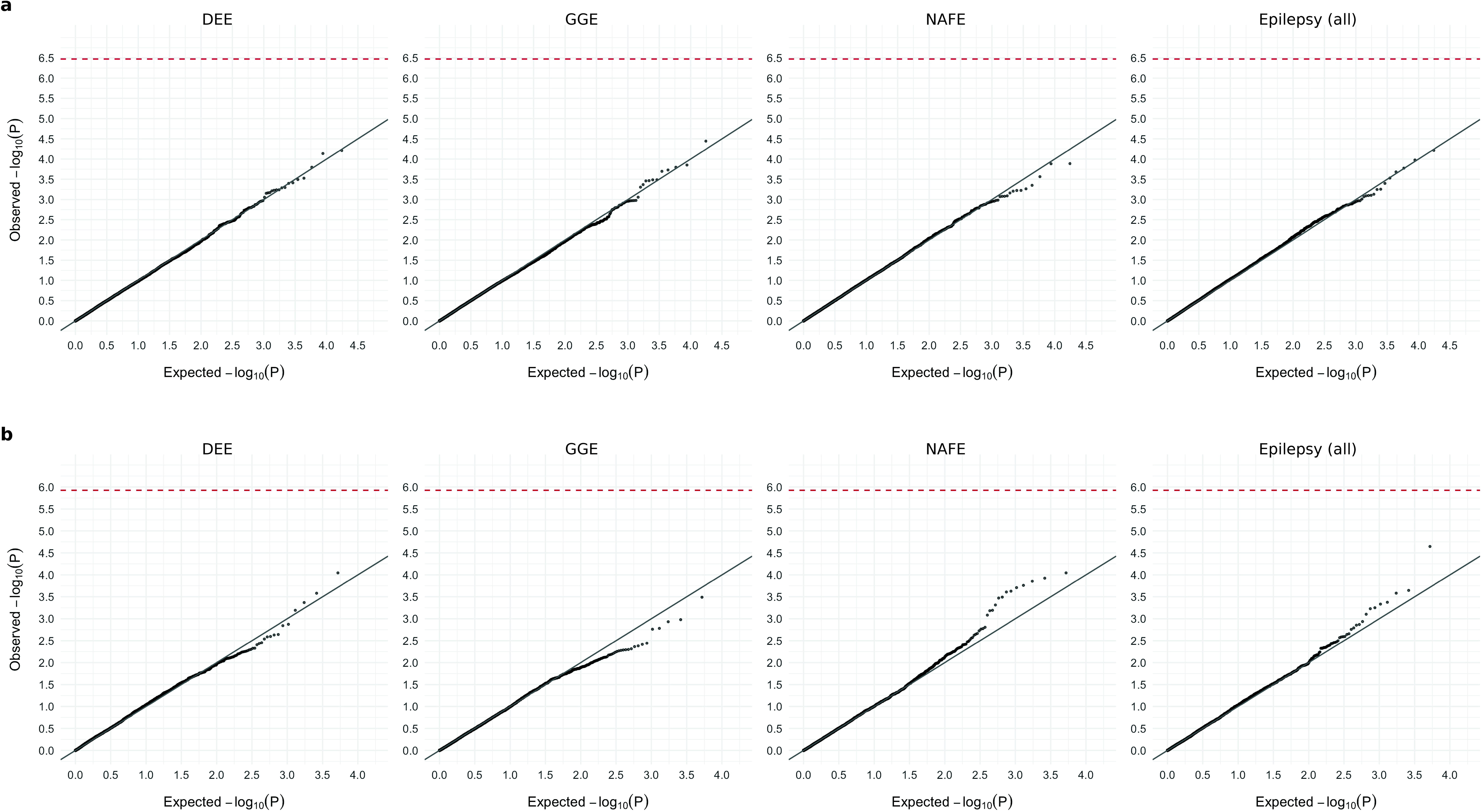
Results from burden analysis of synonymous URVs. **a**,**b**, Burden of synonymous URVs at the individual-gene (**a**) and the gene-set (**b**) level. The observed −log_10_-transformed *P* values are plotted against the expectation given a uniform distribution. Burden analyses are performed across four epilepsy groups – 1,938 DEEs, 5,499 GGE, 9,219 NAFE, and 20,979 epilepsy-affected individuals combined – versus 33,444 controls. *P* values are computed using a Firth logistic regression model testing the association between the case-control status and the number of URVs (two-sided); the red dashed line indicates exome-wide significance *P*=3.4×10^-7^ after Bonferroni correction (see Methods).

**Extended Data Fig. 2:**
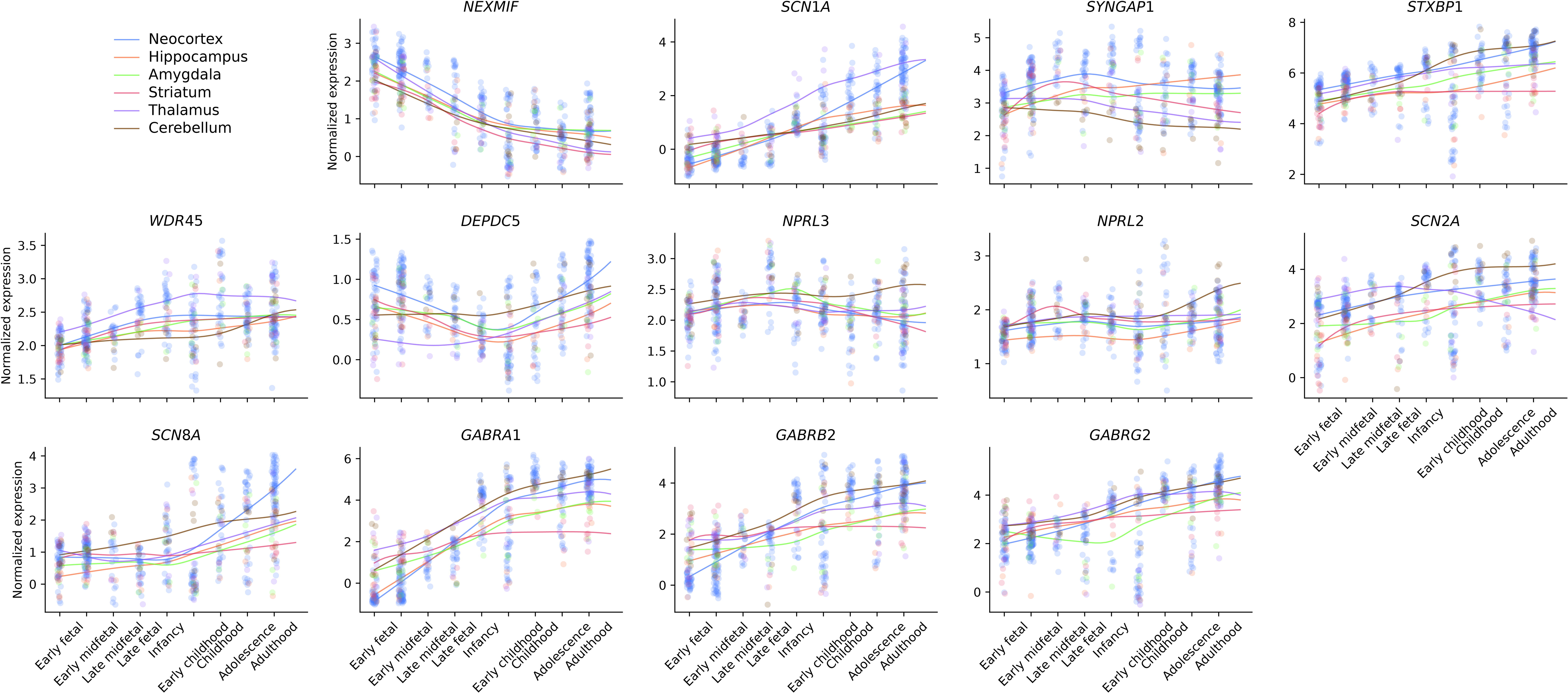
Spatiotemporal expression of 13 exome-wide significant genes in the human brain. Expression values (log_2_[TPM+1]) are normalized to the mean for each BrainSpan sample; each dot represents the expression value of a particular gene in a sample collected in a particular brain region and developmental time (from early fetal to adulthood: N=47/5/5/9/5/4, 69/6/6/7/5/4, 19/2/1/2/1/2, 27/2/2/2/2/3, 30/2/3/2/3/3, 41/3/4/3/4/5, 30/3/3/1/1/3, 36/3/3/2/2/4, and 63/6/6/6/6/6 neocortex/hippocampus/amygdala/striatum/thalamus/cerebellum samples, respectively). LOESS smooth curves are plotted for each brain region across developmental time.

**Extended Data Fig. 3:**
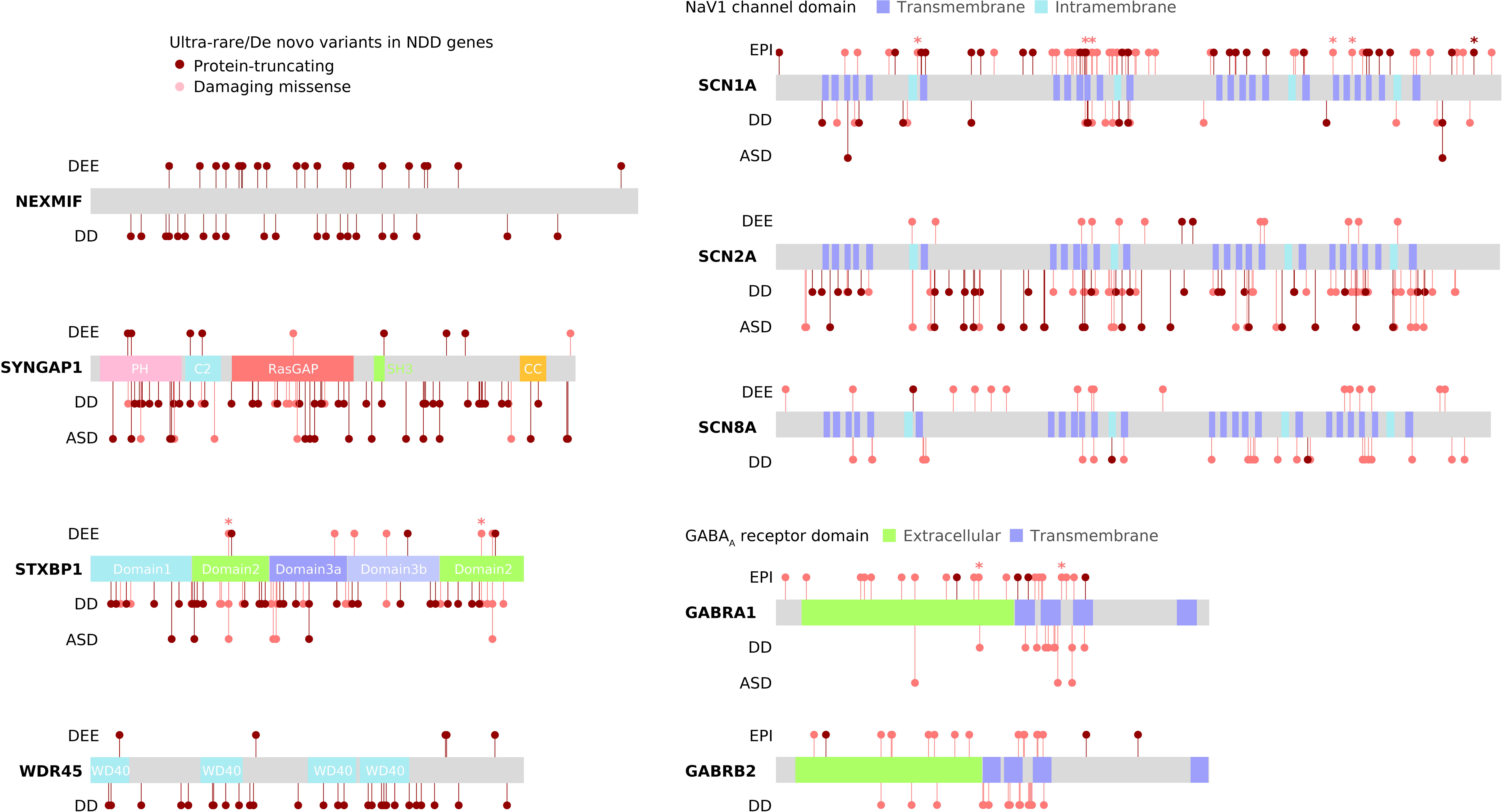
Distributions of URVs from this study and de novo variants from other NDD studies on the same genes. Schematic protein plots of nine genes that are significant in both our epilepsy cohort (DEE: developmental and epileptic encephalopathy; EPI: all-epilepsy combined) and previous large-scale WES studies of severe developmental disorders (DD) and/or autism spectrum disorder (ASD) are shown. Asterisk indicates recurring URVs in epilepsy; recurring de novo variants in DD/ASD as well as detailed variant information are provided in Supplementary Data 13.

**Extended Data Fig. 4:**
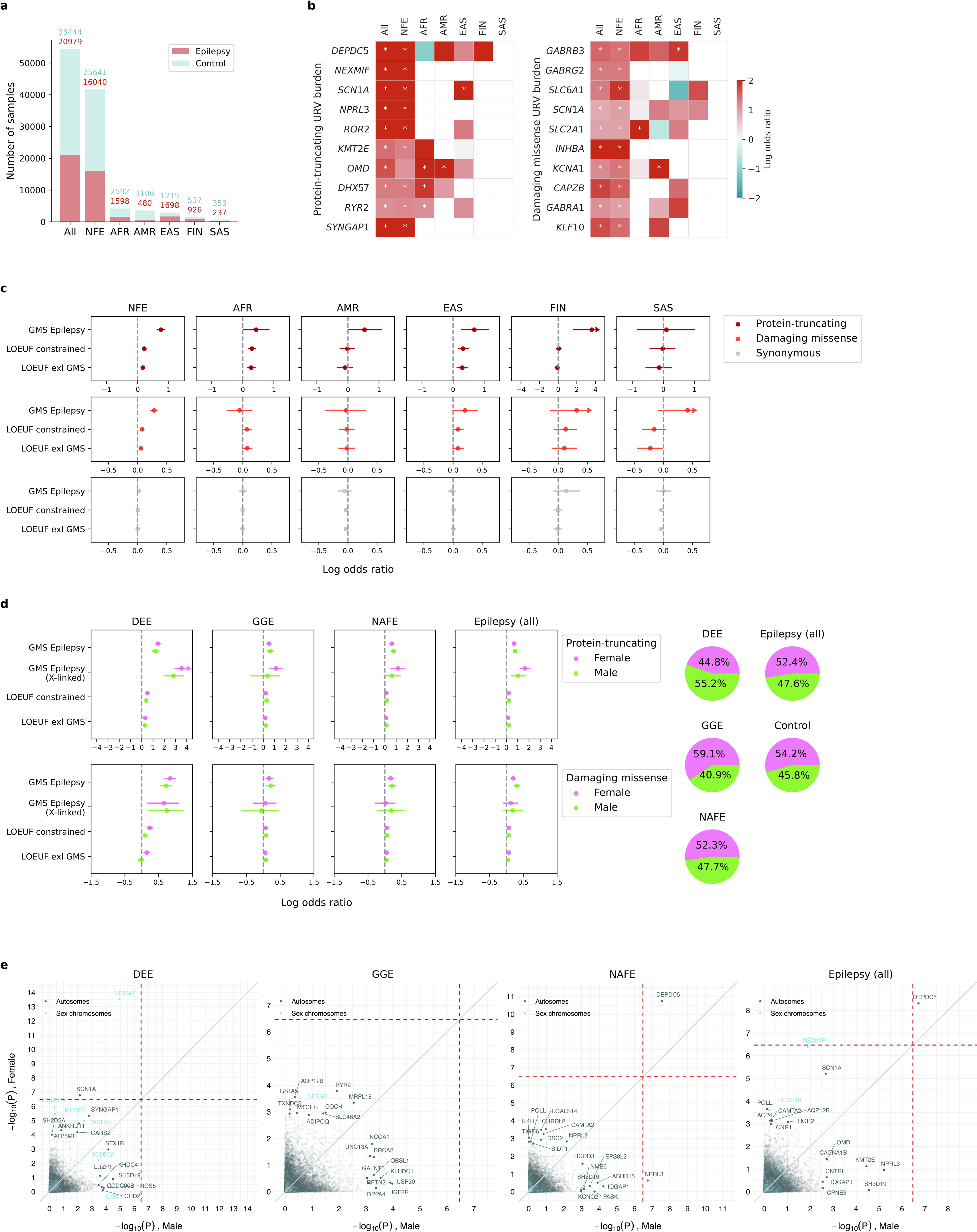
Results from genetic ancestry- and sex-specific burden analyses. **a**, The numbers of epilepsy cases (orange) and controls (blue) by genetic ancestry. **b**, Comparison of protein-truncating (left) and damaging missense (right) URV burden in the top ten genes from the primary analysis (“All”) across genetic ancestry subgroups. Red color indicates enrichment in cases (log[OR]>1), with an asterisk indicating nominal significance (*P*≤0.05; see Supplementary Data 14 for exact *P* values). *P* values are computed using a Firth logistic regression model testing the association between the case-control status and the number of URVs (two-sided). **c**, Genetic ancestry-specific burden of URVs in established epilepsy genes (N=171 curated by the Genetic Epilepsy Syndromes [GMS] panel with a known monogenic/X-linked cause), constrained genes (N=1,917 scored by the loss-of-function observed/expected upper bound fraction [LOEUF] metric as the most constrained 10% genes), and constrained genes excluding established epilepsy genes (N=1,813). Overall, different ancestral groups show at least partially shared burden of deleterious URVs in these gene sets. In **a**-**c**, NFE: Non-Finnish European (N_case_=16,040, N_control_=25,641), AFR: African (N_case_=1,598, N_control_=2,592), AMR: Ad Mixed American (N_case_=480, N_control_=3,106), EAS: East Asian (N_case_=1,698, N_control_=1,215), FIN: Finnish (N_case_=926, N_control_=537), SAS: South Asian (N_case_=237, N_control_=353). **d**, Sex-specific burden of URVs in established epilepsy genes. Burden analyses are performed for three gene sets described in c, with an additional set of 37 X-linked GMS epilepsy genes, across four epilepsy groups (female: N_DEE_=811, N_GGE_=4,807, N_NAFE_=3,511, N_EPI(all)_=11,372, N_control_=18,144; male: N_DEE_=997, N_GGE_=2,579, N_NAFE_=4,395, N_EPI(all)_=10,397, N_control_=15,302). There is an overall trend of shared URV burden between female and male subgroups in these gene sets. In **c** and **d**, the dot represents the log odds ratio and the error bars represent the 95% confidence intervals of the point estimates. For presentation purposes, error bars that exceed a large log odds ratio value are capped, indicated by arrows at the end of the error bars (see Supplementary Data 14 and 15 for exact values). **e**, Comparison of sex-specific burden of protein-truncating URVs at level of the individual genes. For each gene, the −log_10_-transformed *P* value from the female subgroup analysis (y-axis) is plotted against that from the male subgroup analysis (x-axis). Top ten genes with URV burden in epilepsy are labeled for each subgroup, with genes on the sex chromosomes colored in blue. The red dashed line indicates exome-wide significance *P*=3.4×10^-7^ after Bonferroni correction.

**Extended Data Fig. 5:**
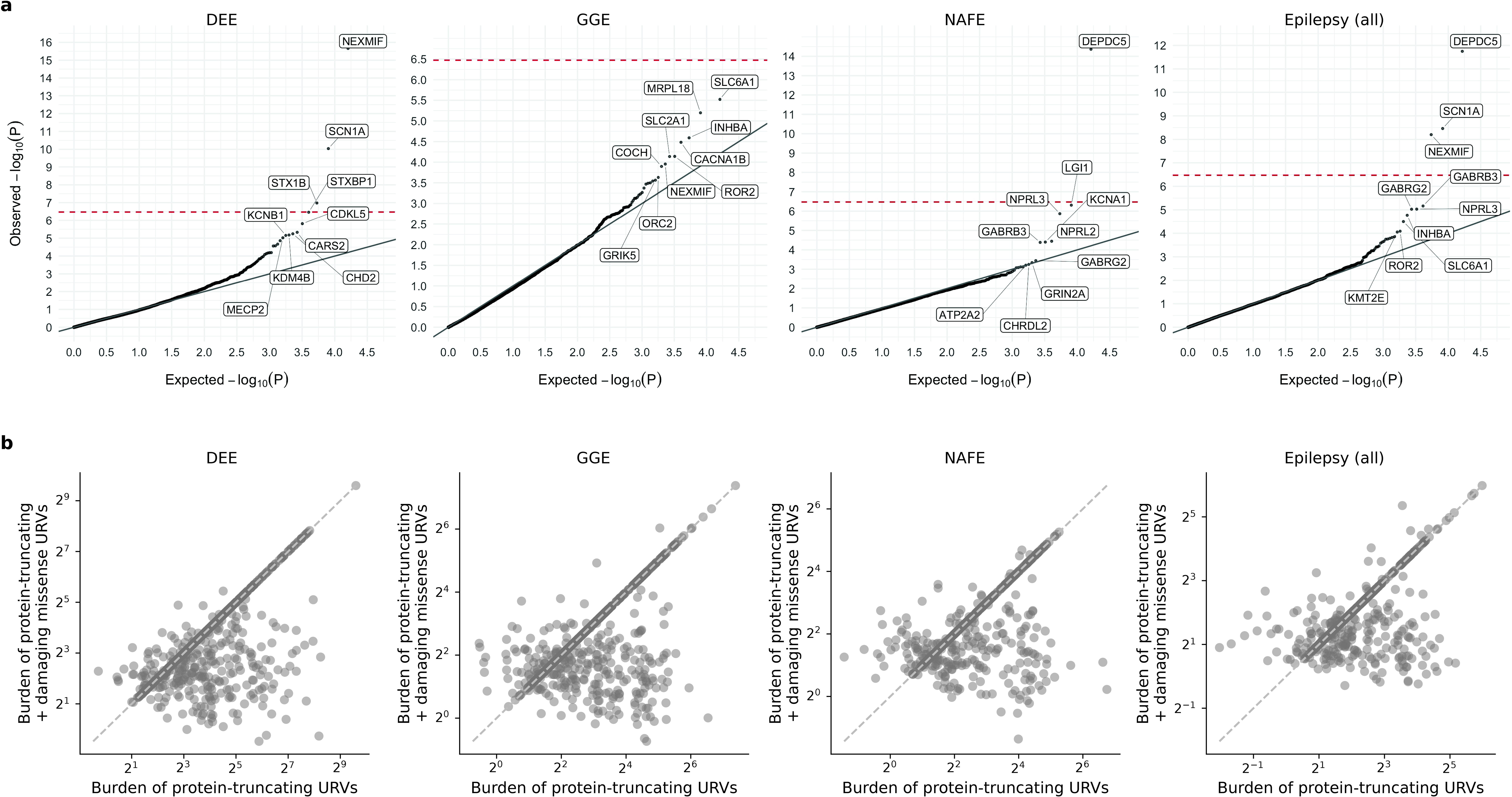
Results from burden analysis of protein-truncating and damaging missense URVs combined. **a**, Joint burden of protein-truncating and damaging missense URVs at the individual-gene level. The observed −log10-transformed *P* values are plotted against the expectation given a uniform distribution. Burden analyses are performed across four epilepsy groups – 1,938 DEEs, 5,499 GGE, 9,219 NAFE, and 20,979 epilepsy-affected individuals combined – versus 33,444 controls. *P* values are computed using a Firth logistic regression model testing the association between the case-control status and the number of URVs (two-sided); the red dashed line indicates exome-wide significance *P*=3.4×10^-7^ after Bonferroni correction (see Methods). **b**, Comparison of the joint burden in **a** with the burden of protein- truncating URVs. The odds ratio (OR) of protein-truncating plus damaging missense URVs (y-axis) and that of protein-truncating URVs alone (x-axis) are compared. Each dot represents a gene with significant enrichment (OR>0 and *P*≤0.05) of either protein-truncating URVs or the two variant classes combined.

**Extended Data Fig. 6:**
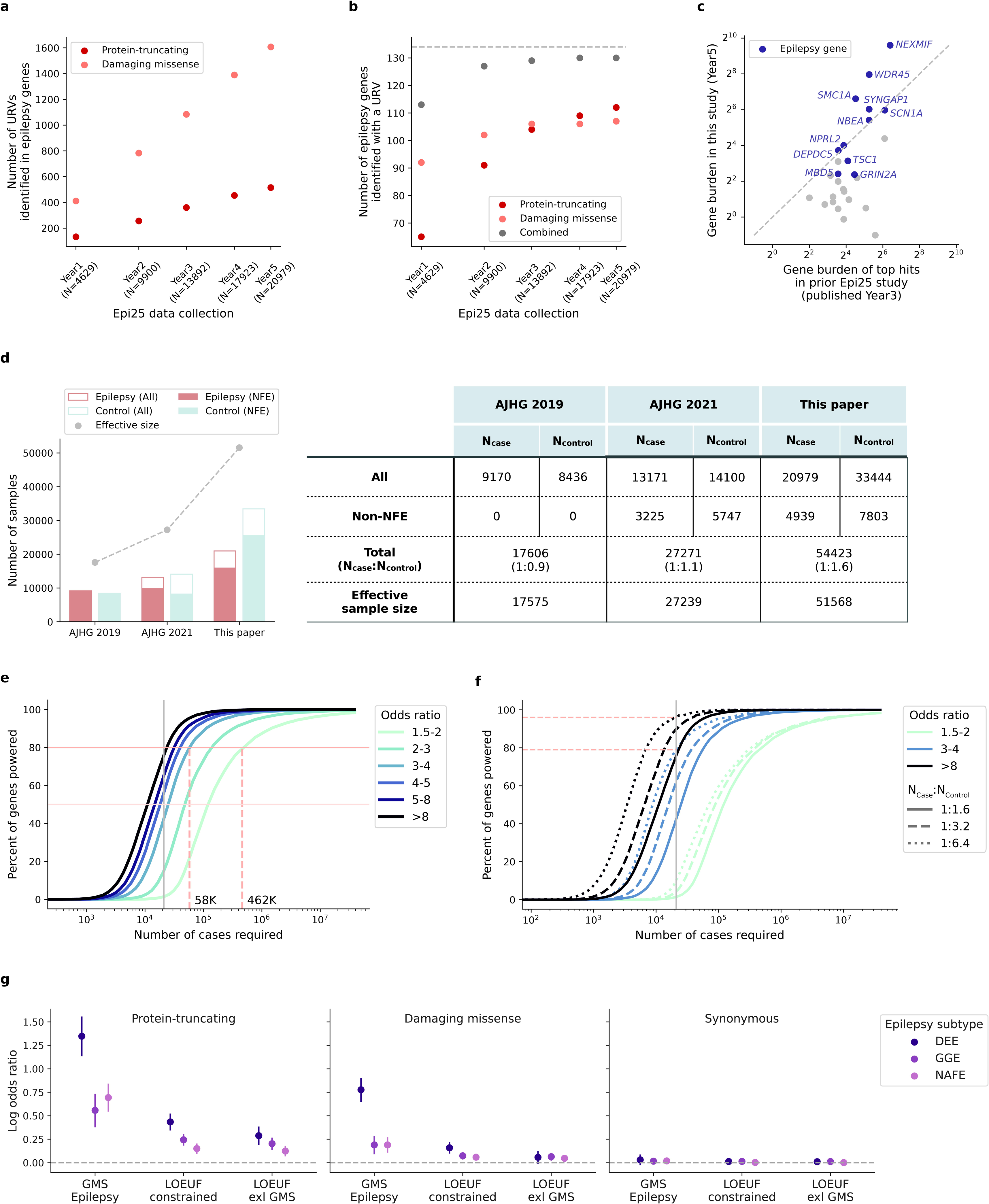
URV discovery and burden results across Epi25 data collection. **a**, Increase in the number of protein-truncating and damaging missense URVs discovered in epilepsy genes with a known monogenic cause. **b**, Increase in the number of monogenic epilepsy genes identified with a protein- truncating or damaging missense URV. In **a** and **b**, variant/gene count is plotted against the year of Epi25 data collection; the total number of epilepsy cases analyzed in each year is indicated in parenthesis. **c**, URV burden of previously top-ranked genes in this study. The odds ratio of protein-truncating URVs in genes from this study (y-axis) and the prior Epi25 publication (x-axis) are compared. Each dot represents one of the top ten genes implicated by our previous burden analysis (across three epilepsy subtypes). Genes with a known monogenic/X-linked cause are labeled and colored in purple. **d**, Increase in the total, non-European ancestry, and effective sample size in this study over our previous publications. The effective sample size is computed as 4/(1/N_case_+1/N_control_). **e**,**f**, The sample size required for well-powered gene burden testing. The percentage of genes powered to detect significant URV burden (Fisher’s exact *P* ≤0.05) at different effect sizes (**e**) and case:control ratios (**f**) is shown as a function of log-scaled sample size of epilepsy cases. Lighter color indicates smaller effect size (weaker burden), which requires a larger sample size to detect. The gray vertical line indicates the current sample size of 20,979 cases. In **e**, horizontal lines indicate 80% and 50% detection power, and vertical dashed lines indicate the estimated number of cases required to achieve 80% at the benchmarked effective sizes. In **f**, dashed and dotted curves indicate power estimation with increased control:case ratios from 1.6 (in this study) to 3.2 and 6.4, respectively; horizontal lines indicate the estimated power achieved by doubling and quadrupling the number of controls at the current sample size of cases. **g**, Epilepsy subtype-specific burden of URVs in established epilepsy genes (N=171 curated by the Genetic Epilepsy Syndromes [GMS] panel with a known monogenic/X-linked cause), constrained genes (N=1,917 scored by the loss-of-function observed/expected upper bound fraction [LOEUF] metric as the most constrained 10% genes), and constrained genes excluding established epilepsy genes (N=1,813). Burden analyses are performed across three epilepsy subtypes – 1,938 DEEs, 5,499 GGE, and 9,219 NAFE – versus 33,444 controls. Protein- truncating and damaging missense URVs from DEEs exhibit the strongest enrichment in epilepsy panel genes, while all epilepsy subtypes show significant enrichment in constrained genes even after excluding the panel genes. No enrichment is observed for synonymous URVs. The dot represents the log odds ratio and the error bars represent the 95% confidence intervals of the point estimates.

## Methods

### Study design and participants

We collected DNA and detailed phenotyping data of individuals with epilepsy from 59 participating Epi25 sites in Europe, North America, Australasia, and Asia (Supplementary Table 1). In total, we analyzed 20,979 epilepsy cases – including 1,938, 5,499, and 9,219 individuals with DDEs, GGE, and NAFE, respectively, and 4,323 with other epilepsies (mostly lesional focal epilepsy [2,495] and febrile seizures [FS]/FS+ [327]) – and 33,444 controls. Control individuals were aggregated from a subset of Epi25 sites, local collections at the Broad Institute, or dbGaP and were not screened for neurological or neuropsychiatric conditions (see Supplementary Table 2).

### Phenotyping procedures

Epilepsies were diagnosed by epileptologists on clinical grounds (see below for specific criteria for DEEs, GGE, and NAFE) in accordance with the International League Against Epilepsy (ILAE) classification at the time of diagnosis and recruitment.^1,9,10^ Phenotyping data were entered into the Epi25 Data repository (https://github.com/Epi25/epi25-edc) via case record forms hosted on the REDCap platform^53^. The data fields do not contain protected health information (PHI). Data collected from previous coordinated efforts with phenotyping on databases (e.g., the Epilepsy Phenome/Genome Project^54^ and the EpiPGX project http://www.epipgx.eu) were integrated via scripted transformations. All phenotyping data underwent review for uniformity among sites and quality control (QC) by automated data checking and manual review as required; the process was overseen by a phenotyping committee with clinical expertise.

### Epilepsy case definitions

Epilepsy diagnoses and classification for Epi25 have been described previously.^9,10^ In brief, diagnosis of DEEs required severe refractory epilepsy of unknown etiology, with developmental plateau or regression, and with epileptiform features on electroencephalogram (EEG). Diagnosis of GGE required a history of generalized seizure types (generalized tonic-clonic, absence, or myoclonic seizures) with generalized epileptiform discharges on EEG; exclusion criteria included focal seizures, moderate-to-severe intellectual disability, and epileptogenic lesions on neuroimaging if available. Diagnosis of NAFE required a history of focal seizures with either focal epileptiform discharges or normal findings on EEG; exclusion criteria included primary generalized seizures, moderate-to-severe intellectual disability, and neuroimaging lesions (except hippocampal sclerosis).

### Informed consent

Adult participants, or the legal guardian of child participants, provided signed informed consent at participating centers based on the local ethical requirements at the time of collection. The consent was required not to exclude data sharing to be included in the study. Consent forms for samples collected after January 25, 2015 required specific language according to the National Institutes of Health’s Genomic Data Sharing Policy. Before sequencing at the Broad Institute of MIT and Harvard, the Mass General Brigham (MGB) Institutional Review Board (IRB) provided approval for secondary use of each of the Epi25 cohorts, after reviewing the consent forms from each cohort (MGB protocol number: 2012P000788). The Broad’s Office of Research Subjects Protection (ORSP) provided approval for each cohort (ORSP approval number: ORSP-1733). For deposition of data into the AnVIL, we obtained Data Use Limitations letters for each cohort from the original IRBs and Institutional Certifications from the Broad’s ORSP.

### Whole-exome sequencing data generation

All samples were sequenced at the Broad Institute of MIT and Harvard on the Illumina HiSeq X or NovaSeq 6000 platforms with 150 bp paired-end reads. Exome capture was performed using multiple kits: the Illumina Nextera Rapid Capture Exomes or TruSeq Rapid Exome enrichment kit (target size 38 Mb) and the Twist Custom Capture (target size 37 Mb). Sequence data in the form of BAM files were generated via the Picard data-processing pipeline and well-calibrated reads were aligned to the human reference GRCh38. Variants were jointly called across all samples via the Genome Analysis Toolkit (GATK) best-practice pipeline^55^ and were annotated using Variant Effect Predictor (VEP)^56^ with custom annotations, including LOFTEE (Loss-Of-Function Transcript Effect Estimator)^57^ and MPC (missense badness, PolyPhen-2, and regional constraint),^11^ using Hail.^58^

### Variant and sample QC

Initial variant QC criteria included: (1) genotype quality (GQ) ≥20, (2) read depth (DP) ≥20, (3) allele balance (AB) ≥0.2 and ≤0.8, (4) passing the GATK Variant Quality Score Recalibration (VQSR) filter, (5) residing in GENCODE coding regions that were well-covered by both capture platforms, where 80% of the Illumina or Twist sequenced samples had at least 10x coverage, and (6) outside of the low-complexity (LCR) regions.^59^ Additional variant QC were applied after sample QC (see below for details): (1) call rate ≥0.98, (2) case-control call rate difference ≤0.02, and (3) Hardy-Weinberg Equilibrium (HWE) test p value ≥10^-6^.

Sample QC criteria, on the basis of all sequenced samples and the initial QC-ed variants, included: (1) mean call rate ≥0.90, (2) mean GQ ≥57, (3) mean DP ≥25, (4) freemix contamination estimate ≤2.5%, (5) percent chimeric reads ≤2%, and (6) the genetically imputed sex matching with self-reported sex. We performed principal component analysis (PCA) to classify samples into genetic ancestral groups, using a random forest model trained on the 1000 Genomes data; samples with a probability ≥0.9 to be one of the six populations – Non-Finnish European (NFE), Finnish (FIN), African (AFR), East Asian (EAS), South Asian (SAS), Ad Mixed American (AMR) – were retained. Within each ancestral group, we examined cryptic relatedness based on identity-by-descent (IBD) estimates and excluded one sample from each pair of related individuals with an IBD>0.2. Additional sample QC were applied on a population- and cohort-specific basis, which excluded outliers with >4 standard deviations from the mean of (1) transition/transversion ratio, (2) heterozygous/homozygous ratio, and (3) insertion/deletion ratio. To control for residual population stratification, we further excluded samples and/or cohorts that show extreme counts of synonymous singletons. The number of samples passed QC at each step is detailed in Supplementary Table 3.

### Exome-wide burden analysis

To evaluate the excess of rare, deleterious protein-coding variants in individuals with epilepsy, we performed burden analysis across the entire exome, at both an individual-gene and a gene-set level. “Ultra-rare” variants (URVs) were defined as variants observed no more than five copies among the combined case-control cohort, which corresponded to a minor allele frequency (MAF) <0.005%. Deleterious variants were defined and categorized into two classes: (1) protein-truncating annotated by LOFTEE and (2) damaging missense with an MPC score ≥2. We tested the burden of each URV class by regressing the case-control status on the URVs aggregated across a target gene or gene set in an individual, using a Firth regression model adjusting for sex and ancestry (the PCA-predicted genetic ancestral group and the top ten PCs). We further included the exome-wide count of synonymous singletons as an additional covariate to better control for residual population stratification not captured by PCs.^9^

We performed the burden analyses for each of the three major epilepsy subtypes – DEEs, GGE, and NAFE – and for all epilepsy-affected individuals combined. At the individual-gene level, we tested all protein-coding genes with at least one epilepsy or control carrier (protein-truncating: N=15,083, 15,236, 15,398, and 15,903 for the analysis of DEEs, GGE, NAFE, and all-epilepsy combined, respectively; damaging missense: N=4,013, 4,057, 4,105, and 4,194; synonymous: N=17,460, 17,463, 17,465, and 17,472). At the gene-set level, we tested collections of gene entities that belong to the same gene family^16^ or encode a particular protein complex^17^ and have at least one epilepsy or control carrier (protein-truncating: N=5,080, 5,070, 5,091, and 5,126 for the analysis of DEEs, GGE, NAFE, and all-epilepsy combined, respectively; damaging missense: N=3,256, 3,279, 3,298, and 3,343; synonymous: N=5,209). Exome-wide significance was determined by Bonferroni correction accounting for 18,531 consensus coding sequence (CCDS) genes or 5,373 gene sets – across four epilepsy groups and two variant classes – at *P*=3.4×10^-7^ and *P*=1.2×10^-6^ for the gene- and gene-set-based burden analysis, respectively.

### Protein structural analysis

We applied a metric^21^ that assesses the change in Gibbs free energy (ΔΔG/ddG in abbreviation) of protein folding induced by a mutation to characterize missense URVs identified in ion channel genes. In total, we computed ddG for 1,782 missense URVs on 16 ion channel protein complexes with experimentally resolved three-dimensional structures available (Supplementary Data 7). A positive ddG value suggests a decrease in Gibbs free energy of protein unfolding, i.e., a destabilizing effect of the mutation on protein, and a negative ddG value suggests a stabilizing effect. In the relevant burden analysis, we used |ddG|≥1 kcal/mol to prioritize variants that are likely to cause a change in protein stability.

### Copy number variant (CNV) calling and burden analysis

To call CNVs from the raw exome data, GATK-gCNV^60^ was used. In brief, GATK-gCNV is a Bayesian CNV caller, which adjusts for biases (i.e. GC content) introduced through capture kits and sequencing, while simultaneously accounting for systematic and technical differences. The raw sequencing files were compressed into counts and used as input across the annotated exons, and a subsequent principal component analysis-based method was used on the observed read counts to differentiate capture kits. This was followed by a hybrid distance- and density-based clustering approach to curate batches of samples to process in parallel. After, the caller was iteratively run for each batch and metrics produced by the Bayesian model were used to account for positive predictive value and sensitivity. GATK-gCNV exome QC filters were previously benchmarked in 8,439 matching genome and exome samples, as described in^22^.

Samples where GATK-gCNV made more than 100 unfiltered calls or more than 10 filtered calls were considered outlier samples and were removed. This resulted in 48,767 samples (∼90% of initial) for the downstream burden analysis, which comprises 18,963 epilepsy cases (including 1,743 DEEs, 4,980 GGE, and 8,425 NAFE) and 29,804 controls. To mitigate false positives, we used previously benchmarked filtering thresholds, where CNVs had to span >4 callable exons and had a site frequency <0.1% and a quality score >200. In the gene-based burden analysis of CNVs, we considered CNVs to affect a gene if ≥ 10% of the non-redundant exon-basepairs overlapped with the deletion (N_gene_= 4,213, 4,417, 4,733, and 6,045 for the analysis of DEEs, GGE, NAFE, and all-epilepsy combined, respectively), or if ≥75% of the non-redundant exon-basepairs overlapped with the duplication (N_gene_= 7,064, 7,282, 7,564, and 8,793 for the analysis of DEEs, GGE, NAFE, and all-epilepsy combined, respectively). When evaluating the joint burden of CNV deletions and protein-truncating SNVs/indels, only the subset of samples passing CNV calling QC were considered.

### Brain transcriptome analysis

The spatiotemporal transcriptome of the human brain was obtained from BrainSpan^34^, which contains 524 bulk RNA-seq samples collected from 26 brain regions of 42 human subjects. Gene expression values were log-transformed (log_2_[TPM+1]) and centered to the mean expression level for each sample. Epilepsy candidate genes identified by this or our recent GWA study were selected for each sample, and their average centered expression values were calculated and compared by region and developmental time, using Wilcoxon signed rank test. Six brain regions and nine developmental periods were defined based on the oncology described in the original study: neocortex (including orbital prefrontal cortex [OFC], dorsolateral prefrontal cortex [DFC], ventrolateral prefrontal cortex [VFC], medial prefrontal cortex [MFC], primary motor M1 cortex [M1C], primary somatosensory S1 cortex [S1C], posterior inferior parietal cortex [IPC], primary auditory A1 cortex [A1C], superior temporal cortex [STC], inferior temporal cortex [ITC], primary visual V1 cortex [V1C]), hippocampus (HIP), amygdala (AMY), striatum (STR), thalamus (MD), cerebellum (CB); early fetal (8-12 post-conceptional weeks [PCWs]), early midfetal (13-18 PCWs), late midfetal (19-23 PCWs), late fetal (24-37 PCWs), infancy (4-11 months), early childhood (1-5 years), childhood (6-11 years), adolescence (12-19 years), Adulthood (≥20 years).

### Gene Ontology (GO) enrichment analysis

We surveyed functional enrichment for candidate epilepsy genes with a prenatal- or postnatal-expression pattern (N=43 and 50, respectively) via the GO enrichment analysis portal (http://geneontology.org/). Enrichment was evaluated by a Fisher’s exact test across 43,008 GO terms (2023-05 release)^36^ and significant terms with a False Discovery Rate (FDR) 0.05 were reported. We manually reviewed and classified the top significant terms into two main functional categories: (1) “gene transcription regulation”, which included 32/43 prenatal genes from GO:0006325 chromatin organization, GO:0006357 regulation of transcription by RNA polymerase II, GO:0003677 DNA binding, GO:0000166 nucleotide binding; (2) “neuronal transmission”, which included 34/50 postnatal genes from GO:0042391 regulation of membrane potential, GO:0007268 chemical synaptic transmission, GO:0034220 monoatomic ion transmembrane transport, GO:0050804 modulation of chemical synaptic transmission, GO:0050808 synapse organization, GO:0099504 synaptic vesicle cycle, GO:0007267 cell-cell signaling, GO:0005509 calcium ion binding, GO:0006536 glutamate metabolic process. Full results of GO enrichment analysis are provided in Supplementary Data 12.

### Gene regulatory network analysis

To examine whether genes with a prenatal expression preference and genes with a postnatal expression preference are connected through gene regulation, we constructed a TF-target network by systematically searching for regulatory targets of the prenatal genes, and evaluated whether the postnatal genes are overrepresented in these targets. We used the ChEA3^61^ database to obtain TF-target connections (determined by ChIP-Seq experiments and manually curated individual TF studies) and co-expression of TFs with other genes based on RNA-seq data in the human brain. In total, we found target data for 10 of 43 prenatal genes, which resulted in a network of 10,560 TF-target connections with 7,792 genes as nodes and directed edges as regulation relationships. Forty connections were found between 10 TF prenatal genes and 26 postnatal genes (Supplementary Data 12). To assess the significance of the observed connectivity, we performed 10,000 random draws of gene sets matching the postnatal genes with respect to postnatal expression levels and prenatal-postnatal expression preference. An empirical *P* value was computed as the proportion of random gene sets with greater or equal connectivity compared to the postnatal gene set.

### Power estimation

We estimated the power of gene burden testing as the percentage of genes to be detected with sufficient number of protein-truncating URVs to obtain a *P* value≤0.05 by Fisher’s exact test, and benchmarked the power by different effect sizes (OR ranges from 1.5 to 8) and case:control ratios (ranges from 1:1.6 [in this study] to 1:3.2 and 1:6.4). For a given effect size and case:control ratio, the minimum numbers of URVs in cases and controls were determined, and the number of samples required to discover corresponding total number of URVs was estimated using a linear model of log(number of URVs) ∼ log(number of samples) from downsampling the WES dataset in this study.

## Data Availability

We provide summary-level data at the variant and gene level in an online browser for visualization and download (https://epi25.broadinstitute.org/). There are no restrictions on the aggregated data released on the browser. Full results from the exome-wide burden analysis are also available in Supplementary Data 1 and 4. WES data from Epi25 cohorts are available via the NHGRI’s controlled-access AnVIL platform (https://anvilproject.org/; dbGaP accession phs001489). Data availability of non-Epi25 control cohorts is provided in Supplementary Information. Source data are provided with this paper.

Publicly available datasets analyzed in this study include

Gene family: https://zenodo.org/records/3582386

CORUM protein complexes: https://mips.helmholtz-muenchen.de/corum/

Protein Data Bank: https://www.rcsb.org/

(Structure analyzed in Fig.3c: https://www.rcsb.org/structure/6x3z)

BrainSpan: https://www.brainspan.org/

Gene Ontology: https://geneontology.org/

ChEA3: https://maayanlab.cloud/chea3/

## Supplementary Information

This file contains summaries of Sequence Data Collection and Quality Control (Supplementary Tables 1-3 and Supplementary Figures 1-5), full descriptions of Supplementary Data, Supplementary Subjects and Methods (including details of individual participating Epi25 cohorts), Supplementary Acknowledgments, and Supplementary References.

### Supplementary Data

This file contains Supplementary Data items 1-15; see Supplementary Information for full descriptions.

### Code Availability

No custom code was used in this study. For sequence data generation, we used GATK v3.4 and v3.6 (GATK nightly-2015-07-31-g3c929b0, 3.4-89-ge494930, and 3.6-0-g89b7209), Picard version 1.1431, and VerifyBamlD version 1.0.0. Sample and variant QC was performed using functions in Hail 0.1 and 0.2 (website: https://www.hail.is; documentation: https://hail.is/docs/0.1/ and https://hail.is/docs/0.2/; GitHub repository: https://github.com/hail-is/hail). Variant annotation was performed using the Ensembl Variant Effect Predictor (VEP) v85 tool as implemented in Hail 0.1 with the LOFTEE annotation provided as default (https://github.com/konradjk/loftee/tree/27b0040f524348baa7f3257flce58993529e09ef). For phenotyping data, case record forms were hosted on the REDCap platform version 14, and entered into the Epi25 Data repository (https://github.com/Epi25/epi25-edc). For gene burden analysis, we used the R (v3.6.1) package logistf version 1.26.0 (https://cran.r-project.org/web/packages/logistf/index.html) to implement the Firth regression model. Additional processing and visualization was performed using R functions in the tidyverse library version vl.3.0 (https://www.tidyverse.org/packages/).

## A full list of authors and affiliations

Siwei Chen^1-3^, Bassel W. Abou-Khalil^4^, Zaid Afawi^5^, Quratulain Zulfiqar Ali^6^, Elisabetta Amadori^7^, Alison Anderson^8, 9^, Joe Anderson^10^, Danielle M. Andrade^6^, Grazia Annesi^11^, Mutluay Arslan^12^, Pauls Auce^13^, Melanie Bahlo^14, 15^, Mark D. Baker^16^, Ganna Balagura^17^, Simona Balestrini^18, 19^, Eric Banks^20^, Carmen Barba^21^, Karen Barboza^6^, Fabrice Bartolomei^22^, Nick Bass^23^, Larry W. Baum^24^, Tobias H. Baumgartner^25^, Betül Baykan^26^, Nerses Bebek^27, 28^, Felicitas Becker^29, 30^, Caitlin A. Bennett^31^, Ahmad Beydoun^32^, Claudia Bianchini^21^, Francesca Bisulli^33, 34^, Douglas Blackwood^35^, Ilan Blatt^5, 36^, Ingo Borggräfe^37^, Christian Bosselmann^29^, Vera Braatz^18, 19^, Harrison Brand^2, 38, 39^, Knut Brockmann^40^, Russell J. Buono^41-43^, Robyn M. Busch^44-46^, S. Hande Caglayan^47^, Laura Canafoglia^48^, Christina Canavati^49^, Barbara Castellotti^50^, Gianpiero L. Cavalleri^51, 52^, Felecia Cerrato^1^, Francine Chassoux^53^, Christina Cherian^54^, Stacey S. Cherny^55^, Ching-Lung Cheung^56^, I-Jun Chou^57^, Seo-Kyung Chung^16, 58, 59^, Claire Churchhouse^1-3^, Valentina Ciullo^60, 61^, Peggy O. Clark^62^, Andrew J. Cole^63^, Mahgenn Cosico^41, 64^, Patrick Cossette^65^, Chris Cotsapas^66^, Caroline Cusick^1^, Mark J. Daly^1-3, 67^, Lea K. Davis^68-71^, Peter De Jonghe^72-74^, Norman Delanty^51, 52, 75^, Dieter Dennig^76^, Chantal Depondt^77^, Philippe Derambure^78^, Orrin Devinsky^79^, Lidia Di Vito^34^, Faith Dickerson^80^, Dennis J. Dlugos^41, 81^, Viola Doccini^21^, Colin P. Doherty^52, 82^, Hany El-Naggar^52, 75^, Colin A. Ellis^83^, Leon Epstein^84^, Meghan Evans^85^, Annika Faucon^86^, Yen-Chen Anne Feng^1, 2, 87-89^, Lisa Ferguson^45^, Thomas N. Ferraro^42, 90^, Izabela Ferreira Da Silva^91^, Lorenzo Ferri^33, 34^, Martha Feucht^92^, Madeline C. Fields^93^, Mark Fitzgerald^41, 64, 83^, Beata Fonferko-Shadrach^16^, Francesco Fortunato^94^, Silvana Franceschetti^95^, Jacqueline A. French^79^, Elena Freri^96^, Jack M. Fu^2, 38, 39^, Stacey Gabriel^2^, Monica Gagliardi^11^, Antonio Gambardella^94^, Laura Gauthier^20^, Tania Giangregorio^97^, Tommaso Gili^60, 98^, Tracy A. Glauser^62^, Ethan Goldberg^41, 64^, Alica Goldman^99^, David B. Goldstein^100^, Tiziana Granata^96^, Riley Grant^2^, David A. Greenberg^101^, Renzo Guerrini^21, 102^, Aslı Gundogdu-Eken^47^, Namrata Gupta^2^, Kevin Haas^4^, Hakon Hakonarson^41^, Garen Haryanyan^26^, Martin Häusler^103^, Manu Hegde^104^, Erin L. Heinzen^105^, Ingo Helbig^41, 64, 83, 106–108^, Christian Hengsbach^29^, Henrike Heyne^2, 109^, Shinichi Hirose^110^, Edouard Hirsch^111^, Chen-Jui Ho^112^, Olivia Hoeper^31^, Daniel P. Howrigan^1-3^, Donald Hucks^68, 71^, Po-Chen Hung^57^, Michele Iacomino^7^, Yushi Inoue^113^, Luciana Midori Inuzuka^114, 115^, Atsushi Ishii^116^, Lara Jehi^45, 46^, Michael R. Johnson^117^, Mandy Johnstone^35^, Reetta Kälviäinen^118, 119^, Moien Kanaan^49^, Bulent Kara^120^, Symon M. Kariuki^121-123^, Josua Kegele^29^, Yeşim Kesim^26^, Nathalie Khoueiry-Zgheib^124^, Jean Khoury^45, 46^, Chontelle King^85^, Karl Martin Klein^54, 125-130^, Gerhard Kluger^131, 132^, Susanne Knake^130, 133^, Fernando Kok^115, 134^, Amos D. Korczyn^5^, Rudolf Korinthenberg^135^, Andreas Koupparis^136^, Ioanna Kousiappa^136^, Roland Krause^91^, Martin Krenn^137^, Heinz Krestel^66, 138^, Ilona Krey^139^, Wolfram S. Kunz^25, 140^, Gerhard Kurlemann^141^, Ruben I. Kuzniecky^142^, Patrick Kwan^8, 9, 143^, Maite La Vega-Talbott^93^, Angelo Labate^144^, Austin Lacey^51, 52^, Dennis Lal^44, 45^, Petra Laššuthová^145^, Stephan Lauxmann^29^, Charlotte Lawthom^10, 16^, Stephanie L. Leech^31^, Anna-Elina Lehesjoki^146, 147^, Johannes R. Lemke^139^, Holger Lerche^29^, Gaetan Lesca^148^, Costin Leu^18, 44^, Naomi Lewin^41, 64^, David Lewis-Smith^41, 108, 149, 150^, Gloria Hoi-Yee Li^151^, Calwing Liao^1-3, 38^, Laura Licchetta^34^, Chih-Hsiang Lin^112^, Kuang-Lin Lin^57^, Tarja Linnankivi^152-154^, Warren Lo^155^, Daniel H. Lowenstein^104^, Chelsea Lowther^2, 38, 39^, Laura Lubbers^156^, Colin H.T. Lui^157^, Lucia Inês Macedo-Souza^158^, Rene Madeleyn^159^, Francesca Madia^7^, Stefania Magri^160^, Louis Maillard^161^, Lara Marcuse^93^, Paula Marques^6^, Anthony G. Marson^162^, Abigail G. Matthews^163^, Patrick May^91^, Thomas Mayer^164^, Wendy McArdle^165^, Steven M. McCarroll^1, 2, 166^, Patricia McGoldrick^93, 167^, Christopher M. McGraw^63^, Andrew McIntosh^35^, Andrew McQuillan^23^, Kimford J. Meador^168^, Davide Mei^21^, Véronique Michel^169^, John J. Millichap^170^, Raffaella Minardi^34^, Martino Montomoli^21^, Barbara Mostacci^34^, Lorenzo Muccioli^33^, Hiltrud Muhle^106^, Karen Müller-Schlüter^171^, Imad M. Najm^45, 46^, Wassim Nasreddine^32^, Samuel Neaves^165, 172^, Bernd A. Neubauer^173^, Charles R.J.C. Newton^121-123, 174^, Jeffrey L. Noebels^99^, Kate Northstone^165^, Sam Novod^20^, Terence J. O’Brien^8, 9^, Seth Owusu-Agyei^175, 176^, Çiğdem Özkara^177^, Aarno Palotie^1, 3, 63, 87, 178^, Savvas S. Papacostas^136^, Elena Parrini^21, 102^, Carlos Pato^179, 180^, Michele Pato^179, 180^, Manuela Pendziwiat^106, 107^, Page B. Pennell^181^, Slavé Petrovski^8, 182^, William O. Pickrell^16, 183^, Rebecca Pinsky^184^, Dalila Pinto^185, 186^, Tommaso Pippucci^97^, Fabrizio Piras^60^, Federica Piras^60^, Annapurna Poduri^184^, Federica Pondrelli^33^, Danielle Posthuma^187^, Robert H.W. Powell^16,183^, Michael Privitera^188^, Annika Rademacher^106^, Francesca Ragona^96^, Byron Ramirez-Hamouz^185, 186^, Sarah Rau^29^, Hillary R. Raynes^93^, Mark I. Rees^16, 59^, Brigid M. Regan^31^, Andreas Reif^189, 190^, Eva Reinthaler^137^, Sylvain Rheims^191, 192^, Susan M. Ring^165, 172^, Antonella Riva^7, 17^, Enrique Rojas^84^, Felix Rosenow^129, 130, 133^, Philippe Ryvlin^193^, Anni Saarela^118, 119^, Lynette G. Sadleir^85^, Barış Salman^28^, Andrea Salmon^54^, Vincenzo Salpietro^7^, Ilaria Sammarra^11^, Marcello Scala^7, 17^, Steven Schachter^194^, André Schaller^195^, Christoph J. Schankin^138, 196^, Ingrid E. Scheffer^31, 197, 198^, Natascha Schneider^18, 19^, Susanne Schubert-Bast^129, 130, 199^, Andreas Schulze-Bonhage^200^, Paolo Scudieri^7, 17^, Lucie Sedláčková^145^, Catherine Shain^184^, Pak C. Sham^24^, Beth R. Shiedley^184^, S. Anthony Siena^201^, Graeme J. Sills^202^, Sanjay M. Sisodiya^18, 19^, Jordan W. Smoller^203, 204^, Matthew Solomonson^2, 88^, Gianfranco Spalletta^60, 205^, Kathryn R. Sparks^84^, Michael R. Sperling^206^, Hannah Stamberger^72-74^, Bernhard J. Steinhoff^207^, Ulrich Stephani^106^, Katalin Štěrbová^145^, William C. Stewart^101^, Carlotta Stipa^34^, Pasquale Striano^7, 17^, Adam Strzelczyk^129, 130, 133^, Rainer Surges^25^, Toshimitsu Suzuki^208, 209^, Mariagrazia Talarico^11^, Michael E. Talkowski^2, 38, 39^, Randip S. Taneja^4^, George A. Tanteles^136^, Oskari Timonen^119^, Nicholas John Timpson^165, 172^, Paolo Tinuper^33, 34^, Marian Todaro^8, 9^, Pınar Topaloglu^27^, Meng-Han Tsai^112^, Birute Tumiene^210, 211^, Dilsad Turkdogan^212^, Sibel Uğur-İşeri^28^, Algirdas Utkus^210, 211^, Priya Vaidiswaran^41, 64^, Luc Valton^213^, Andreas van Baalen^106^, Maria Stella Vari^7^, Annalisa Vetro^21^, Markéta Vlčková^145^, Sophie von Brauchitsch^129, 130, 133^, Sarah von Spiczak^106, 214^, Ryan G. Wagner^215-217^, Nick Watts^2,88^, Yvonne G. Weber^29, 218^, Sarah Weckhuysen^72-74^, Peter Widdess-Walsh^51, 52, 75^, Samuel Wiebe^54, 125, 127, 219, 220^, Steven M. Wolf^93, 167^, Markus Wolff^221^, Stefan Wolking^29, 218^, Isaac Wong^2, 38^, Randi von Wrede^25^, David Wu^86^, Kazuhiro Yamakawa^208, 209^, Zuhal Yapıcı^27^, Uluc Yis^222^, Robert Yolken^223^, Emrah Yücesan^224^, Sara Zagaglia^18, 19^, Felix Zahnert^130, 133^, Federico Zara^7, 17^, Fritz Zimprich^137^, Milena Zizovic^91^, Gábor Zsurka^25, 140^, Benjamin M. Neale^1-3^, Samuel F. Berkovic^31^

1. Stanley Center for Psychiatric Research, Broad Institute of MIT and Harvard, Cambridge, MA, USA.
2. Program in Medical and Population Genetics, Broad Institute of MIT and Harvard, Cambridge, MA, USA.
3. Analytic and Translational Genetics Unit, Department of Medicine, Massachusetts General Hospital and Harvard Medical School, Boston, MA 02114, USA.
4. Department of Neurology, Vanderbilt University Medical Center, Nashville, TN, USA.
5. Tel-Aviv University Sackler Faculty of Medicine, Ramat Aviv 69978, Israel.
6. University Health Network, University of Toronto, Toronto, ON, Canada.
7. IRCCS Istituto Giannina Gaslini, Genova, Italy.
8. Department of Medicine, University of Melbourne, Royal Melbourne Hospital, Parkville 3050, Australia.
9. Department of Neuroscience, The School of Translational Medicine, Alfred Health, Monash University, Melbourne 3004, Australia.
10. Neurology Department, Aneurin Bevan University Health Board, Newport, Wales, UK.
11. Department of Medical and Surgical Sciences, Neuroscience Research Center, Magna Graecia University, Catanzaro, Italy.
12. Department of Child Neurology, Gülhane Education and Research Hospital, Health Sciences University, Ankara, Turkey.
13. St George’s University Hospital NHS Foundation Trust, London, UK.
14. Population Health and Immunity Division, The Walter and Eliza Hall Institute of Medical Research, Parkville 3052, Australia. .
15. Department of Biology, University of Melbourne, Parkville 3010, Australia. .
16. Swansea University Medical School, Swansea University, Swansea, Wales, UK.
17. Department of Neurosciences, Rehabilitation, Ophthalmology, Genetics, Maternal and Child Health, University of Genova, Genova, Italy.
18. Department of Clinical and Experimental Epilepsy, UCL Queen Square Institute of Neurology, London WC1N 3BG, UK.
19. Chalfont Centre for Epilepsy, Chalfont-St-Peter, Buckinghamshire SL9 0RJ, UK. .
20. Data Sciences Platform, Broad Institute of MIT and Harvard, Cambridge, MA, USA.
21. Neuroscience Department, Meyer Children’s Hospital IRCCS, Florence, Italy.
22. Clinical Neurophysiology and Epileptology Department, Timone Hospital, Marseille, France.
23. Division of Psychiatry, University College London
24. Department of Psychiatry, The University of Hong Kong, Pokulam, Hong Kong.
25. Department of Epileptology, University of Bonn Medical Centre, Bonn 53127, Germany.
26. Department of Neurology, Istanbul Faculty of Medicine, Istanbul University, Istanbul, Turkey.
27. Department of Child Neurology, Istanbul Faculty of Medicine, Istanbul University, Istanbul, Turkey.
28. Department of Genetics, Aziz Sancar Institute of Experimental Medicine, Istanbul University, Istanbul, Turkey.
29. Department of Neurology and Epileptology, Hertie Institute for Clinical Brain Research, University of Tübingen, Tübingen 72076, Germany.
30. Department of Neurology, University of Ulm, Ulm 89081, Germany.
31. Epilepsy Research Centre, University of Melbourne, Austin Health, Heidelberg 3084, Australia.
32. Department of Neurology, American University of Beirut Medical Center, Beirut, Lebanon.
33. Department of Biomedical and Neuromotor Sciences, University of Bologna, Bologna, Italy.
34. IRCCS Istituto delle Scienze Neurologiche di Bologna, (Reference Center for Rare and Complex Epilepsies - EpiCARE), Bologna, Italy.
35. Division of Psychiatry, Centre for Clinical Brain Sciences, University of Edinburgh, Edinburgh, UK.
36. Department of Neurology, Sheba Medical Center, Ramat Gan, Israel.
37. Department of Pediatric Neurology, Dr von Hauner Children’s Hospital, Ludwig Maximilians University, Munchen, Germany.
38. Center for Genomic Medicine, Massachusetts General Hospital, Boston, MA, USA.
39. Department of Neurology, Massachusetts General Hospital and Harvard Medical School, Boston, MA, USA.
40. Children’s Hospital, Dept. of Pediatric Neurology, University Medical Center Göttingen, Göttingen, Germany.
41. Division of Neurology, Children’s Hospital of Philadelphia, Philadelphia, 3401 Civic Center Blvd, Philadelphia, PA 19104, USA.
42. Department of Biomedical Sciences, Cooper Medical School of Rowan University Camden, NJ 08103, USA.
43. Department of Neurology, Thomas Jefferson University Hospital, Philadelphia, PA 19107, USA.
44. Genomic Medicine Institute, Lerner Research Institute, Cleveland Clinic, Cleveland, OH 44195, USA.
45. Cleveland Clinic Epilepsy Center, Neurological Institute, Cleveland Clinic, Cleveland, OH 44195, USA.
46. Department of Neurology, Neurological Institute, Cleveland Clinic, Cleveland, OH 44195, USA.
47. Department of Molecular Biology and Genetics, Bogaziçi University, Istanbul, Turkey.
48. Integrated Diagnostics for Epilepsy, Fondazione IRCCS Istituto Neurologico C. Besta, Milan, Italy.
49. Hereditary Research Lab, Bethlehem University, Bethlehem, Palestine.
50. Unit of Medical Genetics and Neurogenetics, Department of Diagnostic and Technology, Fondazione IRCCS Istituto Neurologico Carlo Besta Milano, Italy.
51. School of Pharmacy and Biomolecular Sciences, The Royal College of Surgeons in Ireland, Dublin, Ireland.
52. The FutureNeuro Research Centre, Dublin, Ireland.
53. Epilepsy Unit, Department of Neurosurgery, Centre Hospitalier Sainte-Anne, and University Paris Descartes, Paris, France.
54. Department of Clinical Neurosciences, Cumming School of Medicine, University of Calgary, Calgary, Alberta, Canada.
55. Department of Epidemiology and Preventive Medicine, School of Public Health, Sackler Faculty of Medicine, Tel Aviv University, Tel Aviv 6997801, Israel.
56. Department of Pharmacology and Pharmacy, The University of Hong Kong, Pokfulam, Hong Kong.
57. Department of Pediatric Neurology, Chang Gung Memorial Hospital, Taoyuan, Taiwan.
58. Kids Research, Children’s Hospital at Westmead Clinical School, Faculty of Medicine and Health, University of Sydney, Sydney, New South Wales, Australia.
59. Brain & Mind Centre, Faculty of Medicine & Health, University of Sydney, Sydney, New South Wales, Australia.
60. Neuropsychiatry Laboratory, IRCCS Santa Lucia Foundation, Rome, Italy.
61. Department of Neurosciences, Psychology, Drug Research and Child Health, University of Florence, Florence, Italy.
62. Cincinnati Children’s Hospital Medical Center, Cincinnati, Ohio, USA.
63. Neurology, Massachusetts General Hospital, Boston, MA, USA.
64. The Epilepsy NeuroGenetics Initiative (ENGIN), Children’s Hospital of Philadelphia, Philadelphia, 3401 Civic Center Blvd, Philadelphia, PA 19104, USA.
65. Department of Neurosciences, Université de Montréal, Montréal, CA 26758, Canada.
66. Yale School of Medicine, New Haven, CT 06510, USA.
67. Institute for Molecular Medicine Finland, FIMM, HiLIFE, University of Helsinki, Helsinki, Finland
68. Division of Genetic Medicine, Department of Medicine, Vanderbilt University Medical Center, Nashville, TN, USA.
69. Department of Psychiatry and Behavioral Sciences, Vanderbilt University Medical Center, Nashville, TN, USA.
70. Department of Biomedical Informatics, Vanderbilt University Medical Center, Nashville, TN, USA.
71. Vanderbilt Genetics Institute, Vanderbilt University Medical Center, Nashville, TN, USA.
72. Applied & Translational Neurogenomics Group, VIB Center for Molecular Neurology, VIB, Antwerp, Belgium.
73. Department of Neurology, Antwerp University Hospital, Edegem 2650, Belgium.
74. Translational Neurosciences, Faculty of Medicine and Health Science, University of Antwerp, Antwerp, Belgium.
75. Department of Neurology, Beaumont Hospital, Dublin D09 FT51, Ireland.
76. Private Neurological Practice, Stuttgart, Germany.
77. Department of Neurology, CUB Erasme Hospital, Hôpital Universitaire de Bruxelles (H.U.B.), Université Libre de Bruxelles (ULB), 1070 Brussels, Belgium.
78. Department of Clinical Neurophysiology, Lille University Medical Center, EA 1046, University of Lille.
79. Department of Neurology, New York University/Langone Health, New York NY, USA.
80. Sheppard Pratt, 6501 North Charles Street, Baltimore, Maryland, USA.
81. Perelman School of Medicine, University of Pennsylvania, Philadelphia, PA, USA.
82. Neurology Department, St. James’s Hospital, Dublin D03 VX82, Ireland.
83. Department of Neurology, University of Pennsylvania, Perelman School of Medicine, Philadelphia, PA, 19104 USA.
84. Division of Neurology, Ann & Robert H. Lurie Children’s Hospital of Chicago, Chicago, IL USA.
85. Department of Paediatrics and Child Health, University of Otago, Wellington, New Zealand.
86. Human Genetics Training Program, Vanderbilt University, Nashville, TN, USA.
87. Psychiatric & Neurodevelopmental Genetics Unit, Department of Psychiatry, Massachusetts General Hospital and Harvard Medical School, Boston, MA 02114, USA.
88. Analytic and Translational Genetics Unit, Department of Medicine, Massachusetts General Hospital, Boston, MA, USA.
89. Division of Biostatistics, Institute of Epidemiology and Preventive Medicine, College of Public Health, National Taiwan University, Taipei 100, Taiwan.
90. Department of Pharmacology and Psychiatry, University of Pennsylvania Perlman School of Medicine, Philadelphia, PA 19104, USA.
91. Luxembourg Centre for Systems Biomedicine, University of Luxembourg, Esch-sur-Alzette L-4362, Luxembourg.
92. Department of Pediatrics and Neonatology, Medical University of Vienna, Vienna 1090, Austria.
93. Department of Neurology, Icahn School of Medicine at Mount Sinai, New York, NY 10029, USA.
94. Institute of Neurology, Department of Medical and Surgical Sciences, University “Magna Graecia”, Catanzaro, Italy.
95. Neurophysiology, Fondazione IRCCS Istituto Neurologico Carlo Besta, Milan, Italy.
96. Department of Pediatric Neuroscience, Fondazione IRCCS Istituto Neurologico Carlo Besta, Milan, Italy.
97. IRCCS Azienda Ospedaliero-Universitaria di Bologna, Medical Genetics Unit, Bologna, Italy.
98. IMT School for Advanced Studies Lucca, Lucca, Italy.
99. Department of Neurology, Baylor College of Medicine.
100. Institute for Genomic Medicine, Columbia University Medical Center, New York, NY 10032, USA.
101. Department of Pediatrics, Nationwide Children’s Hospital, Columbia, Ohio, USA.
102. Department of NEUROFARBA, University of Florence, Florence, Italy.
103. Division of Neuropediatrics and Social Pediatrics, Department of Pediatrics, University Hospital, RWTH Aachen, Aachen, Germany.
104. Department of Neurology, University of California, San Francisco, CA 94143, USA.
105. Division of Pharmacotherapy and Experimental Therapeutics, Eshelman School of Pharmacy, University of North Carolina at Chapel Hill, Chapel Hill, NC, 27599, USA.
106. Department of Neuropediatrics, University Medical Center Schleswig-Holstein, Christian- Albrechts-University, Kiel, Germany.
107. Institute of Clinical Molecular Biology, Christian-Albrechts-University of Kiel, Kiel, Germany.
108. Department of Biomedical and Health Informatics (DBHi), Children’s Hospital of Philadelphia, Philadelphia, PA, 19104 USA.
109. Hasso Plattner Institute, Digital Engineering Faculty, University of Potsdam, Germany.
110. General Medical Research Center, School of Medicine, Fukuoka University, Japan.
111. Department of Neurology, University Hospital of Strasbourg, Strasbourg, France.
112. Department of Neurology, Kaohsiung Chang Gung Memorial Hospital, Kaohsiung, Taiwan.
113. National Epilepsy Center, Shizuoka Institute of Epilepsy and Neurological Disorder, Shizuoka, Japan.
114. Epilepsy Clinic, Hospital Sirio-Libanes, Sao Paulo, Brazil.
115. Department of Neurology, University of Sao Paulo School of Medicine, Brazil.
116. Department of Pediatrics, Fukuoka Sanno Hospital, Japan.
117. Division of Brain Sciences, Imperial College London, London SW7 2AZ, UK.
118. Kuopio Epilepsy Center, Neurocenter, Kuopio University Hospital, Kuopio 70210, Finland.
119. Institute of Clinical Medicine, University of Eastern Finland, Kuopio 70210, Finland.
120. Department of Child Neurology, Medical School, Kocaeli University, Kocaeli, Turkey.
121. Neuroscience Unit, KEMRI-Wellcome Trust Research Programme, Kilifi, Kenya.
122. Department of Public Health, Pwani University, Kilifi, Kenya.
123. Department of Psychiatry, University of Oxford, Oxford, UK.
124. Department of Pharmacology and Toxicology, American University of Beirut Faculty of Medicine, Beirut, Lebanon.
125. Department of Community Health Sciences, Cumming School of Medicine, University of Calgary, Calgary, Alberta, Canada.
126. Department of Medical Genetics, Cumming School of Medicine, University of Calgary, Calgary, Alberta, Canada.
127. Hotchkiss Brain Institute, Cumming School of Medicine, University of Calgary, Calgary, Alberta, Canada.
128. Alberta Children’s Hospital Research Institute, University of Calgary, Calgary, Alberta, Canada.
129. Epilepsy Center Frankfurt Rhine-Main, Center of Neurology and Neurosurgery, Goethe University Frankfurt, Frankfurt, Germany.
130. LOEWE Center for Personalized Translational Epilepsy Research (CePTER), Goethe University Frankfurt, Germany.
131. Neuropediatric Clinic and Clinic for Neurorehabilitation, Epilepsy Center for Children and Adolescents, Vogtareuth, Germany.
132. Research Institute Rehabilitation / Transition, / Palliation, PMU Salzburg, Austria.
133. Epilepsy Center Hessen-Marburg, Department of Neurology, Philipps University Marburg, Marburg, Germany.
134. Mendelics Genomic Analysis, São Paulo, Brazil.
135. Department of Neuropediatrics and Muscular Disorders, University Medical Center, University of Freiburg, Freiburg, Germany.
136. Cyprus Institute of Neurology and Genetics, Nicosia, Cyprus.
137. Department of Neurology, Medical University of Vienna, Vienna 1090, Austria.
138. Department of Neurology, Inselspital, Bern University Hospital, University of Bern, Bern 3010, Switzerland.
139. Institute of Human Genetics, University of Leipzig Medical Center, Leipzig, Germany.
140. Institute of Experimental Epileptology and Cognition Research, Medical Faculty, University of Bonn, Bonn, Germany.
141. Bonifatius Hospital Lingen, Neuropediatrics Wilhelmstrasse 13, 49808 Lingen, Germany.
142. Department of Neurology, Hofstra-Northwell Medical School, New York, NY, USA.
143. Department of Medicine and Therapeutics, Chinese University of Hong Kong, Hong Kong, China.
144. Neurophysiopatology and Movement Disorders Clinic, University of Messina, Messina, Italy.
145. Department of Paediatric Neurology, 2nd Faculty of Medicine, Charles University and Motol Hospital, Prague, Czech Republic.
146. Folkhälsan Research Center, Helsinki 00290, Finland.
147. Medicum, University of Helsinki, Helsinki 00290, Finland.
148. Department of Medical Genetics, Hospices Civils de Lyon and University of Lyon, Lyon, France.
149. Translational and Clinical Research Institute, Newcastle University, Newcastle Upon Tyne, UK.
150. Department of Clinical Neurosciences, Newcastle Upon Tyne Hospitals NHS Foundation Trust, Newcastle Upon Tyne, UK.
151. Department of Health Technology and Informatics, The Hong Kong Polytechnic University, Hung Hum, Hong Kong.
152. Child Neurology, New Childreńs Hospital, Helsinki, Finland.
153. Pediatric Research Center, University of Helsinki, Helsinki, Finland.
154. Helsinki University Hospital, Helsinki, Finland.
155. Department of Pediatrics and Neurology, Nationwide Children’s Hospital, Columbus, OH, USA.
156. Citizens United for Research in Epilepsy, Chicago, Illinois, USA.
157. Department of Medicine, Tseung Kwan O Hospital, Hong Kong.
158. Department of Biology, Institute of Biological Sciences and Center for Study on Human Genome, University of São Paulo, São Paulo, Brazil.
159. Department of Pediatrics, Filderklinik, Filderstadt, Germany.
160. Unit of Genetics of Neurodegenerative and Metabolic Diseases, Fondazione IRCCS Istituto Neurologico Carlo Besta, Milan, Italy.
161. Neurology Department, University Hospital of Nancy, UMR 7039, CNRS, Lorraine University, Nancy, France.
162. Department of Pharmacology and Therapeutics, University of Liverpool, Liverpool L69 3GL, UK.
163. The Emmes Company, Rockville, MD, USA.
164. Epilepsy Center Kleinwachau, Radeberg 01454, Germany. .
165. Population Health Sciences, Bristol Medical School, University of Bristol, Bristol BS8 2BN, UK.
166. Department of Genetics, Harvard Medical School, Boston, MA, USA.
167. Department of Neurology, Boston Children’s Health Physicians, Maria Fareri Children’s Hospital at Westchester Medical Center, New York Medical College, New York, NY 10595, USA.
168. Stanford University, Palo Alto, CA, USA.
169. Department of Neurology, Hôpital Pellegrin, Bordeaux, France.
170. Neurology, Northwestern University, Chicago, IL USA.
171. Epilepsy Center for Children, University Hospital Ruppin-Brandenburg, Brandenburg Medical School, 16816 Neuruppin, Germany.
172. MRC Integrative Epidemiology Unit at University of Bristol, Bristol BS8 2BN, UK.
173. Pediatric Neurology, University of Giessen, Germany.
174. Department of Psychiatry, University of Cape Town, South Africa.
175. Kintampo Health Research Centre, Ghana Health Service, Kintampo, Ghana.
176. University of Health and Allied Science in Ho, Ghana.
177. Istanbul University-Cerrahpaşa, Cerrahpaşa Medical Faculty, Department of Neurology, Istanbul, Turkey.
178. Institute for Molecular Medicine Finland (FIMM), University of Helsinki, Helsinki 0014, Finland. .
179. Departments of Psychiatry, Rutgers University, Robert Wood Johnson Medical School and New Jersey Medical School, New Brunswick, NJ, USA.
180. Department of Psychiatry and Behavioral Sciences, SUNY Downstate Medical Center, Brooklyn, NY, USA.
181. University of Pittsburgh, Pittsburgh, PA, USA.
182. Centre for Genomics Research, Discovery Sciences, BioPharmaceuticals R&D, AstraZeneca, Cambridge CB2 0AA, UK.
183. Department of Neurology, Morriston Hospital, Swansea Bay University Bay Health Board, Swansea, Wales, UK.
184. Epilepsy Genetics Program, Division of Epilepsy and Clinical Neurophysiology, Department of Neurology, Boston Children’s Hospital, Boston, MA, USA.
185. Department of Psychiatry, Icahn School of Medicine at Mount Sinai, New York, NY 10029, USA.
186. Department of Genetics and Genomic Sciences, Icahn School of Medicine at Mount Sinai, New York, NY 10029, USA.
187. Department of Complex Trait Genetics, Center for Neurogenomics and Cognitive Research, Amsterdam Neuroscience, VU Amsterdam, Amsterdam, the Netherlands
188. Department of Neurology, Gardner Neuroscience Institute, University of Cincinnati Medical Center, Cincinnati, OH 45220, USA.
189. Department of Psychiatry, Psychosomatic Medicine and Psychotherapy, University Hospital Frankfurt.
190. Department of Psychiatry, Psychotherapy and Psychosomatics, University Hospital Würzburg.
191. Department of Functional Neurology and Epileptology, Hospices Civils de Lyon and University of Lyon, France.
192. Lyon’s Neuroscience Research Center, INSERM U1028 / CNRS UMR 5292, Lyon, France.
193. Department of Clinical Neurosciences, Centre Hospitalo-Universitaire Vaudois, Lausanne, Switzerland.
194. Departments of Neurology, Beth Israel Deaconess Medical Center, Massachusetts General Hospital, and Harvard Medical School, Boston, MA 02215, USA. .
195. Institute of Human Genetics, Bern University Hospital, Bern, Switzerland.
196. Department of Neurology, Ludwig Maximilians University, Munich, Germany.
197. Florey and Murdoch Children’s Research Institutes, Parkville, Victoria 3052, Australia.
198. Department of Paediatrics, The University of Melbourne, Royal Children’s Hospital, Parkville, Victoria 3052, Australia.
199. Department of Neuropediatrics, Children’s Hospital, Goethe University Frankfurt, Frankfurt, Germany.
200. Department of Epileptology, University Hospital Freiburg, Freiburg, Germany.
201. Medical School, Nova Southeastern University, Fort Lauderdale, FL, USA.
202. School of Life Sciences, University of Glasgow, Glasgow G12 8QQ, UK.
203. Psychiatric and Neurodevelopmental Genetics Unit, Center for Genomic Medicine, Massachusetts General Hospital, Boston, MA, USA.
204. Department of Psychiatry, Harvard Medical School, Boston, MA, USA.
205. Division of Neuropsychiatry, Menninger Department of Psychiatry and Behavioral Sciences, Baylor College of Medicine, Houston, TX, USA.
206. Department of Neurology and Comprehensive Epilepsy Center, Thomas Jefferson University, Philadelphia, PA 19107, USA. .
207. Epilepsy Center Kork, Kehl-Kork 77694, Germany.
208. Department of Neurodevelopmental Disorder Genetics, Institute of Brain Science, Nagoya City University Graduate School of Medical Science, Nagoya, Aichi, Japan.
209. Laboratory for Neurogenetics, RIKEN Center for Brain Science, Wako, Saitama, Japan.
210. Centre for Medical Genetics, Vilnius University Hospital Santaros Klinikos, Vilnius, Lithuania.
211. Institute of Biomedical Sciences, Faculty of Medicine, Vilnius University, Vilnius, Lithuania.
212. Department of Child Neurology, Medical School, Marmara University, Istanbul, Turkey.
213. Department of Neurology, UMR 5549, CNRS, Toulouse University Hospital, University of Toulouse, Toulouse, France.
214. DRK-Northern German Epilepsy Centre for Children and Adolescents, 24223 Schwentinental- Raisdorf, Germany.
215. MRC/Wits Rural Public Health & Health Transitions Research Unit (Agincourt), School of Public Health, Faculty of Health Sciences, University of the Witwatersrand, Johannesburg, South Africa.
216. Department of Epidemiology and Global Health, Umeå University, Umeå, Sweden.
217. Department of Clinical Sciences, Neurosciences, Umeå University, Umeå, Sweden.
218. Department of Neurology and Epileptology, University of Aachen, Aachen 52074, Germany.
219. O’Brien Institute for Public Health, University of Calgary, Calgary, Alberta, Canada.
220. Clinical Research Unit, Cumming School of Medicine, University of Calgary, Calgary, Alberta, Canada.
221. Department of Pediatric Neurology, Vivantes Hospital Neukölln, 12351 Berlin, Germany.
222. Department of Child Neurology, Medical School, Dokuz Eylul University, Izmir, Turkey.
223. Stanley Division of Developmental Neurovirology, Johns Hopkins University, Baltimore, Maryland, USA.
224. Bezmialem Vakif University, Institute of Life Sciences and Biotechnology, Istanbul, Turkey.

