## Supplementary Information for "Exome sequencing of 20,979 individuals with epilepsy reveals shared and distinct ultra-rare genetic risk across disorder subtypes"

Epi25 Collaborative

### Supplementary Tables

**Table S1 | Summary of Epi25 epilepsy cohorts.**
DEE: developmental and epileptic encephalopathy; GGE: genetic generalized epilepsy; NAFE: non-acquired focal epilepsy

| Site name | Site code | Epilepsy | | | | Total |
|---|---|---|---|---|---|---|
| | | DEE | GGE | NAFE | Other | |
| Australia: Alfred | AUSALF | 1 | 1 | 4 | 10 | 16 |
| Australia: Melbourne | AUSAUS | 199 | 512 | 521 | 431 | 1663 |
| Australia: Royal Melbourne | AUSRMB | 2 | 146 | 284 | 135 | 567 |
| Austria: Vienna | AUTMUV | 13 | 37 | 25 | 32 | 107 |
| Belgium: Antwerp | BELATW | 93 | 45 | 24 | 1 | 163 |
| Belgium: Brussels | BELULB | 6 | 84 | 204 | 124 | 418 |
| Brazil: Sao Paolo | BRAUSP | 3 | 1 | 0 | 0 | 4 |
| Canada: Calgary | CANCAL | 5 | 60 | 139 | 58 | 262 |
| Canada: Andrade | CANUTN | 43 | 57 | 16 | 18 | 134 |
| Switzerland: Bern | CHEUBB | 24 | 0 | 10 | 5 | 39 |
| Cyprus | CYPCYP | 9 | 57 | 60 | 18 | 144 |
| Czech Republic: Prague | CZEMTH | 16 | 0 | 0 | 0 | 16 |
| Germany: Frankfurt/Marburg | DEUPUM | 8 | 118 | 191 | 147 | 464 |
| Germany: Giessen | DEUUGS | 0 | 0 | 389 | 0 | 389 |
| Germany: Bonn | DEUUKB | 0 | 362 | 1592 | 592 | 2546 |
| Germany: Kiel | DEUUKL | 63 | 103 | 33 | 11 | 210 |
| Germany: Leipzig | DEUULG | 0 | 0 | 105 | 0 | 105 |
| Germany: Tuebingen | DEUUTB | 132 | 543 | 458 | 542 | 1675 |
| Finland: Kuopio | FINKPH | 63 | 87 | 723 | 55 | 928 |
| Finland: Helsinki | FINUVH | 27 | 52 | 23 | 1 | 103 |
| France: Lyon | FRALYU | 0 | 0 | 765 | 0 | 765 |
| Wales: Swansea | GBRSWU | 0 | 103 | 165 | 32 | 300 |
| UK: UCL | GBRUCL | 5 | 347 | 311 | 39 | 702 |
| UK: Imperial/Liverpool | GBRUNL | 0 | 188 | 339 | 0 | 527 |
| Ghana: Kintampo | GHAKNT | 0 | 109 | 45 | 164 | 318 |
| Hong Kong | HKGHKK | 0 | 21 | 418 | 228 | 667 |
| Croatia | HRVUZG | 22 | 6 | 4 | 3 | 35 |
| Ireland: Dublin | IRLRCI | 18 | 176 | 559 | 182 | 935 |
| Italy: Milan | ITAICB | 95 | 173 | 36 | 60 | 364 |
| Italy: Genova | ITAIGI | 342 | 284 | 21 | 71 | 718 |
| Italy: Bologna | ITAUBG | 154 | 104 | 227 | 111 | 596 |
| Italy: Catanzaro | ITAUMC | 6 | 85 | 290 | 51 | 432 |

| | | | | | | |
|---|---|---|---|---|---|---|
| Italy: Florence | ITAUMR | 415 | 258 | 175 | 190 | 1038 |
| Japan: Fukuoka | JPNFKA | 171 | 0 | 3 | 0 | 174 |
| Japan: RIKEN Institute | JPNRKI | 30 | 67 | 0 | 2 | 99 |
| Kenya: Kilifi | KENKIL | 0 | 18 | 35 | 205 | 258 |
| Lebanon: Beirut | LEBABM | 63 | 299 | 619 | 174 | 1155 |
| Lithuania | LTUUHK | 60 | 118 | 95 | 21 | 294 |
| New Zealand: Otago | NZLUTO | 91 | 70 | 87 | 20 | 268 |
| Turkey: Bogazici | TURBZU | 126 | 15 | 14 | 14 | 169 |
| Turkey: Istanbul | TURIBU | 15 | 58 | 81 | 8 | 162 |
| Taiwan | TWNCGM | 16 | 56 | 503 | 110 | 685 |
| USA: BCH | USABCH | 114 | 66 | 29 | 103 | 312 |
| USA: Baylor | USABLC | 0 | 0 | 0 | 223 | 223 |
| USA: Cleveland Clinic | USACCF | 3 | 116 | 153 | 108 | 380 |
| USA: Cincinnati CAE | USACCH | 0 | 358 | 0 | 0 | 358 |
| USA: Philadelphia/CHOP | USACHP | 188 | 597 | 709 | 269 | 1763 |
| USA: Philadelphia/Rowan | USACRW | 0 | 323 | 236 | 0 | 559 |
| USA: EPGP | USAEGP | 126 | 2 | 1 | 1 | 130 |
| USA: FEBSTAT | USAFEB | 0 | 0 | 0 | 31 | 31 |
| USA: NYU HEP | USAHEP | 0 | 0 | 329 | 6 | 335 |
| USA: Lurie | USALCH | 6 | 0 | 0 | 1 | 7 |
| USA: MGH Partners Biobank | USAMGH | 0 | 64 | 0 | 0 | 64 |
| USA: MONEAD | USAMON | 1 | 91 | 166 | 36 | 294 |
| USA: Mt.Sinai | USAMSS | 36 | 45 | 35 | 21 | 137 |
| USA: Nationwide Childrens | USANCH | 0 | 309 | 0 | 5 | 314 |
| USA: Penn/CHOP | USAUPN | 54 | 196 | 333 | 266 | 849 |
| USA: Vanderbilt (BioVU) | USAVAN | 0 | 301 | 169 | 13 | 483 |
| South Africa: Aguincourt | ZAFAGN | 0 | 81 | 53 | 71 | 205 |
| **Total** | | **2864** | **7369** | **11806** | **5019** | **27058** |

**Table S2 | Summary of control collection.**

| Control collection | General control | AFIB/CAD/IBD* | Total |
|---|---|---|---|
| Mass General Brigham (MGB) Biobank | 11473 | 1433 | 12906 |
| National Institute of Diabetes and Digestive and Kidney (NIDDK) | 3762 | 4120 | 7882 |
| Epi25 controls | 5294 | 0 | 5294 |
| Genomic Psychiatry Cohort (GPC) | 4684 | 0 | 4684 |
| USA controls | 3619 | 0 | 3619 |
| Avon Longitudinal Study of Parents and Children (ALSPAC) | 2971 | 0 | 2971 |
| UK/IRL controls | 1220 | 0 | 1220 |
| MIGen Leicester controls | 1100 | 0 | 1100 |
| Dutch controls | 939 | 0 | 939 |
| Epi4K controls | 711 | 0 | 711 |
| FINRISK controls | 670 | 0 | 670 |
| German controls | 414 | 0 | 414 |
| Hong Kong controls | 114 | 0 | 114 |
| Italian controls | 106 | 0 | 106 |
| **Total** | **37077** | **5553** | **42630** |

* Atrial fibrillation, coronary artery disease, or inflammatory bowel disease.

**Table S3 | Summary of sample quality control (QC).**

| Sample QC metric | Number of cases (%) | Number of controls (%) |
|---|---|---|
| **Initial number** | **28955 (100%)** | **42630 (100%)** |
| **Hard filters**<br>Mean call rate ≥0.90<br>Mean genotype quality (GQ) ≥57<br>Mean depth (DP) ≥25<br>Freemix contamination estimate ≤2.5%<br>Percent chimeric reads ≤2% | 26947 (93.1%) | 41195 (96.7%) |
| **Sex check**<br>Concordance between imputed sex and reported sex | 26441 (91.3%) | 40914 (96.0%) |
| **Ancestry assignment**<br>Predicted genetic ancestry probability ≥0.9<br>Case-control ancestry matching (PCA) | 22080 (76.3%) | 35119 (82.4%) |
| NFE | 16765 | 26951 |
| AFR | 1746 | 2744 |
| EAS | 1741 | 1243 |
| FIN | 1052 | 582 |
| AMR | 513 | 3666 |
| SAS | 254 | 358 |
| **Relatedness filtering**<br>Identity by descent (IBD) ≤0.2 | 21212 (73.3%) | 33593 (78.8%) |
| **Outlier filters**<br>Transition/transversion,<br>heterozygous/homozygous,<br>and insertion/deletion ratio<br>≤4 standard deviations from the mean | 21061 (72.7%) | 33447 (78.5%) |
| **Residual population stratification control**<br>Synonymous singleton count outlier filter<br>Case-control ancestry matching (post-QC PCA) | 20979 (72.5%) | 33444 (78.5%) |
| NFE | 16040 | 25641 |
| EAS | 1698 | 1215 |
| AFR | 1598 | 2592 |
| FIN | 926 | 537 |
| AMR | 480 | 3106 |
| SAS | 237 | 353 |

### Supplementary Figures

**Figure S1 | Hard filters in sample QC.**

**a-e**, The distribution (x-axis) and thresholding (dashed vertical line) of each sample QC metric by cohort (y-axis) and case/control status (color). Mean call rate $\geq$0.90 (**a**), mean genotype quality $\geq$57 (**b**), mean depth $\geq$25 (**c**), freemix contamination estimate $\leq$2.5% (**d**), and percent chimeric reads $\leq$2% (**e**) were applied for hard filtering.

a

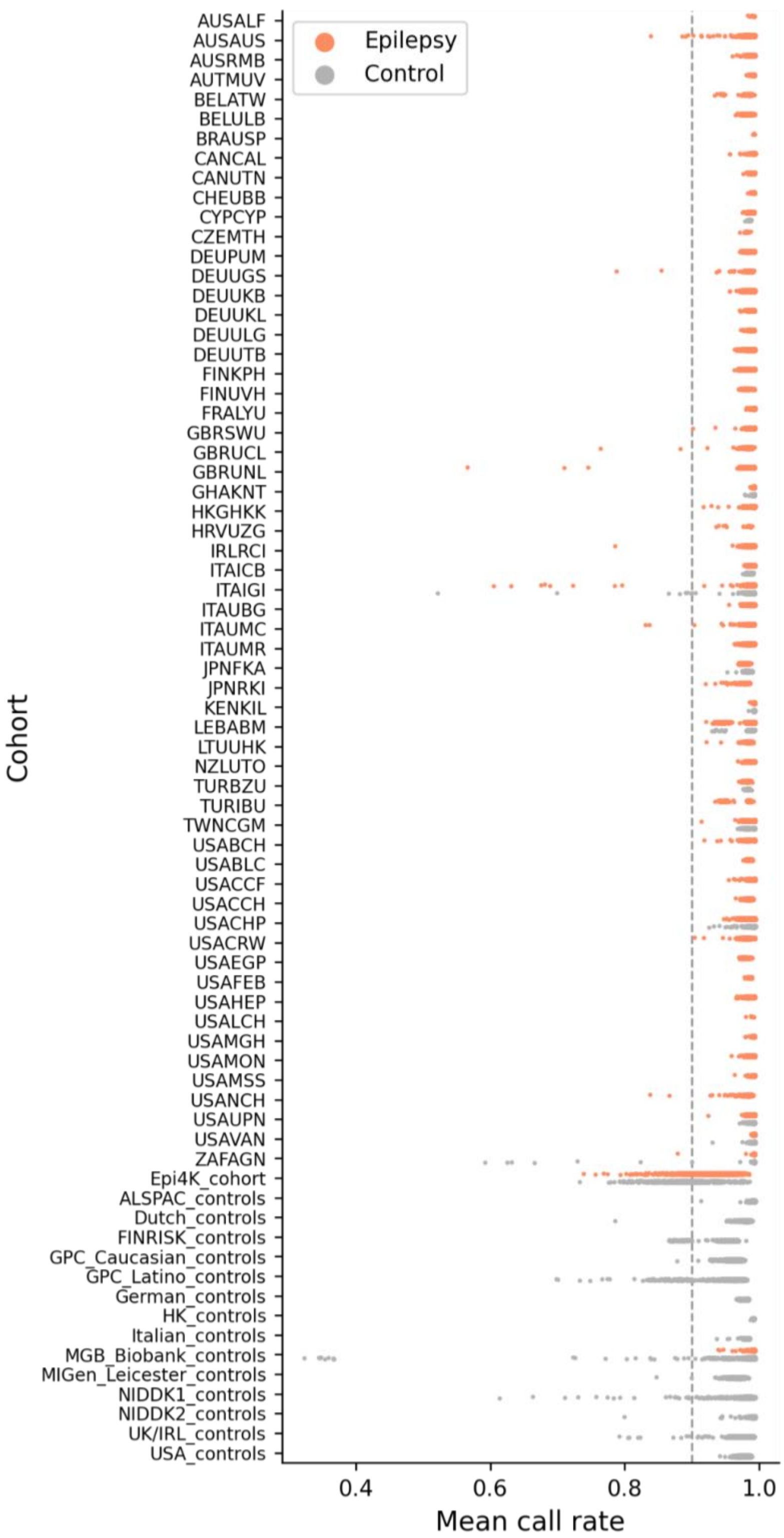

Epilepsy
Control
AUSALF
AUSAUS
AUSRMB
AUTMUV
BELATW
BELULB
BRAUSP
CANCAL
CANUTN
CHEUBB
CYPCYP
CZEMTH
DEUPUM
DEUUGS
DEUUKB
DEUUKL
DEUULG
DEUUTB
FINKPH
FINUVH
FRALYU
GBRSWU
GBRUCL
GBRUNL
GHAKNT
HKGHKK
HRVUZG
IRLRCI
ITAICB
ITAIGI
ITAUBG
ITAUMC
ITAUMR
JPNFKA
JPNRKI
KENKIL
LEBABM
LTUUHK
NZLUTO
TURBZU
TURIBU
TWNCGM
USABCH
USABLC
USACCF
USACCH
USACHP
USACRW
USAEGP
USAFEB
USAHEP
USALCH
USAMGH
USAMON
USAMSS
USANCH
USAUPN
USAVAN
ZAFAGN
Epi4K_cohort
ALSPAC_controls
Dutch_controls
FINRISK_controls
GPC_Caucasian_controls
GPC_Latino_controls
German_controls
HK_controls
Italian_controls
MGB_Biobank_controls
MIGen_Leicester_controls
NIDDK1_controls
NIDDK2_controls
UK/IRL_controls
USA_controls
Cohort
0.4
0.6
0.8
1.0
Mean call rate

b

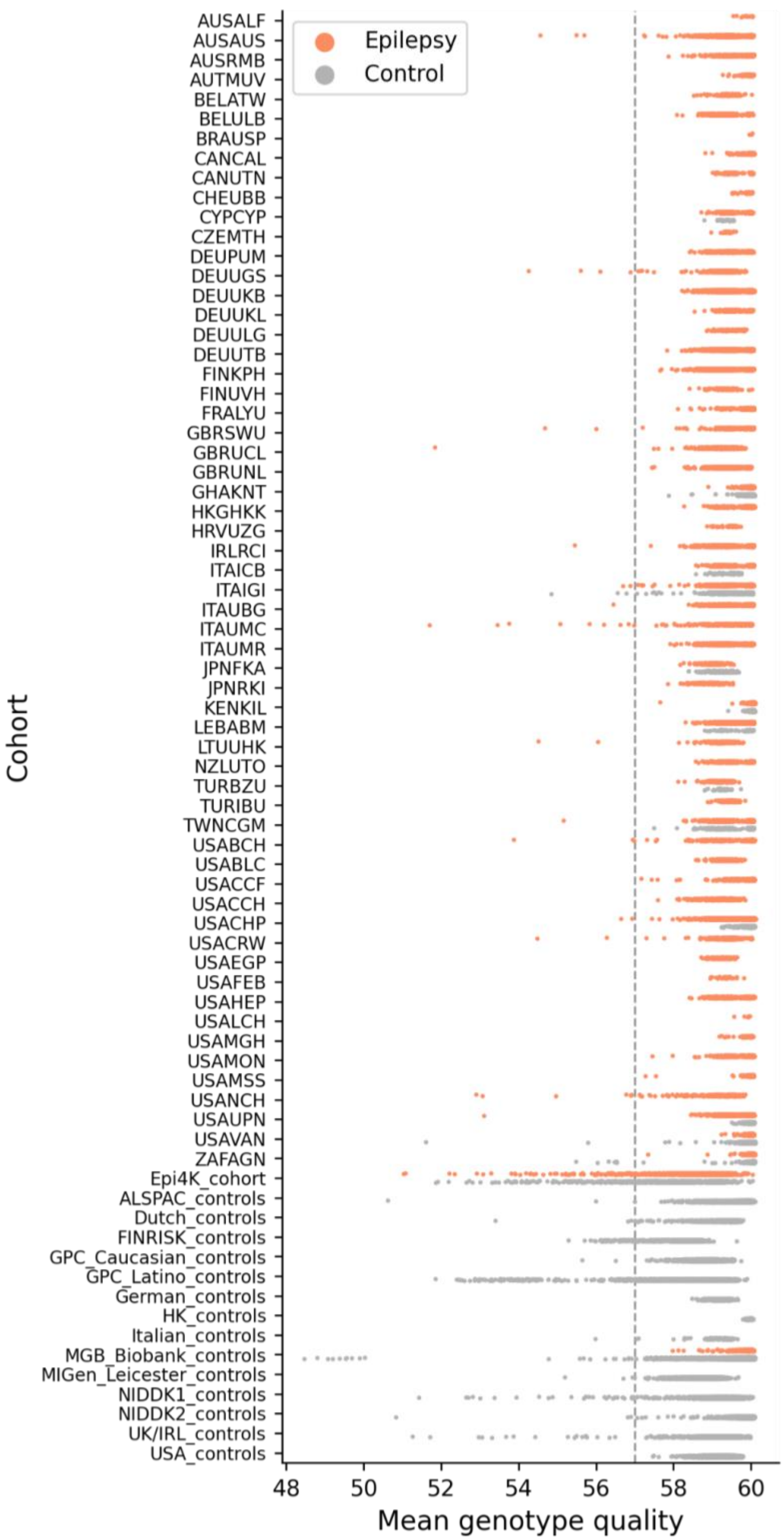

Epilepsy
Control
Cohort
AUSALF
AUSAUS
AUSRMB
AUTMUV
BELATW
BELULB
BRAUSP
CANCAL
CANUTN
CHEUBB
CYPCYP
CZEMTH
DEUPUM
DEUUGS
DEUUKB
DEUUKL
DEUULG
DEUUTB
FINKPH
FINUVH
FRALYU
GBRSWU
GBRUCL
GBRUNL
GHAKNT
HKGHKK
HRVUZG
IRLRCI
ITAICB
ITAIGI
ITAUBG
ITAUMC
ITAUMR
JPNFKA
JPNRKI
KENKIL
LEBABM
LTUUHK
NZLUTO
TURBZU
TURIBU
TWNCGM
USABCH
USABLC
USACCF
USACCH
USACHP
USACRW
USAEGP
USAFEB
USAHEP
USALCH
USAMGH
USAMON
USAMSS
USANCH
USAUPN
USAVAN
ZAFAGN
Epi4K_cohort
ALSPAC_controls
Dutch_controls
FINRISK_controls
GPC_Caucasian_controls
GPC_Latino_controls
German_controls
HK_controls
Italian_controls
MGB_Biobank_controls
MIGen_Leicester_controls
NIDDK1_controls
NIDDK2_controls
UK/IRL_controls
USA_controls
48
50
52
54
56
58
60
Mean genotype quality

c

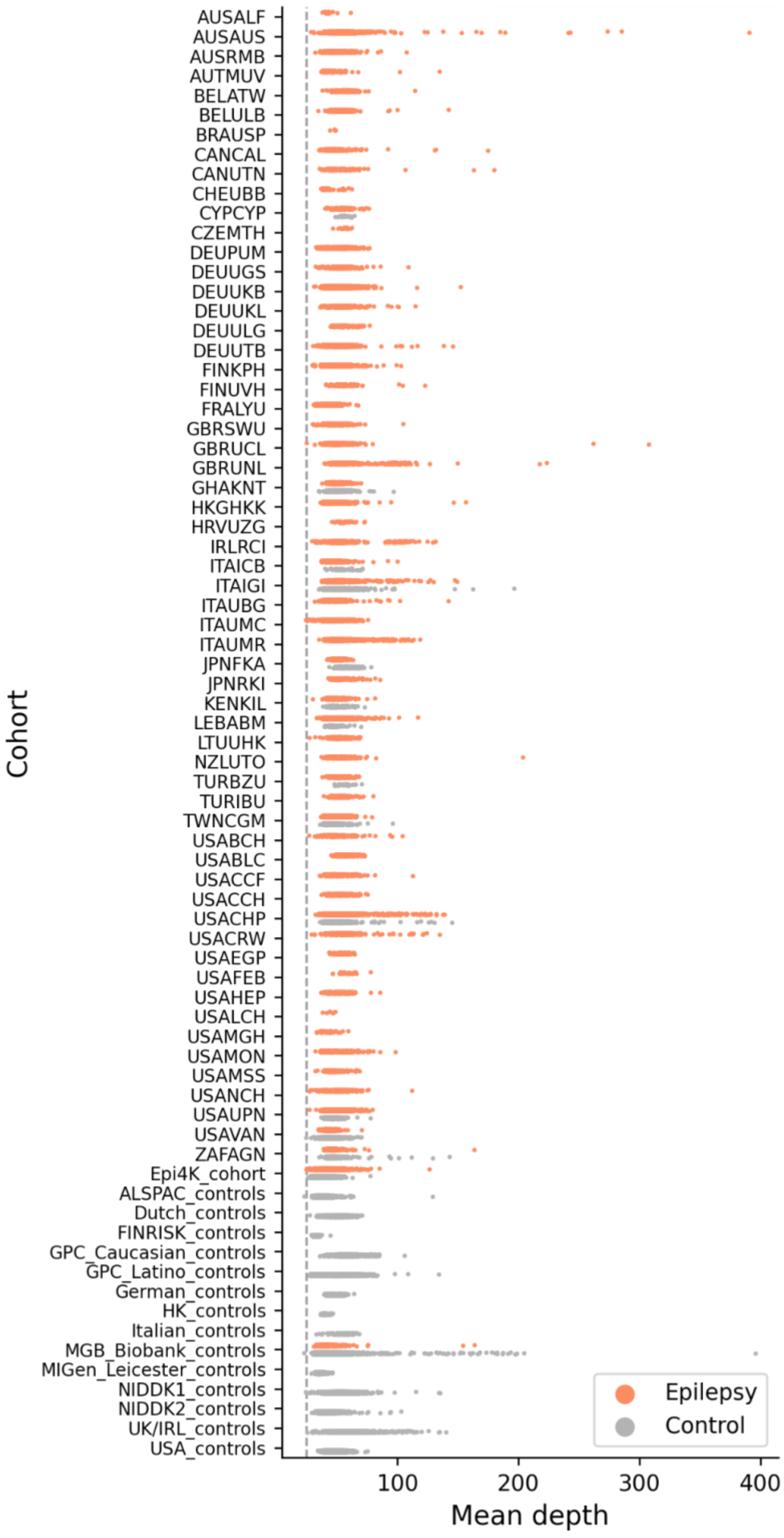
AUSALF
AUSAUS
AUSRMB
AUTMUV
BELATW
BELULB
BRAUSP
CANCAL
CANUTN
CHEUBB
CYPCYP
CZEMTH
DEUPUM
DEUUGS
DEUUKB
DEUUKL
DEUULG
DEUUTB
FINKPH
FINUVH
FRALYU
GBRSWU
GBRUCL
GBRUNL
GHAKNT
HKGHKK
HRVUZG
IRLRCI
ITAICB
ITAIGI
ITAUBG
ITAUMC
ITAUMR
JPNFKA
JPNRKI
KENKIL
LEBABM
LTUUHK
NZLUTO
TURBZU
TURIBU
TWNCGM
USABCH
USABLC
USACCF
USACCH
USACHP
USACRW
USAEGP
USAFEB
USAHEP
USALCH
USAMGH
USAMON
USAMSS
USANCH
USAUPN
USAVAN
ZAFAGN
Epi4K_cohort
ALSPAC_controls
Dutch_controls
FINRISK_controls
GPC_Caucasian_controls
GPC_Latino_controls
German_controls
HK_controls
Italian_controls
MGB_Biobank_controls
MIGen_Leicester_controls
NIDDK1_controls
NIDDK2_controls
UK/IRL_controls
USA_controls
Cohort
100
200
300
400
Mean depth
Epilepsy
Control

d

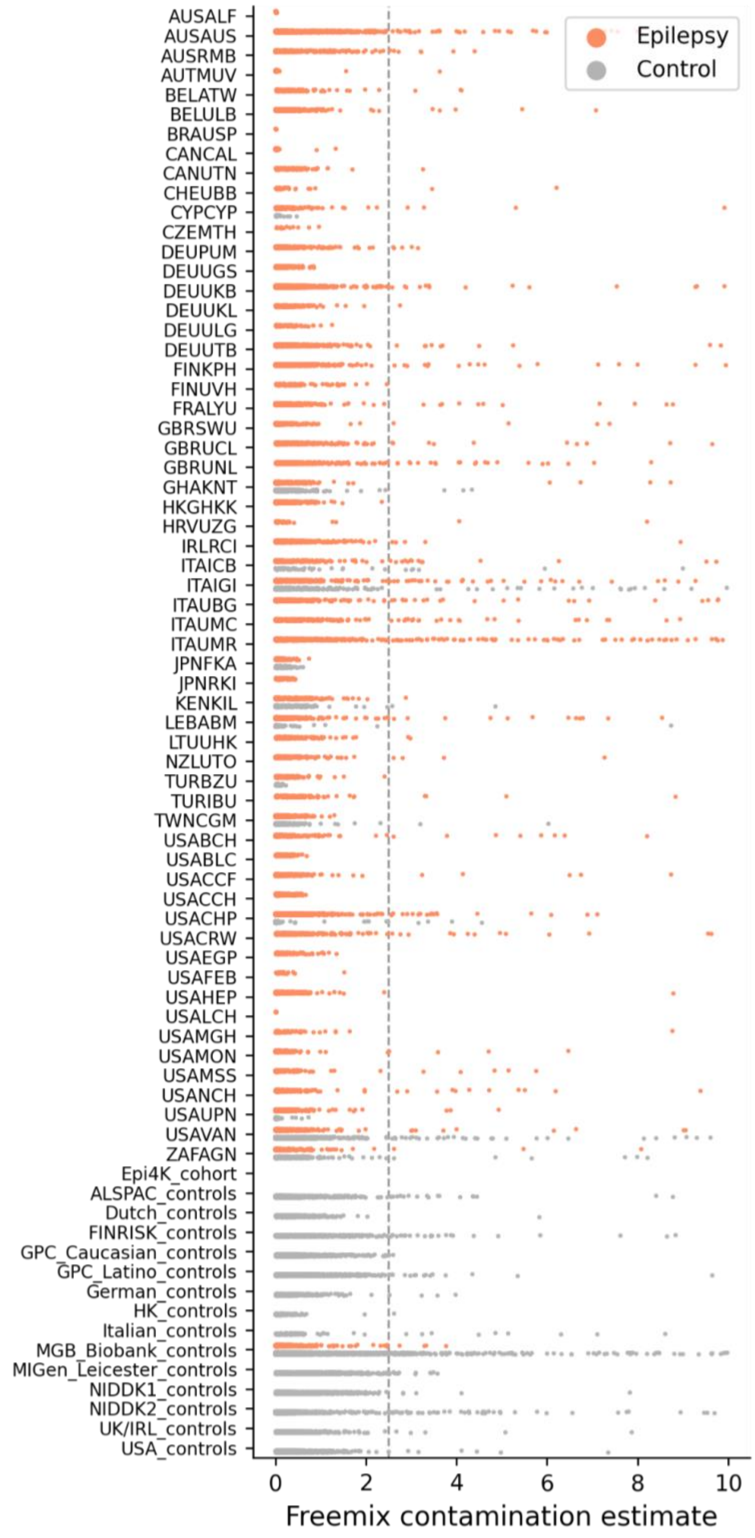

Epilepsy
Control
Cohort
AUSALF
AUSAUS
AUSRMB
AUTMUV
BELATW
BELULB
BRAUSP
CANCAL
CANUTN
CHEUBB
CYPCYP
CZEMTH
DEUPUM
DEUUGS
DEUUKB
DEUUKL
DEUULG
DEUUTB
FINKPH
FINUVH
FRALYU
GBRSWU
GBRUCL
GBRUNL
GHAKNT
HKGHKK
HRVUZG
IRLRCI
ITAICB
ITAIGI
ITAUBG
ITAUMC
ITAUMR
JPNFKA
JPNRKI
KENKIL
LEBABM
LTUUHK
NZLUTO
TURBZU
TURIBU
TWNCGM
USABCH
USABLC
USACCF
USACCH
USACHP
USACRW
USAEGP
USAFEB
USAHEP
USALCH
USAMGH
USAMON
USAMSS
USANCH
USAUPN
USAVAN
ZAFAGN
Epi4K_cohort
ALSPAC_controls
Dutch_controls
FINRISK_controls
GPC_Caucasian_controls
GPC_Latino_controls
German_controls
HK_controls
Italian_controls
MGB_Biobank_controls
MIGen_Leicester_controls
NIDDK1_controls
NIDDK2_controls
UK/IRL_controls
USA_controls
0
2
4
6
8
10
Freemix contamination estimate

e

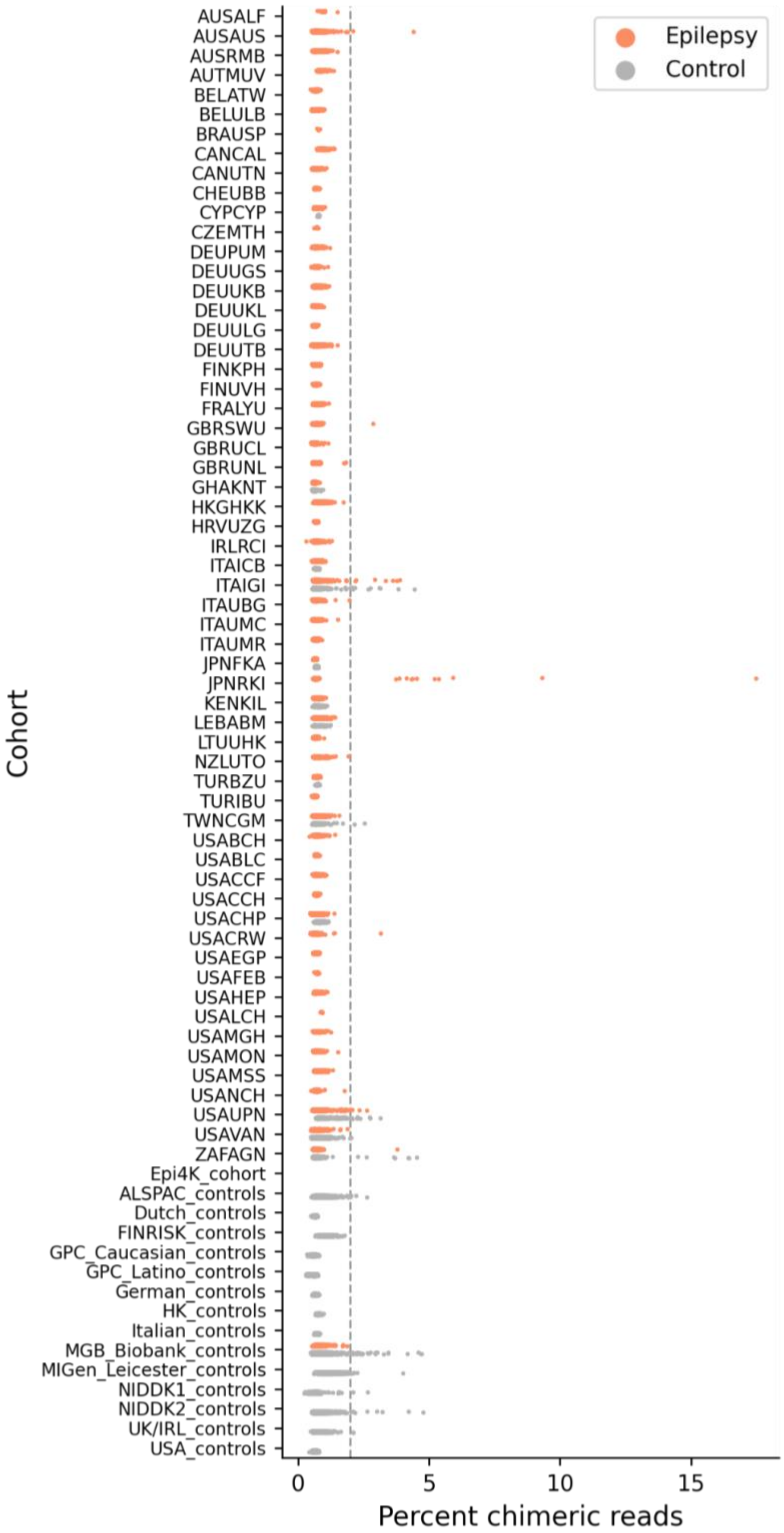

Epilepsy
Control
Cohort
AUSALF
AUSAUS
AUSRMB
AUTMUV
BELATW
BELULB
BRAUSP
CANCAL
CANUTN
CHEUBB
CYPCYP
CZEMTH
DEUPUM
DEUUGS
DEUUKB
DEUUKL
DEUULG
DEUUTB
FINKPH
FINUVH
FRALYU
GBRSWU
GBRUCL
GBRUNL
GHAKNT
HKGHKK
HRVUZG
IRLRCI
ITAICB
ITAIGI
ITAUBG
ITAUMC
ITAUMR
JPNFKA
JPNRKI
KENKIL
LEBABM
LTUUHK
NZLUTO
TURBZU
TURIBU
TWNCGM
USABCH
USABLC
USACCF
USACCH
USACHP
USACRW
USAEGP
USAFEB
USAHEP
USALCH
USAMGH
USAMON
USAMSS
USANCH
USAUPN
USAVAN
ZAFAGN
Epi4K_cohort
ALSPAC_controls
Dutch_controls
FINRISK_controls
GPC_Caucasian_controls
GPC_Latino_controls
German_controls
HK_controls
Italian_controls
MGB_Biobank_controls
MIGen_Leicester_controls
NIDDK1_controls
NIDDK2_controls
UK/IRL_controls
USA_controls
0
5
10
15
Percent chimeric reads

**Figure S2 | Sex check in sample QC.**

Sex was imputed using the X-chromosome homozygosity rate, or F-statistics (y-axis): ≤0.5 were female and ≥0.8 were male; otherwise unknown. Samples with either unknown genetic sex or discordance between imputed and reported sex were excluded.

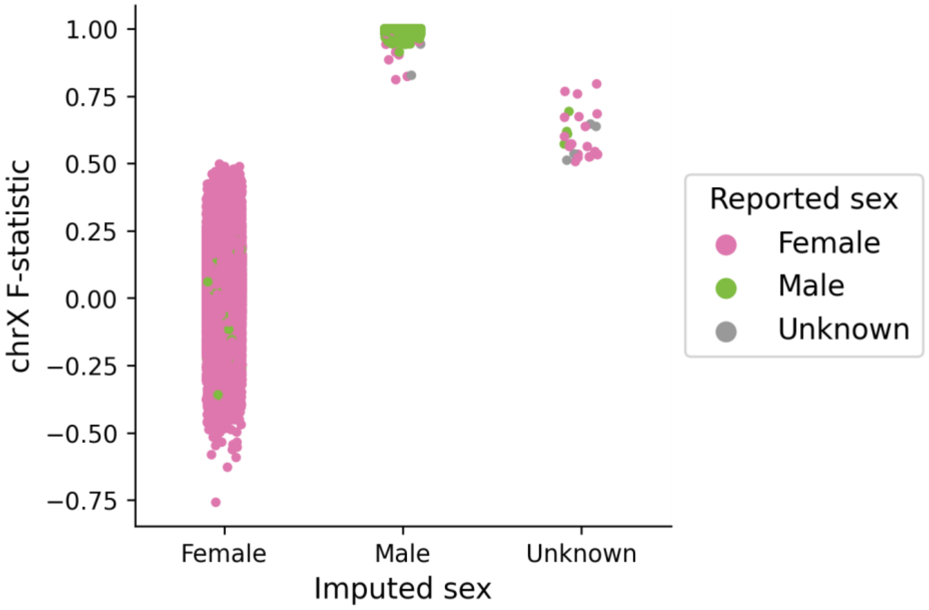

**Figure S3 | Relatedness filtering in sample QC.**

The relationship between each pair of samples within the same assigned ancestry) was inferred using PLINK's $\hat{\pi}$ ("pi-hat"), which estimates the probability of any two samples sharing 0, 1, or 2 alleles (identity-by-decent; $IBD_0$, $IBD_1$, $IBD_2$): $\hat{\pi}$>0.707 were duplicates or monozygotic twins, [0.354, 0.707) were first-degree relatives, [0.177,0.354) were second-degree relatives. One sample in each of the related pairs with a $\hat{\pi}$ >0.2 was excluded. For clarity, only pairs of samples with a $\hat{\pi}$ >0.125 and up to 1,000 pairs per relationship category are shown.

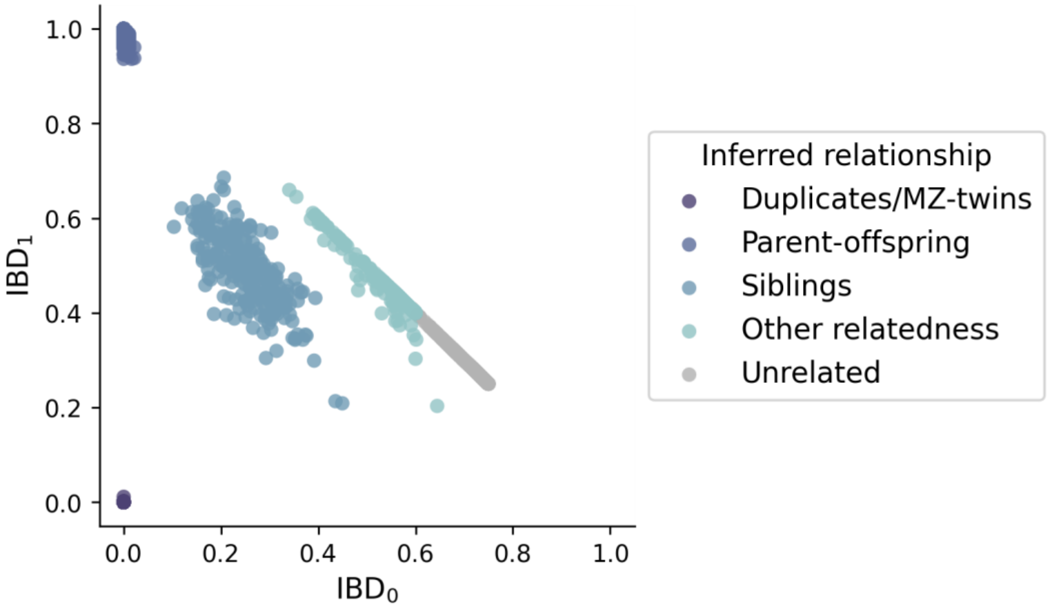

**Figure S4 | Outlier filters in sample QC.**

**a-d**, The distribution (x-axis) and outliers (blue color) of each sample QC metric by cohort (y-axis). Samples with >4 standard deviations from the mean of transition/transversion ratio (**a**), heterozygous/homozygous ratio (**b**), insertion/deletion ratio (**c**), or synonymous singleton count (**d**) within each cohort were excluded. **e**, The distribution of synonymous singleton count (after outlier filtering in **d**; x-axis) by cohort (y-axis) and case/control status (color). Cohorts are ordered by the mean value of synonymous singleton count; the vertical dashed lines indicate the mean values of all cases (orange) and controls (gray), respectively. Cohorts with extreme synonymous singleton counts were excluded (Italian_controls, LEBABM, TURIBU, TURBZU, CYPCYP).

a

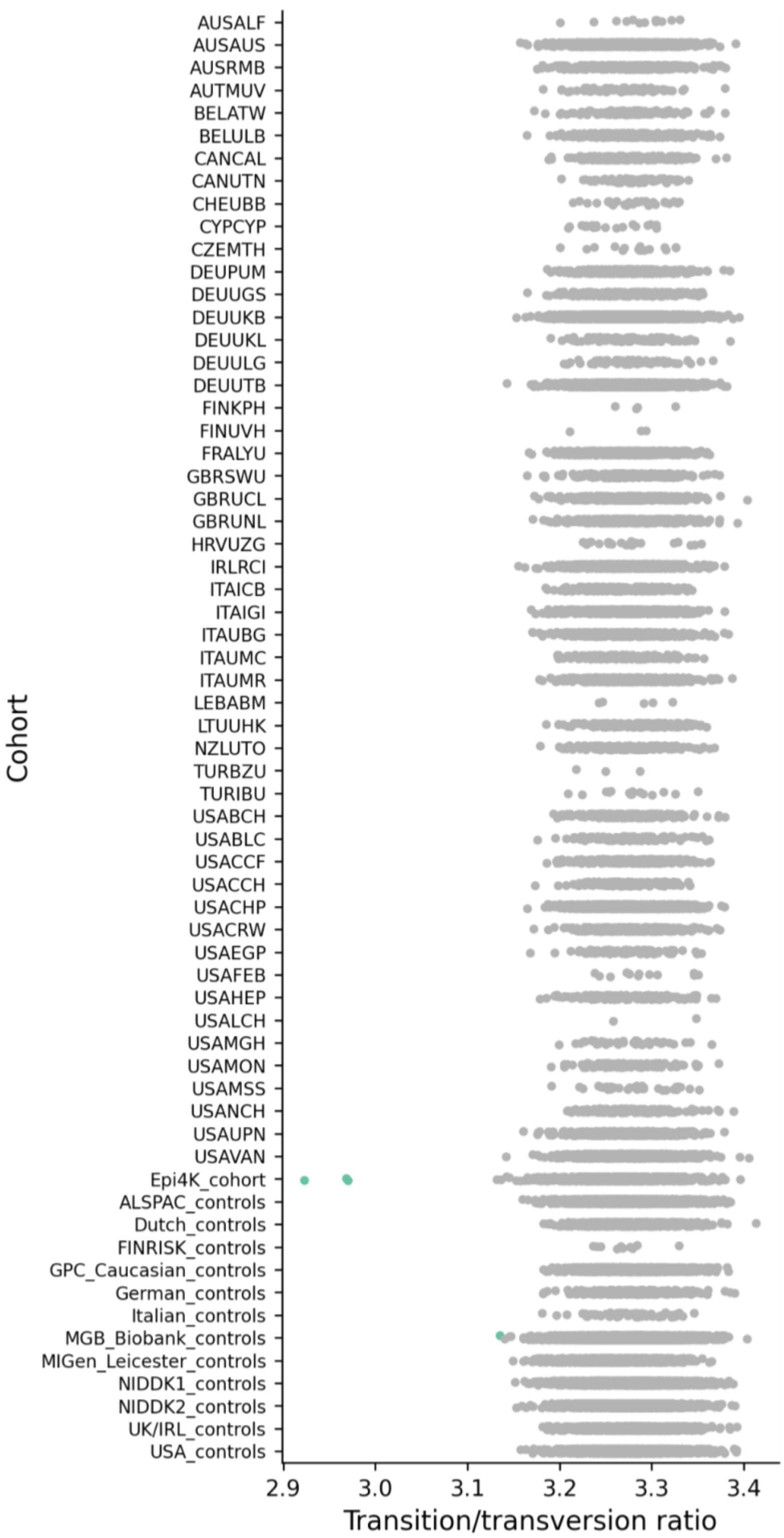

AUSALF
AUSAUS
AUSRMB
AUTMUV
BELATW
BELULB
CANCAL
CANUTN
CHEUBB
CYPCYP
CZEMTH
DEUPUM
DEUUGS
DEUUKB
DEUUKL
DEUULG
DEUUTB
FINKPH
FINUVH
FRALYU
GBRSWU
GBRUCL
GBRUNL
HRVUZG
IRLRCI
ITAICB
ITAIGI
ITAUBG
ITAUMC
ITAUMR
LEBABM
LTUUHK
NZLUTO
TURBZU
TURIBU
USABCH
USABLC
USACCF
USACCH
USACHP
USACRW
USAEGP
USAFEB
USAHEP
USALCH
USAMGH
USAMON
USAMSS
USANCH
USAUPN
USAVAN
Epi4K_cohort
ALSPAC_controls
Dutch_controls
FINRISK_controls
GPC_Caucasian_controls
German_controls
Italian_controls
MGB_Biobank_controls
MIGen_Leicester_controls
NIDDK1_controls
NIDDK2_controls
UK/IRL_controls
USA_controls
Cohort
2.9
3.0
3.1
3.2
3.3
3.4
Transition/transversion ratio

b

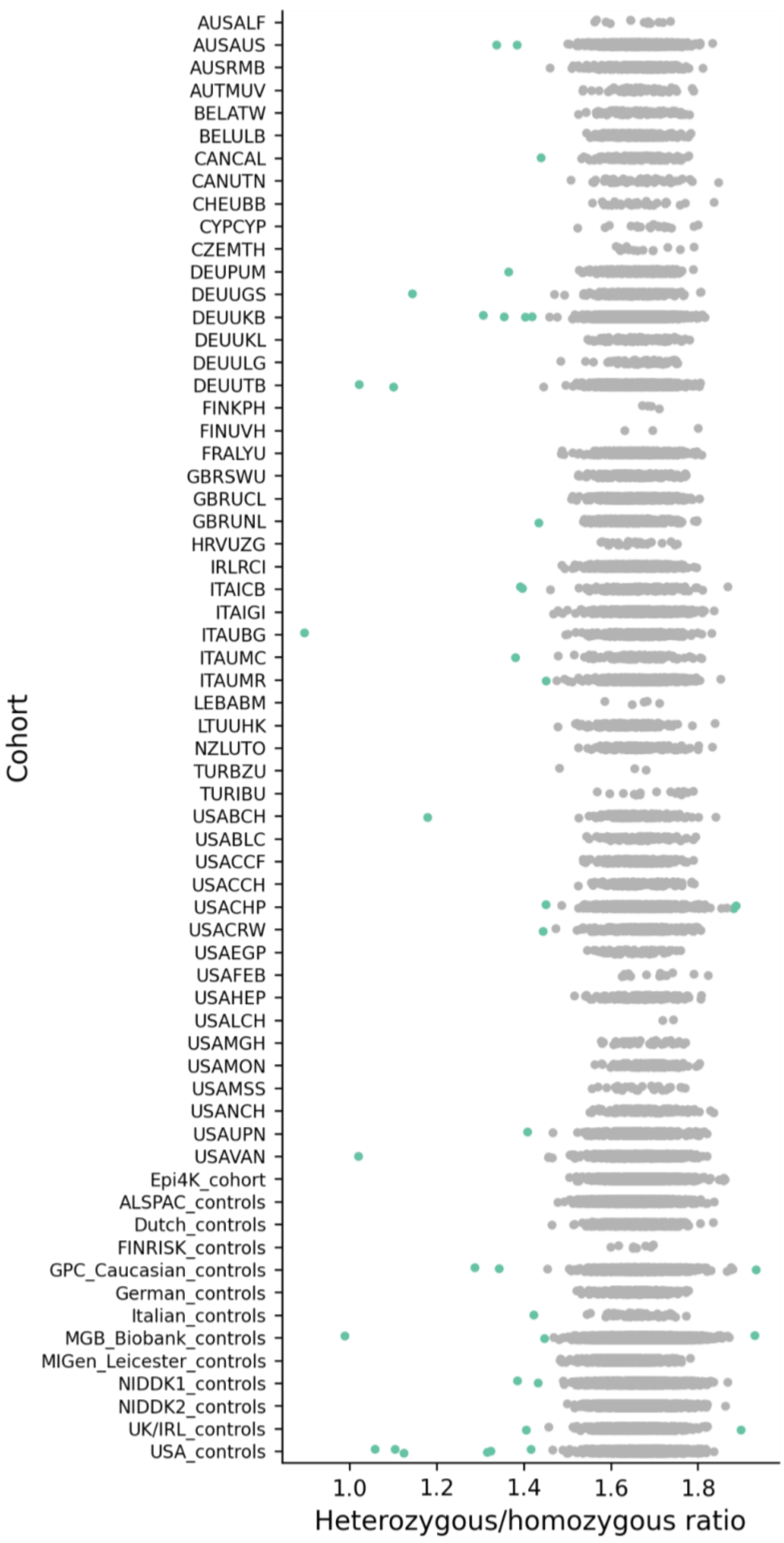
Cohort
AUSALF
AUSAUS
AUSRMB
AUTMUV
BELATW
BELULB
CANCAL
CANUTN
CHEUBB
CYPCYP
CZEMTH
DEUPUM
DEUUGS
DEUUKB
DEUUKL
DEUULG
DEUUTB
FINKPH
FINUVH
FRALYU
GBRSWU
GBRUCL
GBRUNL
HRVUZG
IRLRCI
ITAICB
ITAIGI
ITAUBG
ITAUMC
ITAUMR
LEBABM
LTUUHK
NZLUTO
TURBZU
TURIBU
USABCH
USABLC
USACCF
USACCH
USACHP
USACRW
USAEGP
USAFEB
USAHEP
USALCH
USAMGH
USAMON
USAMSS
USANCH
USAUPN
USAVAN
Epi4K_cohort
ALSPAC_controls
Dutch_controls
FINRISK_controls
GPC_Caucasian_controls
German_controls
Italian_controls
MGB_Biobank_controls
MIGen_Leicester_controls
NIDDK1_controls
NIDDK2_controls
UK/IRL_controls
USA_controls
1.0
1.2
1.4
1.6
1.8
Heterozygous/homozygous ratio

c

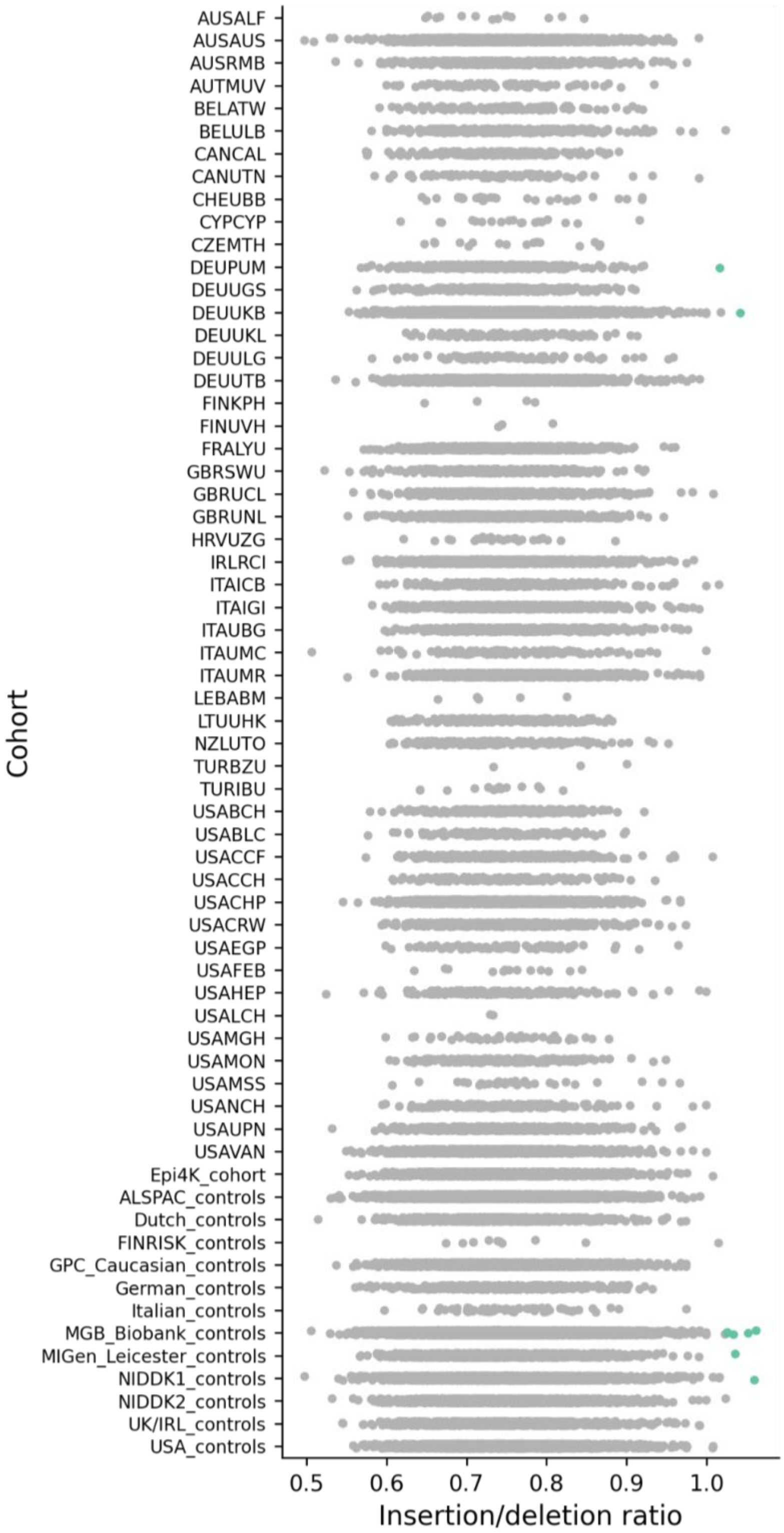

Cohort
AUSALF
AUSAUS
AUSRMB
AUTMUV
BELATW
BELULB
CANCAL
CANUTN
CHEUBB
CYPCYP
CZEMTH
DEUPUM
DEUUGS
DEUUKB
DEUUKL
DEUULG
DEUUTB
FINKPH
FINUVH
FRALYU
GBRSWU
GBRUCL
GBRUNL
HRVUZG
IRLRCI
ITAICB
ITAIGI
ITAUBG
ITAUMC
ITAUMR
LEBABM
LTUUHK
NZLUTO
TURBZU
TURIBU
USABCH
USABLC
USACCF
USACCH
USACHP
USACRW
USAEGP
USAFEB
USAHEP
USALCH
USAMGH
USAMON
USAMSS
USANCH
USAUPN
USAVAN
Epi4K_cohort
ALSPAC_controls
Dutch_controls
FINRISK_controls
GPC_Caucasian_controls
German_controls
Italian_controls
MGB_Biobank_controls
MIGen_Leicester_controls
NIDDK1_controls
NIDDK2_controls
UK/IRL_controls
USA_controls
0.5
0.6
0.7
0.8
0.9
1.0
Insertion/deletion ratio

d

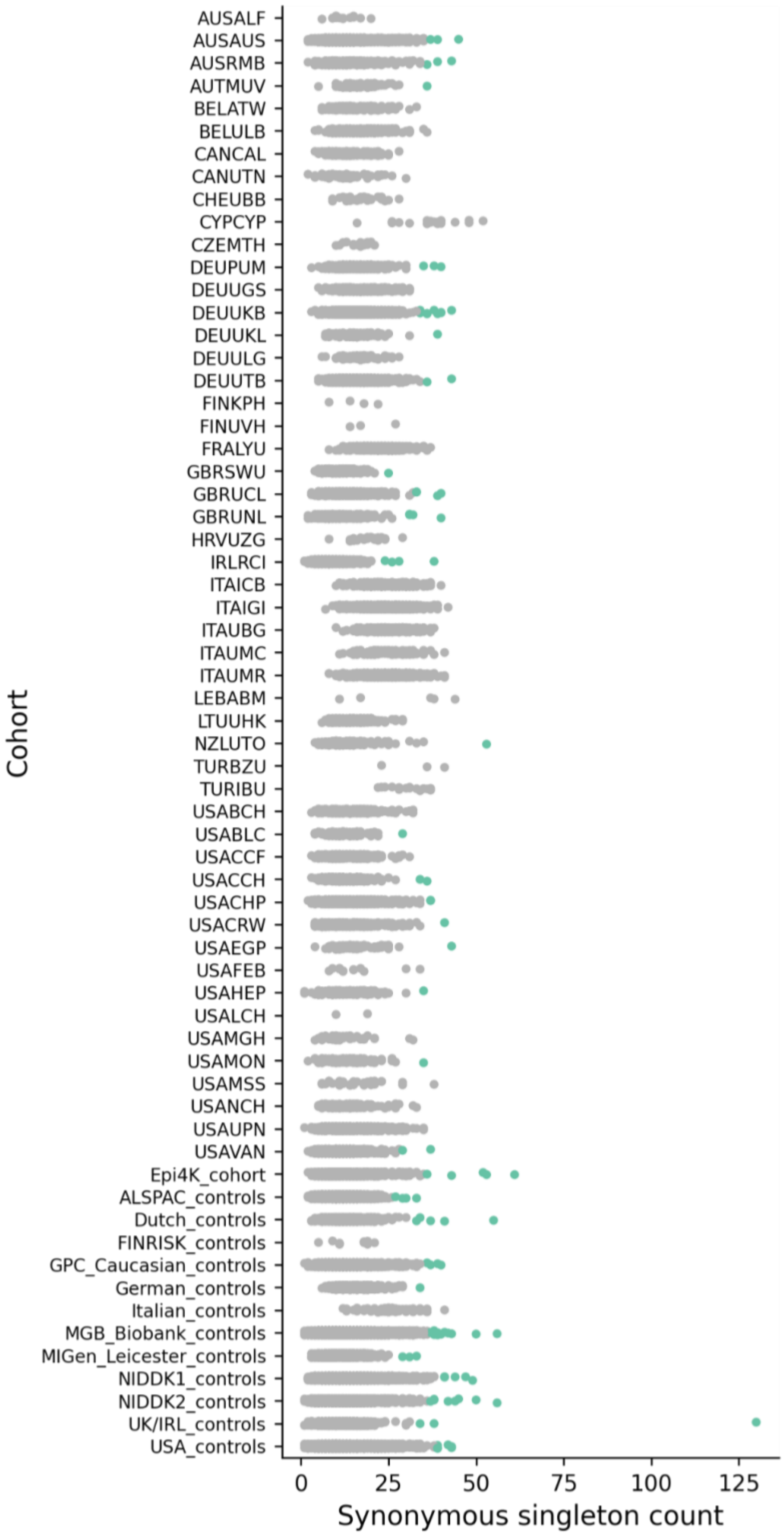

Cohort
AUSALF
AUSAUS
AUSRMB
AUTMUV
BELATW
BELULB
CANCAL
CANUTN
CHEUBB
CYPCYP
CZEMTH
DEUPUM
DEUUGS
DEUUKB
DEUUKL
DEUULG
DEUUTB
FINKPH
FINUVH
FRALYU
GBRSWU
GBRUCL
GBRUNL
HRVUZG
IRLRCI
ITAICB
ITAIGI
ITAUBG
ITAUMC
ITAUMR
LEBABM
LTUUHK
NZLUTO
TURBZU
TURIBU
USABCH
USABLC
USACCF
USACCH
USACHP
USACRW
USAEGP
USAFEB
USAHEP
USALCH
USAMGH
USAMON
USAMSS
USANCH
USAUPN
USAVAN
Epi4K_cohort
ALSPAC_controls
Dutch_controls
FINRISK_controls
GPC_Caucasian_controls
German_controls
Italian_controls
MGB_Biobank_controls
MIGen_Leicester_controls
NIDDK1_controls
NIDDK2_controls
UK/IRL_controls
USA_controls
0
25
50
75
100
125
Synonymous singleton count

e

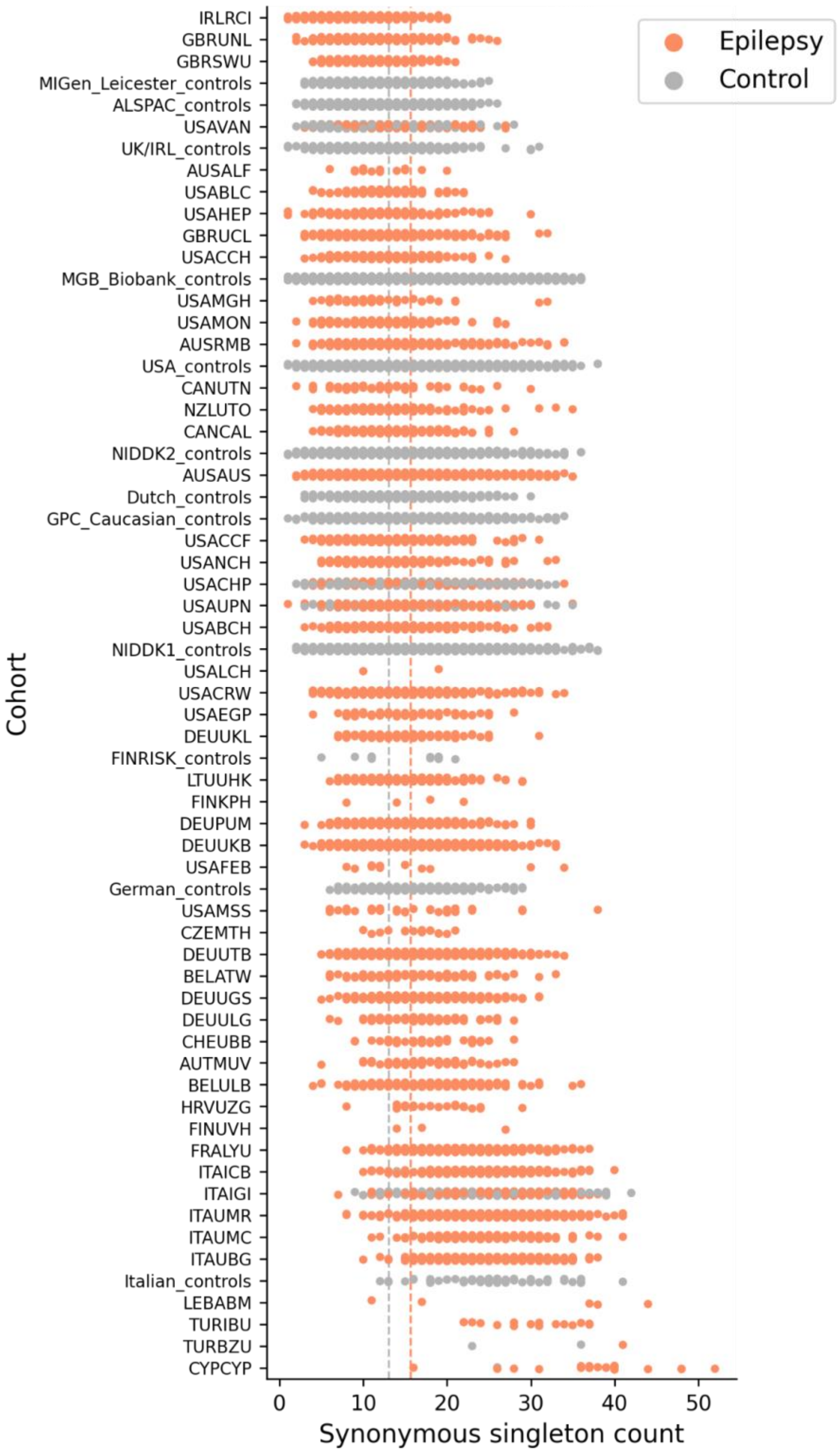
Epilepsy
Control
Cohort
IRLRCI
GBRUNL
GBRSWU
MIGen_Leicester_controls
ALSPAC_controls
USAVAN
UK/IRL_controls
AUSALF
USABLC
USAHEP
GBRUCL
USACCH
MGB_Biobank_controls
USAMGH
USAMON
AUSRMB
USA_controls
CANUTN
NZLUTO
CANCAL
NIDDK2_controls
AUSAUS
Dutch_controls
GPC_Caucasian_controls
USACCF
USANCH
USACHP
USAUPN
USABCH
NIDDK1_controls
USALCH
USACRW
USAEGP
DEUUKL
FINRISK_controls
LTUUHK
FINKPH
DEUPUM
DEUUKB
USAFEB
German_controls
USAMSS
CZEMTH
DEUUTB
BELATW
DEUUGS
DEUULG
CHEUBB
AUTMUV
BELULB
HRVUZG
FINUVH
FRALYU
ITAICB
ITAIGI
ITAUMR
ITAUMC
ITAUBG
Italian_controls
LEBABM
TURIBU
TURBZU
CYPCYP
0
10
20
30
40
50
Synonymous singleton count

**Figure S5 | Ancestry assignment of post-QC samples.**

**a**, Inferred ancestry of samples that passed all sample QCs. Genetic ancestry of epilepsy cases and controls were inferred using a random forest model trained on the 1000 Genomes data, based on the top six principle components (PCs); samples with a probability ≥0.9 to be one of the six populations – Non-Finnish European (NFE), Finnish (FIN), African (AFR), East Asian (EAS), South Asian (SAS), Ad Mixed American (AMR) – were retained. **b**, The distribution of epilepsy cases and controls across the inferred ancestral groups.

a

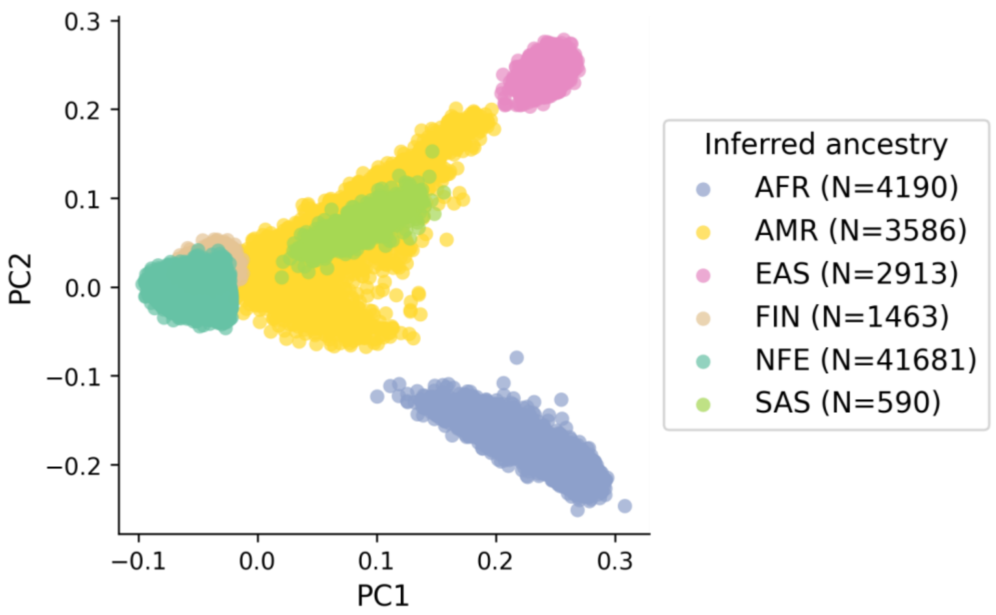

b

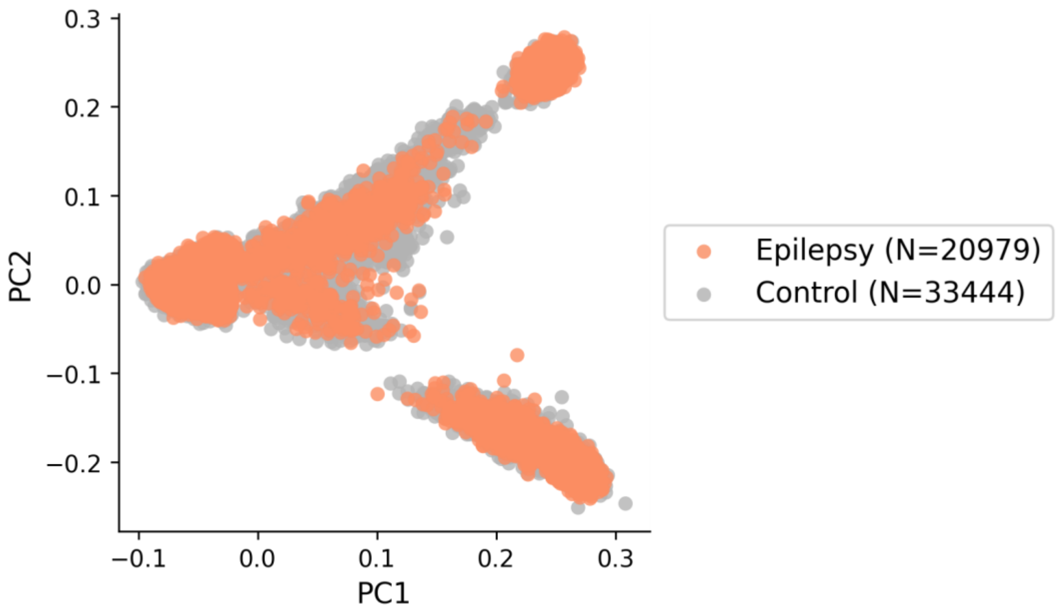

### Supplementary Data Descriptions

**Supplementary Data 1 | Results from exome-wide gene-based burden analysis of URVs. a**,**b**, Burden of protein-truncating (**a**) and damaging missense (MPC≥2; **b**) URVs in each protein-coding gene with at least one epilepsy or control carrier. For each variant class, burden analyses are performed across four epilepsy groups – 1,938 DEEs, 5,499 GGE, 9,219 NAFE, and 20,979 epilepsy-affected individuals combined ('EPI') – versus 33,444 controls. *P* values are computed using a Firth logistic regression model with adjustment for sex and ancestry.

**Supplementary Data 2 | Results from burden analysis of GATOR1 genes.** Burden of protein-truncating URVs in GATOR1 complex and the three GATOR1-encoing genes (*DEPDC5*, *NPRL3*, and *NPRL2*), analyzed separately for familial (N=1162) and non-familial (N=8,014) NAFE cases. *P* values are computed using a Firth logistic regression model with adjustment for sex and ancestry.

**Supplementary Data 3 | List of damaging missense URVs in *SLC6A1* and *GABRB3*.** Damaging missense (MPC≥2) URVs identified in *SLC6A1* and *GABRB3* with at least one epilepsy carrier; 'novel' indicates that the variant has not been previously reported. Coordinates are on GRCh38.

**Supplementary Data 4 | Results from exome-wide gene-set-based burden analysis of URVs. a**,**b**, Burden of protein-truncating (**a**) and damaging missense (MPC≥2; **b**) URVs in each gene set (gene family/protein complex) with at least one epilepsy or control carrier. For each variant class, burden analyses are performed across four epilepsy groups – 1,938 DEEs, 5,499 GGE, 9,219 NAFE, and 20,979 epilepsy-affected individuals combined ('EPI') – versus 33,444 controls. *P* values are computed using a Firth logistic regression model with adjustment for sex and ancestry.

**Supplementary Data 5 | List of protein-truncating URVs in GATOR1 genes.** Protein-truncating URVs identified in GATOR1-encoding genes (*DEPDC5*, *NPRL3*, and *NPRL2*) with at least one NAFE carrier; 'novel' indicates that the variant has not been previously reported. Coordinates are on GRCh38.

**Supplementary Data 6 | Results from burden analysis of $GABA_A$ receptor complex.** Burden of damaging missense (MPC≥2) URVs in the $(\alpha1)_2(\beta2)_2(\gamma2)$ $GABA_A$ receptor complex with respect to its structural domain; ECD: extracellular domain, TMD: transmembrane domain, TMD-2: the second TMD that forms the ion channel pore. For each domain, burden analyses are performed across three epilepsy groups – 1,938 DEEs, 5,499 GGE, and 9,219 NAFE – versus 33,444 controls. *P* values are computed using a Firth logistic regression model with adjustment for sex and ancestry.

**Supplementary Data 7 | Results from protein structural analysis of ion channel complexes.** ddG values of missense URVs across 16 ion channel protein complexes with experimentally resolved three-dimensional structures available ('PDB_ID'). A higher absolute ddG value suggests a more deleterious effect on protein stability; positive and negative values suggest destabilizing and stabilizing effects, respectively. Coordinates are on GRCh38.

**Supplementary Data 8 | Results from burden analysis of ion channel complexes by ddG.** Burden of damaging missense (MPC≥2) URVs in 16 ion channel protein complexes stratified by ddG. ddG≥1 and ddG≤-1 are applied to define destabilizing and stabilizing missense URVs, respectively; |ddG|≥1 comprises both. *P* values are computed using a Firth logistic regression model with adjustment for sex and ancestry.

**Supplementary Data 9 | Results from burden analysis of $GABA_A$ receptor complex by ddG.** Burden of (de)stabilizing missense ($|ddG| \geq 1$) URVs in the $(\alpha1)_2(\beta2)_2(\gamma2)$ $GABA_A$ receptor complex with respect to its structural domain; ECD: extracellular domain, TMD: transmembrane domain. ddG≥1 and ddG≤-1 are applied to define destabilizing and stabilizing missense URVs, respectively; $|ddG| \geq 1$ comprises both. *P* values are computed using a Firth logistic regression model with adjustment for sex and ancestry.

**Supplementary Data 10 | Results from exome-wide burden analysis of rare CNVs. a**-**d**, Genomic disorder (GD)-based burden of CNVs (**a**) and gene-based burden of CNV deletions (**b**), CNV deletions plus protein-truncating URVs (**c**), CNV duplications (**d**) with at least one epilepsy or control carrier. For each variant class, burden analyses are performed on the subset of samples that passed CNV calling QC, across four epilepsy groups – 1,743 DEEs, 4,980 GGE, 8,425 NAFE, and 18,963 epilepsy-affected individuals combined ('EPI') – versus 29,804 controls. *P* values are computed using a Firth logistic regression model with adjustment for sex and ancestry.

**Supplementary Data 11 | Results from burden analysis of GGE GWAS genes. a**, Gene-set-based burden of URVs in 23 genes implicated by GGE GWAS loci. Burden analyses are performed across four variant classes and two epilepsy groups – 5,499 GGE and 9,219 NAFE – versus 33,444 controls. **b**, Gene-based burden of protein-truncating URVs in 14 GGE GWAS genes with enrichment in GGE ('GGE_logOR'>0). *P* values are computed using a Firth logistic regression model with adjustment for sex and ancestry.

**Supplementary Data 12 | Results from functional analysis of candidate epilepsy genes. a**, List of candidate epilepsy genes with a prenatal (N=43) or postanal (N=50) expression bias in the human brain. Ten prenatal genes with a transcription factor function are indicated in 'prenatal_TF' and their regulatory targets overlapping with the postnatal genes are listed in 'postnatal_targets'. **b**,**c**, Gene Ontology (GO) terms enriched for the 43 prenatal and 50 postnatal genes (**b**) and all regulatory target genes linked to the 10 prenatal TFs (**c**). GO enrichment analysis are performed via the Gene Ontology Enrichment Analysis webserver (http://geneontology.org/).

**Supplementary Data 13 | Results from burden analysis of NDD genes. a,** Gene-set-based burden of URVs in genes implicated by WES of severe developmental disorders (DD; N=285), autism spectrum disorder (ASD; N=185), and schizophrenia (SCZ; N=32); and on the subsets of mutually exclusive genes (i.e., 196 DD-only, 99 ASD-only, and 22 SCZ-only genes). **b,** Gene-based burden of URVs in genes analyzed in **a**. In **a** and **b**, *P* values are computed using a Firth logistic regression model with adjustment for sex and ancestry. **c**, **d**, Protein-truncating and damaging missense (MPC≥2) variants identified in nine genes that are significant in both this and other NDD WES studies (**c**) and in *KDM6B* (**d**).

**Supplementary Data 14 | Results from ancestry-specific burden analysis of URVs. a**,**b**, Burden of protein-truncating (**a**) and damaging missense (MPC≥2; **b**) URVs in each protein-coding gene with at least one epilepsy or control carrier. **c**, Gene-set-based burden of URVs in established epilepsy genes (N=171 curated by the Genetic Epilepsy Syndromes [GMS] panel), constrained genes (N=1,917 scored by the loss-of-function observed/expected upper bound fraction [LOEUF] metric), and constrained genes excluding established epilepsy genes (N=1,813). Burden analyses are performed across six genetic ancestry groups – 16040/25641, 1598/2592, 480/3106, 1698/1215, 926/537, 237/353 case/control – of Non-Finnish European (NFE), African (AFR), Ad Mixed American (AMR), East Asian (EAS), Finnish (FIN), South Asian (SAS) samples, respectively. *P* values are computed using a Firth logistic regression model with adjustment for sex.

**Supplementary Data 15 | Results from sex-specific burden analysis of URVs. a**,**b**, Burden of protein-truncating (**a**) and damaging missense (MPC≥2; **b**) URVs in each protein-coding gene with at least one epilepsy or control carrier. **c**, Gene-set-based burden of URVs in established epilepsy genes (N=171 curated by the Genetic Epilepsy Syndromes [GMS] panel), X-linked GMS genes (N=37), constrained genes (N=1,917 scored by the loss-of-function observed/expected upper bound fraction [LOEUF] metric), and constrained genes excluding established epilepsy genes (N=1,813). Burden analyses are performed for female and male subgroups separately, across four epilepsy groups – 868/1070 DEEs, 3251/2248 GGE, 4818/4401 NAFE, and 11001/9978 all-epilepsy combined ('EPI') female/male cases – versus 18143/15301 female/male controls. *P* values are computed using a Firth logistic regression model with adjustment for ancestry.

### Supplementary Subjects and Methods

#### Details of individual participating Epi25 cohorts and funding sources

**Australia: Austin Hospital, Melbourne (AUSAUS)**

The Epilepsy Research Centre at the Austin Hospital in Melbourne, Australia, has been investigating the genetic basis of the epilepsies for over 30 years. The cohort in the Epi25 Collaborative were recruited to the epilepsy genetics research program over this period from the Austin Hospital, epilepsy clinics around Melbourne, and referrals from neurologists Australia-wide. Informed consent was obtained from patients or their parent/guardian as appropriate. DNA was extracted from blood or saliva samples. A skilled team of researchers and clinicians conducted detailed clinical phenotyping which involved a systematic review of medical records, including EEG and MRI reports, and a validated epilepsy questionnaire. Information on family history of seizures and other neurological disorders has also been collected via interviews with the patients and their families.

Most patients in the cohort are of European descent (‘Anglo-Australian’) although there is a diverse range of ethnic backgrounds including Asian, Middle Eastern, Indigenous Australian and mixed ethnicities. There is a known family history of seizures in 44% of the cohort (57% in the subset with GGE). The majority of the patients have had some previous genetic testing, including CNV testing and single gene testing. The DEE cohort have been extensively investigated with multiple iterations of a research panel of known, novel and putative genes for epilepsy. In addition, many patients with focal epilepsy have had a panel of known genes.

Funding and acknowledgements:
Collection of samples and phenotyping was supported by Australian NHMRC Program Grant (1091593 Berkovic, Scheffer), NHMRC Investigator Grants (1196637 Berkovic; 1172897 Scheffer) and NIH Grant (1R03NS108145-01 Lowenstein, Berkovic).

**Australia: Royal Melbourne and Alfred Hospitals (AUSRMB & AUSALF)**

The Royal Melbourne Hospital cohort, Melbourne, Australia, was prospectively recruited from the Epilepsy and First Seizure Clinics of The Epilepsy Program of the Royal Melbourne Hospital. Phenotypic information was obtained by direct interview, and review of the medical records, for the enrolled patients. Blood for DNA extraction was obtained on all participants and stored in the Biobank of the RMH Epilepsy Program, in the Department of Medicine, The Royal Melbourne Hospital, The University of Melbourne. Written informed consent was obtained for all participants, and the recruitment and study procedures were approved by the Human Research and Ethics Committee of Melbourne Health (The Royal Melbourne Hospital) - HREC #2002.232 & 2017.450. Funding sources: NHMRC Program, Project and Investigator Grants.

Relevant publications:

Speed D, Hoggart C, Petrovski S, et al. A genome-wide association study and biological pathway analysis of epilepsy prognosis in a prospective cohort of newly treated epilepsy. Hum Mol Genet. 2014;23(1):247-58. doi: 10.1093/hmg/ddt403. Epub 2013 Aug 19. PMID: 23962720; PMCID: PMC3857947.
Perucca P, Anderson A, Jazayeri D, et al…EpiPGX and EPIGEN Consortia. Antiepileptic Drug Teratogenicity and De Novo Genetic Variation Load. Ann Neurol. 2020;87(6):897-906. doi: 10.1002/ana.25724. Epub 2020 Apr 15. PMID: 32215971.

**Austria: Vienna (AUTMUV)**

The Austrian cohort consists of adult individuals diagnosed with epilepsy (mostly NAFE and GGE) that were recruited by the epilepsy monitoring unit or epilepsy outpatient clinic of our department. All patients gave written informed consent prior to inclusion, and the collection of data was approved by the local ethics committee (protocol number: 2051/2016). Epilepsy diagnosis was based on clinical information (as reported by the treating neurologist/epileptologist), routine and/or video EEG recordings and brain MRI scans. Relevant phenotype information was extracted from electronic medical records. DNA was extracted from peripheral blood using standard protocols.

**Belgium: Antwerp (BELATW)**

Patients were recruited by the VIB-Applied & Translational Neurogenomics Group of the University of Antwerp through epilepsy clinics at the different university hospitals in Belgium. All patients were diagnosed with a (so far) unexplained presumed genetic epilepsy and should have had at least 1 MRI of the brain excluding acquired causal lesions. The study was approved by the ethics committee of the University of Antwerp, and parents or the legal guardian of each proband signed an informed consent form for participation in the study. Genomic DNA of individuals was extracted from peripheral blood according to standard procedures. Clinical information was extracted from clinical files, as reported by their treating (paediatric) neurologists, and a subset was reviewed independently by two research team clinicians to ensure data quality and consistency. Sarah Weckhuysen receives funding from FWO-FKM (1861419N) and GSKE.

**Belgium: Brussels (BELULB)**

Adult patients with epilepsy were recruited consecutively through outpatient clinics and hospitalizations at Hôpital Erasme, Brussels, Belgium (between October 2004 and June 2020) and UZ Gasthuisberg, Leuven, Belgium (between October 2004 and June 2009). The study was approved by the Institutions' Review Boards. All patients provided written informed consent for data collection; patients with learning disability were included after consent from a parent or guardian. DNA was extracted from peripheral blood lymphocytes. Clinical information was collected from medical records and stored in a secured, web-based database.

Relevant publications:

Conte, F, Legros, B, Van Paesschen, W, et al. Long-term seizure outcomes in patients with drug resistant epilepsy. Seizure. 2018;62:74-78.

**Canada: Calgary (CANCAL)**

KMK, CC, SW and AS contributed 267 samples. The individuals were recruited within the Calgary Comprehensive Epilepsy Program mainly from the outpatient clinics and the seizure monitoring units at Foothills Medical Centre and South Health Campus, Calgary. All individuals were phenotyped in detail by epilepsy specialists (KMK, SW, AS). EEGs and MRIs were completed as part of the clinical workup. Final phenotypic classification and documentation of the phenotypic data in the study database were performed by KMK and CC. Individual selection and data export from the database for the study was done by KMK. DNA was extracted from peripheral blood or saliva. All individuals provided written informed consent.

**Canada: Toronto (CANUTN)**

There are 156 patients (51 DEE, 67 GGE, 18 NAFE, 20 Lesional Focal) in the Andrade cohort, typically from the Greater Toronto region of Ontario, Canada. They are mostly of Caucasian ancestry, but also African, South Asian, East Asian, Latino, Middle Eastern, Jewish and Indigenous. Patients were recruited to each group through an REB protocol allowing for the collection of blood or saliva and data collaboration.

Patients that were previously consented were re-consented to allow for Whole Exome Sequencing and data sharing with the EPI25 group. After collection, the sample was de-identified, and then extracted and stored at the Hospital for Sick Children, Toronto, Canada.

**Switzerland: Bern (CHEUBB)**

In the recruitment of our cohort, the Departments of Neurology and BioMedical Research, Bern University Hospital and University of Bern, Bern, Switzerland, and the Institute of Human Genetics, Bern University Hospital, Bern, Switzerland, were involved. The Swiss study population encompasses > 90 patients (28 in year 2, 70 in year 4) with epilepsy between 2 and 63 years of age. All patients have been de-identified for the Epi25 Study. The patient ascertainment protocol was according to Epi25 phenotyping requirements. Phenotyping information was taken from medical records, stored in the hospital's database, and entered in de-identified form into a RedCap database provided by Ep25. DNA source was patients' venous blood. DNA was extracted with standard kits at the Institutes of Human Genetics or Clinical Chemistry of Bern University Hospital and stored there at -80 degrees C. Informed consent declarations are available from all patients and have been approved by the American Institutional Review Board involved in the Epi25 Study. The Cantonal Ethics Committee Bern, Switzerland, granted permission for participation of Bern University Hospital and University of Bern in the Epi25 Study including all steps described above.

**Cyprus (CYPCYP)**

Epilepsy-affected subjects of the Cyprus cohort were recruited and enrolled in the Epi25 Consortium by physicians during routine clinical visits in the Cyprus Institute of Neurology and Genetics. Phenotypic data were collected at the time of enrolment and submitted into the Epi25 REDCap database in a de-identified manner. There are 152 unrelated individuals of Southern European ancestry in the Cyprus cohort; 63 GGE subjects, 62 NAFE, 14 Lesional Focal Epilepsy and 13 DEE. All subjects had clinical, neuroimaging and EEG characteristics meeting the International League against Epilepsy (ILAE) 2017 Seizure Classification criteria. A control Cyprus cohort of 32 individuals of Southern European ancestry with no epilepsy or other neuropsychiatric phenotypes was also included. Genomic DNA samples were extracted from whole blood with the Gentra Puregene Blood Kit (Qiagen, Hilden, Germany) according to the manufacturer's guidelines. This study was carried out in compliance with the Cyprus National Bioethics Committee (ΕΕΒΚ/ΕΠ/2015/22) and written informed consent was obtained from all study participants or their legal guardians.

**Czech Republic: Prague (CZEMTH)**

Patients in our cohort have been diagnosed with West syndrome, myoclonic-astatic epilepsy or epileptic encephalopathy of unknown aetiology. Brain magnetic resonance imaging and metabolic screening excluded any underlying pathology. Patients were collected at the Department of Child Neurology of the 2nd Medical Faculty and University Hospital Motol. Legal guardians of patients signed an informed consent. The study was approved by the local ethics committee. Supported by the Ministry of Health of Czech Republic AZV 15–33041 and DRO 00064203.

**Germany: Frankfurt/Marburg (DEUPUM)**

The individuals were recruited from the outpatient clinics and the video EEG monitoring units at the Epilepsy Centers Frankfurt Rhine-Main and Hessen-Marburg. All individuals were phenotyped in detail by epilepsy specialists (KMK, FR, SK, PSR, AS, FZ, SVB) within the EpimiRNA project (European Union's 'Seventh Framework' Programme (FP7) under Grant Agreement no. 602130). EEGs and MRIs were performed as part of the clinical workup. Phenotypic classification and data entry for the biobank for paroxysmal neurological disorders was performed by KMK, FR, PSR, FZ and SVB. Individual selection and data export from the biobank for the study was performed by KMK, PSR, FZ and SVB. DNA was extracted from peripheral blood or saliva. All individuals provided written informed consent.

**Germany: Giessen (DEUUGS)**

Diagnosis of Rolandic Epilepsy was performed according to the International Classification of Seizures and Epilepsies as described. Sleep activation, characteristic shape, and classification by two independent individuals were required for classification of the EEG trait. Atypical benign partial epilepsy of childhood (ABPE) was diagnosed employing the following criteria: Characteristic EEG trait of CTS, however, with trains of continuous generalized nocturnal discharges as a prerequisite of diagnosis in all ABPE cases. In addition at least one of the following two features needed to be present: (1) seizures and EEG trait compatible with BECTS plus one or more additive seizure types like astatic seizures, atypical absences ("dreamy states") or myoclonic seizures as reported. MF is a member of ERN EpiCARE.

Relevant publications:

Bobbili DR, Lal D, May P, et al.; EUROEPINOMICS COGIE Consortium. Exome-wide analysis of mutational burden in patients with typical and atypical Rolandic epilepsy. Eur J Hum Genet. 2018;26(2):258-264.

**Germany: University of Bonn, Bonn (DEUUKB)**

The sample recruitment site is the Department of Epileptology at the University of Bonn. The collection of 2,588 blood DNA samples from patients with epilepsy which were included in the present study was conducted from 2007 till 2015 within the projects Epicure (Functional Genomics in Neurobiology of Epilepsy: A Basis for New Therapeutic Strategies) and NGEN-Plus (Genetic basis of Levetiracetam pharmacoresistance and side effects in human epilepsy) and has been approved by the Ethics committee of University Bonn Medical Center (approval code: 040/07). Genomic DNA was isolated from 10 ml aliquots of EDTA-anticoagulated blood by a salting-out technique. From selected samples of this cohort GWAS data have been published in several studies.

Funding/acknowledgements:

Funding for the DEUUKB cohort was obtained from European Union (FP7 project EpiPGX, grant 279062 to WSK).

Relevant publications:

Miller SA, Dykes DD, Polesky HF. A simple salting out procedure for extracting DNA from human nucleated cells. Nucleic Acids Res. 1988;16:1215.

Brainstorm C, Anttila V, Bulik-Sullivan B, et al. Analysis of shared heritability in common disorders of the brain. Science. 2018;360:eaap8757.

McCormack M, Gui H, Ingason A, et al. Genetic variation in CFH predicts phenytoin-induced maculopapular exanthema in European-descent patients. Neurology. 2018;90:e332-e341.

Lal D, Ruppert AK, Trucks H, et al. Burden analysis of rare microdeletions suggests a strong impact of neurodevelopmental genes in genetic generalised epilepsies. PLoS Genet. 2015;11:e1005226.

**Germany: Kiel (DEUUKL)**

Patients were recruited by the Neuropedriatics Group of the University Hospital of Schleswig-Holstein and through the Israeli-Palestinian Family Consortium. The recruitment and analysis of these samples is covered by the Kiel IRB. Patients, their parents or the legal guardian of each proband signed an informed consent form for participation in the study. Clinical data was collected from clinical files and a subset of patients from Israel or Palestine was interviewed by a research team of clinicians to provide their clinical data. Genomic DNA of patients was extracted from peripheral blood according to standard procedures.

**Germany: Leipzig (DEUULG)**

Patients were recruited by the Swiss Epilepsy Center in Zurich, Switzerland and samples were transferred for research and storage to the Institute of Human Genetics at the University of Leipzig, Germany. All patients were diagnosed with temporal lobe epilepsy due to an indicative EEG. Most patients had at least 1 MRI of the brain with focus on focal abnormalities, especially of the temporal lobe / hippocampal structures. The study was approved by the "Kantonale Ethikkommission Zürich". Parents or the legal guardian of each proband signed an informed consent form for participation in research studies including whole genome analyses. Genomic DNA of individuals was extracted from peripheral blood according to standard procedures. Clinical information was extracted from clinical files, as reported by their treating neurologists.

**Germany: Tuebingen (DEUUTB)**

Our study cohort consists of more than 1 500 samples with mainly caucasian origin. These samples were recruited at Tubingen and 38 other cooperating departments of neurology from university clinics and outpatient clinics in Germany. The Ethics / informed consent was approved by the ethics committee of the Medical Faculty of the Eberhard-Karls University and at the University Hospital Tubingen. Patients with GGE, DEE, NAFE and some structural epilepsies were systematically recruited in outpatient clinics of university and other hospitals, and from neurological practices by a letter of invitation sent to patients. Retrospective data from medical reports of epileptologists were used. If deemed necessary, personal interviews of patients were undertaken. The DNA source was blood. There is no single publication describing the whole sample. Typical publications including part of these samples are from the following consortia: Epicure, EuroEPINOMICS, EpiPGX, ILAE consortium on the genetics of complex epilepsies, Epi25.

Funding/acknowledgements:

Funding for the DEUUTB cohort was obtained from the German Research Foundation (DFG) and the Fond Nationale de la Recherche (FNR) in Luxembourg in the frame of the Research Unit FOR-2715 (grants Le1030/14-1 & /23-1, Le1030/16-1 & /16-2, We4896/4-1 & /4-2, He5415/7-1 & /7-2, INTER/DFG/21/16394868 MechEPI2), an additional DFG grant (DFG: WE4896/3-1), by the rare disease program of the Federal Ministry of Education and Research (BMBF, Treat-ION, grants 01GM1907A, 01GM2210A & 01GM2210B), by the German Society for Epileptology, and by the foundation no-epilep. This research was part funded by Science Foundation Ireland (SFI) under Grant Number 16/RC/3948 and co-funded under the European Regional Development Fund and by FutureNeuro industry partners

**Finland: Kuopio (FINKPH)**

Patients diagnosed with epilepsy and visiting Epilepsy Center, Kuopio University Hospital (KUH), Finland have given their written informed consent to record their clinical data to an epilepsy research registry of KUH and University of Eastern Finland and collect a blood sample for DNA analysis. Consent was collected from the legal quardian, if applicable. The ethics committee of KUH has approved the study.

**Finland: Helsinki (FINUVH)**

Patients were recruited at the University of Helsinki through pediatric epilepsy clinics in Helsinki and Tampere University Hospitals in Finland. All patients were diagnosed with a presumed genetic epilepsy, the etiology remaining unknown. All patients had an MRI done to exclude acquired causal lesions. This cohort included 95 patients: 26 patients with developmental and epileptic encephalopathy (DEE), 51 patients with genetic generalized epilepsy (GGE) and 18 patients with non-acquired focal epilepsy (NAFE). Clinical phenotyping involved a systematic review of medical records, including EEG and MRI reports. The study was approved by an ethics committee of The Hospital District of Helsinki and Uusimaa, Finland. The parents or the legal guardian of each proband signed an informed consent form for participation in the study. Genomic DNA of the patients was extracted from peripheral blood or saliva according to standard procedures.

**France: Lyon (FRALYU)**

The REPO2MSE study is a multicenter prospective study, based on the French National Research Network on SUDEP predictors, which was approved by ethics committee (CPP Sud Est II n°2010-006-AM6) and competent authority (ANSM n° B100108-40). Its primary objective is to individualize risk factors of SUDEP in patients suffering from drug-resistant focal epilepsy. 1069 Adult patients (age ≥16 years) with drug-resistant focal epilepsy according to ILAE classifications who underwent long term monitoring using either video scalp EEG or intracranial EEG recordings and who gave written informed consent were recruited in 16 French epilepsy monitoring units. For all included patients, we collected demographic and detailed clinical data, MRI data, inter-ictal EEG data, results of non-systematic complementary investigations performed to better localize the epileptogenic zone (i.e 18FDG PET, ictal SPECT) and raw data of all recorded seizures, which include EEG, video, pulse oximetry and EKG. For patients who gave specific consent, we also collected blood samples for genetic analyses which were centrally stored in the Department of Clinical Genetics at Hospices Civils de Lyon. All patients then received specific information about the collaboration between the REPOMSE study and the EPI25 project. Overall, 810 patients from twelve participating centers confirmed their consent for transmission of their blood samples to the Broad Institute.

Relevant publications:

Alexandre V, Mercedes B, Valton L, et al. Risk factors of postictal generalized EEG suppression in generalized convulsive seizures. Neurology. 2015;85:1598-603.

Rheims S, Alvarez BM, Alexandre V, et al. Hypoxemia following generalized convulsive seizures: Risk factors and effect of oxygen therapy. Neurology. 2019;92:e183-93.

**New Wales: Swansea (GBRSWU)**

The samples from Wales: Swansea are part of the Swansea Neurology Biobank (SNB). The SNB has been approved by the Welsh Research Ethics Committee (REC 17/WA/0290). Participants are recruited into the biobank, with written informed consent or assent, from regional National Health Service (NHS) neurology and epilepsy clinics. Participants provide written consent to share their clinical and genetic information anonymously with ethical research collaborations. Participants' medical records (including EEG and MRI results and epilepsy clinic letters) are reviewed by the research and clinical team (which include experienced epileptologists) to confirm diagnosis. SNB participants blood samples are sent to the UK Porton Down ECACC facility for DNA extraction and the DNA is then returned to be stored securely at Swansea University. For epi25 we have submitted approximately 310 bio-samples and patient records in Yrs. 1-5.

**UK: UCL (GBRUCL)**

Participant recruitment took place at the National Hospital for Neurology and Neurosurgery (United Kingdom). Written informed consent or assent was obtained between 10/01/2000 and 01/25/2015 from all participants according to local and national requirements and blood samples were collected for DNA extraction. 709 epilepsy cases were submitted for analysis. Allocation to the following groups was based on the clinical diagnosis and the specific inclusion and exclusion criteria of the Epi25 consortium: generalized genetic epilepsy (n=393, 145 male), non-acquired focal epilepsy (n=313, 146 male), developmental and epileptic encephalopathy (n=3, 2 male). Additionally, relatives were included, where samples were available (n=3, 2 male). Phenotypic information was obtained from local medical records by clinical or trained non-clinical researchers.

Funding/acknowledgments:
The GBRUCL cohort thanks the Epilepsy Society and Muir Maxwell Trust for funding support. The work conducted at GBRUCL was undertaken at University College London Hospitals, which received a proportion of funding from the NIHR Biomedical Research Centres funding scheme.

**UK: Imperial/Liverpool (GBRUNL)**

GBRUNL samples are derived from four separate, UK-wide, ethically approved studies coordinated by the University of Liverpool (UK) and Imperial College London (UK). The SANAD and MESS linked DNA Bank and Relational Database study recruited individuals with newly-diagnosed focal, generalised or unclassified epilepsy from out-patient neurology clinics between 2003-2006 [Leschziner et al, 2006; Speed et al, 2014]. The Pharmacogenetics of GABAergic Mechanisms of Benefit and Harm in Epilepsy study recruited individuals with refractory focal epilepsy, previously or prospectively exposed to adjunctive treatment with clobazam or vigabatrin, from out-patient neurology clinics between 2005-2009. The Refractory Juvenile Myoclonic Epilepsy Cohort (ReJuMEC) study recruited individuals with valproic acid resistant juvenile myoclonic epilepsy from out-patient neurology clinics between 2009 and 2010. The Standard and New Antiepileptic Drugs (SANAD-II) study recruited individuals with newly-diagnosed focal, generalised or unclassified epilepsy from out-patient neurology clinics between 2013-2019. In all cases, study participants provided written informed consent to the collection (via blood or saliva sampling) and analysis of their DNA for use in genetic and pharmacogenetic research related to epilepsy and its treatment. All studies were approved by research ethics committees in operation at the relevant time (SANAD DNA bank, North West MREC ref 02/8/45; GABAergic mechanisms, UCLH REC ref 04/Q0505/95; ReJuMEC, Cheshire REC ref 09/H1017/55; SANAD-II, North West REC ref 12/NW/0361).

Relevant publications:

Speed D, Hoggart C, Petrovski S, et al. A genome-wide association study and biological pathway analysis of epilepsy prognosis in a prospective cohort of newly treated epilepsy. Hum Mol Genet. 2014;23:247-258.

Leschziner G, Jorgensen AL, Andrew T, et al. Clinical factors and ABCB1 polymorphisms in prediction of antiepileptic drug response: a prospective cohort study. Lancet Neurol. 2006;5:668-676.

**Hong Kong (HKGHKK)**

Epilepsy patients of Han Chinese ethnicity aged between 2 and 91 years were recruited from neurology clinics of five regional hospitals in Hong Kong covering a combined catchment population of approximately 3 million. Syndromic classification was adapted from the revised international organization of phenotypes in epilepsy. DNA was extracted from venous blood. The study was approved by ethics committees of the participating hospitals, and all patients or their legal guardians gave written informed consent. The sample collection methodology has been described in Guo Y, Baum LW, Sham PC, et al. Two-stage genome-wide association study identifies variants in CAMSAP1L1 as susceptibility loci for epilepsy in Chinese. Hum Mol Genet. 2012;21:1184-1189.

Funding/acknowledgments:
Research Grants Council of the Hong Kong Special Administrative Region, China (project number CUHK4466/06M).

**Ireland: Dublin (IRLRCI)**

Patients were all adults and recruited from a specialized epilepsy clinic at Beaumont Hospital, Dublin, Ireland. Patients were mostly of Irish ethnicity. This study was approved by the Beaumont Hospital Ethics Committee.

**Italy: Milan (ITAICB)**

Our cohort included the DNA samples of 352 individuals with epilepsy and 62 controls (healthy subjects not related with the individuals with epilepsy, and without epilepsy history). The population included individuals with generalized epilepsy (GGE), individuals with developmental and epileptic encephalopathy (DEE), individuals with focal epilepsy due to cerebral malformations (mainly nodular heterotopia), and individuals with nonacquired focal epilepsy (NAFE). All of the individuals were diagnosed and followed at our Institute. Diagnosis of epilepsy was based on clinical, EEG and neurophysiological data, neuroimaging (MRI). Metabolic screening, karyotype, CGH array, analyses of single genes and customized panels were performed in some cases, when appropriate. The individuals did not undergo exome sequencing analysis (before the Epi25 collection). The DNA of the individuals was extracted from peripheral blood, according to standard procedures, after signature of an Informed Consent form. The genetic study was approved by The Ethic Committee of our Institute. No publications have described genetic findings pertaining to the collected individuals until now. Clinical information was extracted from clinical files, as reported by their treating (paediatric and adult) neurologists.

**Italy: Genova (ITAIGI)**

Patients with generalized and focal epilepsy or developmental epileptic encephalopathy referred to ‘IRCCS G. Gaslini Institute’. The study was approved by the IRB and written informed consent was signed by the patients/parents. Clinical information, including data on EEG and antiepileptic therapy, were recorded on data collection forms. Genomic DNA isolation and genetic analysis was carried out with the Nimblegen-SeqCapEZ-V244M enrichment kit on the Illumina HiSeq2000 system.

Relevant publications:

Wolking S, Campbell C, Stapleton C, et al. Role of Common Genetic Variants for Drug-Resistance to Specific Anti-Seizure Medications. Front Pharmacol. 2021;12:688386

Muir AM, Myers CT, Nguyen NT, et al. Genetic heterogeneity in infantile spasms. Epilepsy Res. 2019;156:106181.

Leu C, Stevelink R, Smith AW, et al. Polygenic burden in focal and generalized epilepsies. Brain. 2019;142:3473-3481.

Wolking S, Moreau C, Nies AT, et al. Testing association of rare genetic variants with resistance to three common antiseizure medications. Epilepsia. 2020;61:657-666.

Accogli A, Severino M, Riva A, et al. Targeted re-sequencing in malformations of cortical development: genotype-phenotype correlations. Seizure. 2020;80:145-152.

Funding/acknowledgements:

'Research supported by Project RIN - RCR-2022-23682290 and PNRR-MUR-M4C2 PE0000006 Research Program “MNESYS”—A multiscale integrated approach to the study of the nervous system in health and disease. IRCCS ‘G. Gaslini’ is a member of ERN-Epicare

**Italy: Bologna (ITAUBG)**

610 unrelated patients with epilepsy of uncertain aetiology were consecutively recruited by the adult and pediatric neurologists of the IRCCS Istituto delle Scienze Neurologiche di Bologna. The cohort included patients with developmental and/or epileptic encephalopathy (n=167), genetic generalized epilepsy (n=167), and non-acquired focal epilepsy with or without brain lesion (n=337). All patients underwent EEG

and neuroimaging (CT and/or MRI). Clinical information collected from medical records have been revised and entered by expert neurologists in a pseudonymized manner into the REDCap database, provided by the Epi25 collaborative. The local ethics committee approved the study (CE:16057) and specific consent was obtained from all study participants or their parents/legal guardians, as appropriate. Genomic DNA of patients was extracted from peripheral blood according to standard procedures.

Relevant publications:
Licchetta L, Bisulli F, Vignatelli L, et al. Sleep-related hypermotor epilepsy: Long-term outcome in a large cohort. Neurology. 2017;88(1):70-77. doi: 10.1212/WNL.0000000000003459
Bisulli F, Menghi V, Vignatelli L, et al. Epilepsy with auditory features: Long-term outcome and predictors of terminal remission. Epilepsia. 2018;59(4):834-843. doi: 10.1111/epi.14033
Licchetta L, Pippucci T, Baldassari S, et al.; Collaborative Group of Italian League Against Epilepsy (LICE) Genetic Study Group on SHE. Sleep-related hypermotor epilepsy (SHE): Contribution of known genes in 103 patients. Seizure. 2020;74:60-64. doi: 10.1016/j.seizure.2019.11.009
Minardi R, Licchetta L, Baroni MC, et al. Whole-exome sequencing in adult patients with developmental and epileptic encephalopathy: It is never too late. Clin Genet. 2020;98(5):477-485. doi: 10.1111/cge.13823
Bisulli F, Rinaldi C, Pippucci T, et al. Epilepsy with auditory features: Contribution of known genes in 112 patients. Seizure. 2021;85:115-118. doi: 10.1016/j.seizure.2020.12.015.
Gozzelino L, Kochlamazashvili G, Baldassari S, et al. Defective lipid signalling caused by mutations in PIK3C2B underlies focal epilepsy. Brain. 2022;145(7):2313-2331. doi: 10.1093/brain/awac082. PMID: 35786744; PMCID: PMC9337808.

**Italy: Catanzaro (ITAUMC)**
Patients were recruited by the Epilepsy Group of the University Magna Graecia of Catanzaro (Italy) that includes a Pediatric and Adult Neurologic Unit with a specific focus on genetic epilepsy. In each patient, the diagnosis of epilepsy syndrome is based on comprehensive clinical, neuropsychological, electroencephalographic, and MR evaluations. Clinical data are stored into a database. The study was approved by the ethics committee of the University of Catanzaro Italy, and parents or the legal guardian of each proband signed an informed consent form for participation in the study. Genomic DNA of individuals was extracted from peripheral blood according to standard procedures.

**Italy: Florence (ITAUMR)**
Individuals were studied at the Clinical Neurology Unit and Neurogenetics lab of the Neuroscience Department of Meyer Children's Hospital-University of Florence. All individuals were diagnosed with an unexplained presumed genetic epilepsy. The study was approved by the Pediatric Ethics Committee of the Tuscany Region. Parents or the legal guardians of each proband signed an informed consent form for participation in the study. Genomic DNA of individuals was extracted from peripheral blood according to standard procedures. Clinical information was extracted from clinical files, as reported by the treating neurologists.

**Japan: Fukuoka (JPNFKA)**
Children with mostly intractable epilepsies. Their ethnicity is exclusively Japanese.

**Japan: RIKEN Institute (JPNRKI)**
Japanese patients with epilepsies were recruited by National Epilepsy Center, Shizuoka Institute of Epilepsy and Neurological Disorder. Epileptic seizures and epilepsy syndrome diagnoses were performed according to the International League Against Epilepsy classification of epileptic syndromes. Genomic DNA was extracted from peripheral venous blood samples using QIAamp DNA Blood Midi Kit according to the

manufacturer's protocol (Qiagen). The experimental protocols were approved by the Ethical Committee of RIKEN Institution and National Epilepsy Center, Shizuoka Institute of Epilepsy and Neurological Disorder. Written informed consent was obtained from all individuals and/or their families in compliance with the relevant Japanese regulations.

**Lebanon: Beirut (LEBABM)**

During years 1-5, the American University of Beirut Medical Center contributed a total of 1358 DNA samples with their corresponding phenotypes to the Epi25 project . All recruited individuals participated in an ongoing centralized study evaluating the electroclinical syndromes of children and adults with new onset unprovoked seizures or epilepsy in Lebanon, which so far enrolled more than 4,000 individuals. As part of the protocol, all individuals were evaluated with a dedicated seizure protocol MRI and a 3-hour sleep deprived video/EEG. A team of epileptologists conducted detailed phenotyping based on a detailed history of the semiology of the events in addition to EEG and MRI findings. The study was approved by the Institutional Review Board of the American University of Beirut Medical Center and all individuals or parents/guardians signed an informed consent form. Genomic DNA of the individuals was extracted from peripheral blood according to standard procedures.

Relevant publications:

El Halabi T, Dirani M, Nasreddine W, et al. The importance of acknowledging diagnostic uncertainty in patients with new-onset paroxysmal spells. Epilepsia Open. 2021;6(4):727-735. doi: 10.1002/epi4.12544.

Hourani R, Nasreddine W, Dirani M, et al. When Should a Brain MRI Be Performed in Children with New-Onset Seizures? Results of a Large Prospective Trial. AJNR Am J Neuroradiol. 2021;42(9):1695-1701. doi: 10.3174/ajnr.A7193.

Nawfal O, Nasreddine W, Hmaimess G, et al. Depression and anxiety in patients from Lebanon with new onset functional seizures. Seizure. 2021;88:22-28. doi: 10.1016/j.seizure.2021.03.014.

Nasreddine W, Fakhredin M, Makke Y, et al. Hyperventilation-induced high-amplitude rhythmic slowing: A mimicker of absence seizures in children. Epilepsy Behav. 2020;103:106510.

Arabi M, Dirani M, Hourani R, et al. Frequency and Stratification of Epileptogenic Lesions in Elderly With New Onset Seizures. Front Neurol. 2018;9:995.

**Lithuania (LTUUHK)**

Patients were recruited in Vilnius University Hospital Santaros Klinikos by a clinical geneticist through a referral of a neurologist or a child neurologist and according to inclusion/ exclusion criteria. The study was approved by Institution’s Research Ethics Committee, and each proband or parents/ legal guardians of a proband signed an informed consent. Samples of genomic DNA were obtained during the routine procedure for blood sampling for genetic testing done in a clinical testing and the majority of patients had chromosomal microarray, metabolic testing and/or gene/gene panel testing prior to the inclusion into the study. Clinical information was extracted from clinical files and obtained during the clinical genetic consultation.

**New Zealand: Otago (NZLUTO)**

The Epilepsy Research Group at the University of Otago, Wellington, New Zealand has been undertaking epilepsy genetic research for over 15 years. The Epi25 cohort consisted of individuals recruited as part of this larger project. Individuals were referred from neurology, paediatric and genetic outpatient services throughout New Zealand. Detailed clinical phenotyping was performed by trained researchers and paediatric neurologists following interviews and reviews of the medical records, EEGs and MRIs. Information on family history was obtained via interviews with the proband and their family. Participants were from the following ethnic groups: New Zealand European, Māori, Pacific Peoples, Asian, Hispanic,

Ethiopian, Middle Eastern. The study protocol was approved by the New Zealand Health and Disability Ethics Committee. Participants and their parent/guardian if appropriate, gave written informed consent for clinical and genetic analysis. DNA was extracted from blood or saliva. Individuals in the GGE and NAFE cohort had no prior genetic testing. The majority of individuals in the DEE cohort had CNV testing but no other genetic investigations.

Funding/acknowledgements:
Cure Kids New Zealand and Health Research Council of New Zealand

**SEEDS cohorts Africa (KENKIL, GHAKNT, ZAFAGN)**
People with epilepsy were recruited from Demographic surveillance sites of three sites in Africa namely: Agincourt, South Africa; Kilifi, Kenya and Kintampo, Ghana. All people with epilepsy were first identified through a community survey, and diagnosis confirmed by epilepsy clinicians and neurologists using medical history, neurologic examination, electroencephalography and magnetic resonance imaging data. Seizure types, age of onset, neurological examinations, past and family history, and features of electroencephalography and neuroimaging were evaluated. The study population consisted of people with genetic generalized epilepsies, lesional focal epilepsies and non-acquired focal epilepsies. Ethics committee approval was obtained from the Scientific and Ethics Review Unit of Kenya Medical Research Institute. Peripheral blood samples were collected from people with epilepsy and age-group and sex matched controls, and stored for future assays following written informed consent. DNA extraction was performed in the laboratories of the KEMRI-Wellcome Trust Research Programme in Kenya and shipped to the Broad Institute for genotyping and exome sequencing.

Relevant publications:
Kariuki SM, Matuja W, Akpalu A, et al. Clinical features, proximate causes, and consequences of active convulsive epilepsy in Africa. Epilepsia. 2014;55(1):76-85. doi: 10.1111/epi.12392. PMID: 24116877; PMCID: PMC4074306.

**Turkey: Bogazici (TURBZU)**
Patients were recruited by the Child Neurology and Neurology clinics at the different university hospitals in Turkey. The study was approved by the Institutional Review Board for Research with Human Subjects (INAREK) of Boğaziçi University, and parents or the legal guardian of each proband signed an informed consent form for participation in the study. Genomic DNA of individuals was extracted from peripheral blood according to standard procedures. Clinical information was reported by their treating (pediatric) neurologists. The cohort included a total of 171 patients (128 EE, 28 GGE and 15 Focal epilepsy patients) and 39 healthy controls. All epileptic encephalopathy patients had severe epilepsy, with developmental delay and regression, normal neuroimaging and epileptiform activity on EEG. Healthy control group included individuals with no symptoms of any neurological disorder.

**Turkey: Istanbul (TURIBU)**
Epilepsy patients were recruited from Epilepsy Clinics of Department of Neurology, Istanbul Faculty of Medicine, Istanbul University and Cerrahpaşa Medical Faculty , Istanbul University-Cerrahpaşa. The study population consisted of patients with idiopathic/genetic generalized epilepsies, lesional or non-lesional focal epilepsies and epileptic encephalopathies, including sporadic and familial cases. All patients were long-term follow-up. Seizure types, age of onset, neurological examinations, past and family history, prognosis and response to treatment, features of electroencephalography and neuroimaging were evaluated. Ethics committee approval was obtained. Peripheral blood samples were collected from all

patients following written informed consent. DNA isolation was performed in the Department of Genetics, Aziz Sancar Institute of Experimental Medicine, Istanbul University.

**USA: Boston Children's Hospital (USABCH)**

USA: Boston Children's Hospital, Boston (USABCH) Cases from Boston Children's Hospital (BCH) were ascertained from 3 local repositories. All repository protocols are approved by the BCH Institutional Review Board and participants were consented under one (or more) of the following protocols. The Genetics of Epilepsy and Related Disorders protocol (PI Dr. Annapurna Poduri) enrolls individuals with a clinical epilepsy diagnosis for genotype/phenotype correlation. Samples are obtained from BCH and non-BCH individuals and biological samples collected for genetic sequencing. Individual medical records (BCH and outside records) are reviewed for phenotyping purposes. The Phenotyping and Banking Repository of Neurological Disorders (PI Dr. Mustafa Sahin) enrolls individuals seen at BCH with any neurological phenotype, including epilepsy. Dr. Poduri is on the Steering Committee for the Core for Neurological Diseases and has requested samples from this source for Epi25. Boston Children's Biobank for Health Discovery (PI Dr. Kenneth Mandl) enrols any individual of BCH, regardless of diagnosis or phenotype. Dr. Poduri serves on the Data and Sample Access Committee for this repository and has requested samples from this source for Epi25. Samples from the two latter repositories are available to BCH researchers through an application process, which includes confirmation of an IRB-approved protocol. Individuals with a clinical diagnosis of epilepsy were reviewed for Epi25 eligibility using their BCH medical records. Dr. Poduri and Rebecca Pinsky have been involved in ascertaining and coordinating the phenotyping of all USABCH Epi25 cases.

**USA: Baylor College of Medicine (USABLC)**

Healthy controls and patients with genetic epilepsies.

**USA: Cleveland Clinic (USACCF)**

Patients were recruited through the Cleveland Clinic Epilepsy Center. All patients had routine EEG and/or video-EEG monitoring and had been diagnosed with epilepsy. The study was approved by the Cleveland Clinic Institutional Review Board, and all participants (or their guardian/legally authorized representative) provided informed consent for study participation. Genomic DNA of individuals was extracted from peripheral blood according to standard procedures. Clinical information was extracted from electronic health records.

**USA: Cincinnati Children's Hospital Medical Center (USACCH)**

The samples were from subjects involved with the NIH funded 32 center Childhood Absence Epilepsy clinical trial (ClinicalTrials.gov Identifier: NCT00088452). The subjects were children between 2.5 and 13 years old with newly diagnosed, EEG proven absence seizures who met ILAE criteria for Childhood Absence Epilepsy. Blood was obtained at the first treatment visit for DNA isolation. All subjects (or their parents/guardians) signed written informed consent permitting DNA isolation, storage, and pharmacogenetic analysis. Those informed consents allowing for sharing and broader genetic analysis were shared with Epi25K. For more details please see:

Glauser TA, Holland K, O'Brien VP, et al. Pharmacogenetics of antiepileptic drug efficacy in childhood absence epilepsy. Annals of Neurology. 2017;81(3):444 DOI: 10.1002/ana.24886.

**USA: Philadelphia/CHOP/Thomas Jefferson University (USACHP) and Philadelphia/ Thomas Jefferson University/Rowan (USACRW)**

The Philadelphia Cohort began in 1997 and collected blood, saliva and brain tissues from patients with common forms of idiopathic human epilepsy, mostly genetic generalized epilepsy (GGE) and non acquired focal epilepsy (NAFE). The collection began at Thomas Jefferson University Hospital in Philadelphia and expanded to include six other sites: The Children's Hospital of Philadelphia, The University of Pennsylvania, The University of Cincinnati, Nationwide Children's Hospital, Beth Israel Deaconess and The University of Montreal. The cohort consists of more than 4000 samples from epilepsy patients collected and supported during two periods of NIH funding (R01NS493060, 2001-2007 RJ Buono PI and R01NS06415401, 2009-2012 RJ Buono and H Hakonarson Co- PI). Over 1000 additional samples from first degree relatives of the patients were also collected. Many of these samples are available to the research community via the NINDS sample repository at the Coriell Institute in Camden NJ. All studies were approved by Institutional Review Boards at each participating site. All patients were identified and recruited by trained epileptologists at tertiary care centers using inclusion and exclusion criteria previously published. Diagnostic methods applied included EEG, MRI, and collection of deep phenotypic information on family history, medications, risk factors, age of onset, and other information. For the Epi25K project, blood and saliva were used as the source of DNA.

Relevant publications:
Buono RJ, Lohoff F, Sander T, et al. Association Between Variation in the Human KCNJ10 Potassium Ion Channel Gene and Seizure Susceptibility. Epilepsy Research. 2004;58:175-183.
International League Against Epilepsy Consortium on Complex Epilepsies. Genetic determinants of common epilepsies: a meta-analysis of genome-wide association studies. Lancet Neurol. 2014;13(9):893-903. doi: 10.1016/S1474-4422(14)70171-1.

Funding/acknowledgements:
NIH funding (R01NS493060, 2001-2007 RJ Buono PI and R01NS06415401, 2009-2012 RJ Buono and H Hakonarson Co- PI). U01HG006830 (PI Hakonarson) and The Children’s Hospital of Philadelphia Endowed Chair in Genomic Research (PI Hakonarson)

**USA: EPGP (USAEGP)**
Infantile spasms (IS), Lennox–Gastaut syndrome (LGS), genetic generalized epilepsy (GGE), and non-acquired focal epilepsy (NAFE) patients were collected through the Epilepsy Phenome/Genome Project (EPGP, http://www.epgp.org). More than 4,000 participants in EPGP were enrolled across 27 clinical sites from around the world. The subset of samples included in Epi25 were enrolled from 20 sites across the USA and in Australia. IS patients were required to have hypsarrhythmia or a hypsarrhythmia variant on EEG. LGS patients were required to have EEG background slowing or disorganization for age and generalized spike and wave activity of any frequency or generalized paroxysmal fast activity (GPFA). IS and LGS cases were enrolled as trios with both biological parents. Participants with NAFE and GGE were required to have a first degree relative who also had NAFE or GGE (did not have to be concordant). All patients had no confirmed genetic or metabolic diagnosis, and no history of congenital TORCH infection, premature birth (before 32 weeks gestation), neonatal hypoxic-ischaemic encephalopathy or neonatal seizures, meningitis/encephalitis, stroke, intracranial haemorrhage, significant head trauma, or evidence of acquired epilepsy. Enrolment required detailed confirmation of detailed phenotypic data including medical record review and abstraction, patient interviews, EEG and MRI, and comprehensive review by expert scientific cores for EEG, MRI, and clinical final diagnosis.

Relevant publications:
EPGP Collaborative. The epilepsy phenome/genome project. Clin Trials. 2013;10(4):568-86. doi: 10.1177/1740774513484392. PMID: 23818435.

Funding/acknowledgements:

EPGP was supported by National Institute of Neurological Diseases and Stroke (NINDS) grant U01 NS053998, as well as planning grants from the Finding a Cure for Epilepsy and Seizures (FACES) Foundation and the Richard Thalheimer Philanthropic Fund.

**USA: HEP (USAHEP)**

Participants were recruited for the Human Epilepsy Project at 33 different medical centers located in the US , Canada, Australia, Austria, Finland, and Ireland. All participants were between 12 and 60 at the age of enrolment, and had a clinical history consistent with a diagnosis of focal epilepsy, as determined by an eligibility panel of epilepsy specialists. Participants were required to have two or more spontaneous seizures with clinically observable features in the past 12 months, and 4 or fewer months of anticonvulsant treatment. Those with major medical comorbidities, intellectual disability, or significant psychiatric disease were excluded, as were those with progressive neurological lesions on imaging or known neurodegenerative disease. Participants completed daily electronic diaries tracking seizures, medication adherence, and mood. Mood and cognition were assessed periodically via standardized instruments, and brain MRIs and EEGs were obtained for all participants. Blood was collected and banked annually, allowing for study of DNA, RNA and protein. HEP was approved by the IRBs at all participating sites, and all participants or their parent/legal guardian gave written informed consent. Minors also gave written assents.

Funding/acknowledgements:
HEP is sponsored by The Epilepsy Study Consortium (ESCI). ESCI is a non-profit organization dedicated to accelerating the development of new therapies in epilepsy to improve patient care. The funding provided to ESCI to support HEP comes from industry, philanthropy and foundations (UCB Pharma, Eisai, Pfizer, Lundbeck, Sunovion, The Andrews Foundation, The Vogelstein Foundation, Finding A Cure for Epilepsy and Seizures (FACES), Friends of Faces and others).

**USA: Lurie Children's Hospital of Chicago (USALCH)**

Recruitment site/Institution: Ann & Robert H. Lurie Children's Hospital of Chicago
Study Population: Children with rare intractable epilepsy
Study: Patient samples banked in Lurie's Epilepsy Biobank.
Diagnostic instruments: --
DNA Source: Extracted DNA from whole blood

**USA: Massachusetts General Hospital (USAMGH)**

Adult patients (18 years and older) with genetic generalized epilepsy were recruited from the MGH Epilepsy clinic based on physician referral and/or chart review. Samples were obtained from the Mass General Brigham Biobank, a biorepository of consented patients samples at Mass General Brigham (parent organization of Massachusetts General Hospital and Brigham and Women's Hospital). Phenotypic information was reviewed prior to specimen submission to confirm eligibility for Epi25, and available specimens from the Biobank were submitted to Epi25.

**USA: Mount Sinai (USAMSS)**

Participants were recruited to the neurodevelopmental disorders genetic research program at Icahn School of Medicine at Mount Sinai (ISMMS) via epilepsy clinics in New York City and referrals from neurologists. Study protocols were approved by the ISMMS Institutional Review Board (IRB-12-0079 PI: Pinto) and supported by a NIH/NIMH grant (R01MH110555, PI: Pinto). Written informed consent was obtained from study participants or their legal guardians. Participants had clinical, neuroimaging and EEG/ video-EEG characteristics meeting the International League against Epilepsy (ILAE) 2017 Seizure Classification. A team of researchers and clinicians conducted clinical phenotyping that involved a review of individual medical records, including EEG and MRI reports. Seizure types, age of onset, EEG features and neuroimaging, neurological examinations, neuropsychological assessments, family history, and response to treatment were evaluated. Information on family history of seizures and other neurological disorders was also collected via interviews with legal guardians, participants, and their families. Genomic DNA was extracted from peripheral blood (93.4%) or Oragene/saliva (DNA Genotek) (6.6%) samples with Gentra Puregene Kits (Qiagen) according to the manufacturer's guidelines.

**USA: Nationwide Children's Hospital (USANCH)**

Patients were recruited from a number of institutions including Mt. Sinai Medical Center, New York, Columbia University, Beth Israel, Montefiore Hospital, Einstein, Long Island Jewish (all in New York); Beth Israel-Deaconess, Brigham and Women's (Boston); and a few others in various places.

Families were ascertained. Families were included if: 1) Proband had idiopathic generalized epilepsy without complications (e.g. psychiatric symptoms, birth trauma); generalized interictal EEG bursts; no evidence of focal symptoms (e.g. aura); no history of head injuries; onset age of seizures btw 8-25. The availability of at least one sibling for inclusion or sufficient family members to have the family be informative.

Relevant publications:

Greenberg DA, Durner M, Keddache M, et al. Reproducibility and complications in gene searches: Linkage, heterogeneity, association, and inheritance in juvenile myoclonic epilepsy. AJHG. 2000;66:508-516.

Durner M, Keddache M, Shinnar SS, et al. Genome scan of idiopathic generalized epilepsy: Evidence for major susceptibility gene and modifying genes influencing seizure type. Annals of Neurology. 2001;49:328-335.

**USA: Penn/CHOP (USAUPN)**

The Penn/CHOP cohort included adult and pediatric patients with epilepsy who were seen in our inpatient or outpatient clinical epilepsy programs and enrolled in our ongoing epilepsy research protocols. Biospecimens stored in our institutional biobanks at the time of enrolment were contributed to Epi25. Phenotypic information was reviewed prior to specimen submission to confirm eligibility for Epi25.

**USA: Vanderbilt University Medical Centre (USAVAN)**

Epilepsy cases and controls were identified through extraction of structured data from the electronic health record at Vanderbilt University Medical Center. Samples were collected from existing bio-banked samples. This work was approved by the VUMC IRB#181185. We identified cases corresponding to each epilepsy subtype, limited to subjects with DNA available for external assays. Subtypes included 1) generalized genetic epilepsy (GGE) cases based on the presence of ICD-9 codes 345.0*, 345.1*, 345.2, 345.3, or ICD-10 codes G40.3*, G40.4*, G40.B* (and excluding subjects with ICD codes for focal epilepsy, as described below), 2) non-acquired focal epilepsy (NAFE) cases based on the presence of ICD-9 codes 345.4* and 345.5*, and ICD-10 codes G40.0*, G40.1*, G40.2*, G40.3* (and excluding subjects with ICD codes for generalized epilepsy, as described above), 3) lesional focal epilepsy, based on the presence of ICD codes for focal epilepsy, as described above, plus the presence of ICD codes/groups related to traumatic brain injury (ICD9: 850*, 851*, 852*, 853*, 854*; ICD10: S06*), stroke (ICD9: 430* - 435*; ICD10: I63*, I65*, I66*), or benign tumor (ICD9: 225.0, 225.1, 225.2, 239.6; ICD10: D33.0, D33.1, D33.2, D43.0, D43.1, D43.2, D43.3), that are excluded from all other sets (except the keyword-only set described below), and 4) epileptic encephalopathy (EE) based on the presence of ICD codes for generalized or focal epilepsy cases, as described above, as well as the presence of ICD codes related to encephalopathies (ICD-9: 348.3,

348.30, 348.31, 348.39; ICD-10: G93.4*), that are excluded from all other sets (except the keyword-only set described below).

Additionally, we conducted a search allowing the presence of ICD codes for either generalized or non-acquired focal epilepsy (as described in 1 and 2, above), and excluding subjects already identified in at least one of those sets. This allowed us to include additional subjects who may have received ICD codes for both subtypes, but whose records, upon careful review, clearly support one type and not the other. Reviewers included a team of board certified epileptologists, one medical student, one graduate student, and one research associate. All reviewers were trained by the clinical domain experts who also adjudicated any ambiguous charts. Based on clinician recommendation, we also performed a keyword search for the presence of "idiopathic generalized epilepsy", "genetic generalized epilepsy", or "primary generalized epilepsy" in clinic notes, excluding subjects already captured by other algorithms.
During chart review of this set, we required the presence of one of these phrases in a diagnostic context within an EEG note. For all datasets (except the keyword-only set) we also required the presence of epilepsy related medications and CPT codes indicative of diagnostic neuroimaging procedures (used in chart review), while excluding ICD codes related to various confounding conditions (e.g., cerebral infarctions) except as noted above. All datasets (with the exception of the keyword-only set) required the presence of at least 4 ICD codes from ICD-9 group 345* or ICD-10 group G40*. All samples selected based on ICD codes were subject to chart review for diagnostic confirmation.

Controls were selected based on the absence of the above case inclusion and exclusion criteria.

**Details of individual control cohorts**

**Italian controls (PI: Spalletta)**

Right-handed healthy individuals were recruited and had whole blood drawn by local advertisements at Santa Lucia Foundation in Rome, Italy. All of the individuals were born and educated in Italy and had Italian-Caucasian ancestry, to reduce the possibility of artifactual association caused by ethnic stratification. Exclusion criteria were: (i) major medical illnesses and/or known or suspected history of alcoholism or drug dependency and abuse; (ii) mental disorders (i.e. schizophrenia, mood, anxiety, personality and/or any other significant mental disorders) according to the DSM-IV-TR criteria assessed by the Structured Clinical Interviews for DSM-IV-TR [SCID-I and SCID-II] and/or neurological disorders diagnosed by an accurate clinical neurological examination; (iii) presence of vascular brain lesions, brain tumor and/or marked cortical and/or subcortical atrophy on magnetic resonance imaging (MRI) scan; and (iv) suspicion of cognitive impairment or dementia based on Mini Mental State Examination (MMSE) scores ≤24 (a cut-off point for dementia screening in the Italian population)[1] and confirmed by a clinical neuropsychological evaluation using the Mental Deterioration Battery[2] and the NINCDS-ADRDA criteria for dementia.[3] The presence of anxiety symptoms was assessed using the Hamilton Rating Scale for Anxiety (HAM-A) (Hamilton). Written, informed consent was obtained from all subjects participating in the study, which was approved by the local ethics committee at the Santa Lucia Foundation of Rome (protocol number CE/11.9).[4] Whole exome sequencing was done by the Broad Genomics Platform and data is available by application to dbGaP/AnVIL under accession number phs0001489.

**UK/Ireland controls 1 (PI: McQuillan)**

The UCL control sample consisted of 480 genomic DNA samples that were extracted from EBV transformed peripheral blood lymphocytes from unscreened healthy British blood donors (https://www.phe-culturecollections.org.uk/products/dna/hrcdna/hrcdna). The remaining DNA samples were extracted from whole blood samples from healthy volunteers of UK or Irish ancestry who were interviewed with the initial clinical screening questions of the SADS-L and selected on the basis of not having a past or present personal history of any RDC-defined mental disorder. Heavy drinking and a family history of schizophrenia, alcohol dependence or bipolar disorder, were also used as exclusion criteria for controls. UK National Health Service multi-centre and local research ethics approvals were obtained and all subjects gave signed informed consent.[5] Whole exome sequencing was done by the Broad Genomics Platform and data is available by application to the European Genome Phenome Archive (EGA) under accession number EGAS00001005851.

**UK/IRL controls 2 (PIs: McIntosh, Blackwood, Johnstone)**

Participants were recruited from clinical service around Edinburgh and Scotland and screened using the SADS-L. DNA samples were extracted from whole blood for genotyping and sequencing. Research was conducted after research ethics and NHS management approvals (Schizophrenia Working Group of the Psychiatric Genomics, C). Whole exome sequencing was done by the Broad Genomics Platform and data is available by application to the European Genome Phenome Archive (EGA) under accession number EGAS00001005843.

**German controls (PI: Reif)**

Subjects have been recruited at the Department of Psychiatry and Psychotherapy, University of Würzburg, Germany, with the exception of the TK samples (n=63, they are anonymous blood donors). All subjects have been screened for the absence of mental disorders (by MINI) as well as severe medical and neurological (including epilepsy) disorders (by self-report). Ethnicity is Caucasian by self-report in all cases, and DNA source is blood. Studies were approved by the IRB, University Hospital Würzburg; all participants gave written informed consent. Whole exome sequencing was done by the Broad Genomics Platform and data is available by application to the European Genome Phenome Archive (EGA) under accession number EGAS00001005858.

**FINRISK controls (PI: Palotie)**

The controls from FINRISK that contributed to the Epi25 WES study were part of the FINRISK inflammatory bowel disease (IBD) cohort. The population-based FINRISK study has been followed up for IBD and other disease end-points using annual record linkage with the Finnish National Hospital Discharge Register, the National Causes-of-Death Register and the National Drug Reimbursement Register. Controls were chosen to have a high polygenic risk score for IBD without an IBD diagnosis. A detailed description of the FINRISK cohort can be found at [6]. Whole exome sequencing was done by the Broad Genomics Platform and data is available via application to the THL Biobank: https://thl-biobank.elixir-finland.org/.

**Genomic Psychiatry Cohort (GPC) controls (PIs: Pato M, Pato C, McCarroll)**

The controls from GPC that contributed to the Epi25 WES study were a subset of the overall control participants of European or Latino ancestry with no personal or family history of schizophrenia or bipolar disorder. All the samples were exome-sequenced at the Broad Institute. A detailed description of the GPC cohort can be found at [7]. Whole exome sequencing was done by the Broad Genomics Platform and data will be made available in dbGaP/AnVIL under accession number phs002041.

**ALSPAC**

ALSPAC is a large geographically homogeneous prospective birth cohort from the southwest of England established to investigate environmental and genetic characteristics that influence health, development and growth of children and their parents.[8-11] ALSPAC is now a three-generational study, comprising 'G0': the cohort of original pregnant women, the biological father and other carers/partners; 'G1': the cohort of index children and 'G2': the cohort of offspring of the index children.Full details of the cohort and study design have been described previously and are available at http://www.alspac.bris.ac.uk. Please note that the study website contains details of all the data that are available through a fully searchable data dictionary and variable search tool (http://www.bristol.ac.uk/alspac/researchers/our-data/). Samples were selected for whole exome sequencing at the Broad Institute from the G1 cohort (the cohort of index children) and were from subjects who were singletons/unrelated and of European/British ancestry, had blood-derived DNA available, and had been genotyped on a whole genome genotyping array. Subjects were also chosen based on the availability of phenotype data. Ethical approval for the study was obtained from the ALSPAC Ethics and Law Committee and the Local Research Ethics Committees. Consent for biological samples was collected in accordance with the Human Tissue Act (2004) and informed consent for the use of data collected via questionnaires and clinics was obtained from participants following recommendations of the ALSPAC Ethics and Law Committee at the time. Written informed consent was obtained from mothers at recruitment, from the main carers (usually the mothers) for assessments on the children from ages 7 to 16 years and, from age 16 years onwards, the children gave written informed consent at all assessments. Data is available via an application through the ALSPAC portal: http://www.bristol.ac.uk/alspac/researchers/access/.

**Hong Kong Osteoporosis Study controls (PIs Cheung, Sham, Li)**
The control samples were part of the follow-up study from the Hong Kong Osteoporosis Study (HKOS), which was described elsewhere.[12] Briefly, community-dwelling Southern Chinese were firstly recruited from public roadshows in Hong Kong from 1995 to 2010. An extensive in-person follow-up study was initiated in 2015. At the in-person follow-up visit, the study participants were required to complete a comprehensive self-reported questionnaire, comprising questions related to their medical history, which were checked by experienced researchers or nurses based on a standard protocol. Fasting blood samples were collected from the study participants and DNA was extracted from the sera samples. Study participants without any history of epilepsy at the in-person follow-up in 2019 were included as controls of the epilepsy project. The study protocol was approved by the Institutional Review Board of the University of Hong Kong and the Hospital Authority Hong Kong West Cluster (Ref: UW 15-236). All HKOS participants provided informed consent for participation in the study. Whole exome sequencing was done by the Broad Genomics Platform and data is available at dbGaP/AnVIL under accession number phs0001489.

**USA controls 1 (PIs: Dickerson, Yolken)**
Control samples were collected as part of a larger study about infectious agents and immune factors in serious mental illness. Psychiatric participants were recruited at a large psychiatric health system and non-psychiatric controls from the same geographic region. The diagnosis of non-psychiatric participants was confirmed with a structured clinical interview[13,14] based on DSM-IV (American Psychiatric Association). All participants provided written informed consent. The study was approved by the IRB of the institution where the study was performed and included a data sharing agreement. Whole exome sequencing was done by the Broad Genomics Platform. Data is hosted on the Terra platform (http://app.terra.bio).

**USA controls 2 (PI: Smoller)**
Samples were collected as part of the International Cohort Collection for Bipolar Disorder (ICCBD). [15,16] The Massachusetts General Hospital site of the ICCBD collected DNA from cases (patients with bipolar disorder) and controls by linking discarded blood samples to de-identified electronic health record (EHR) data. Cases and controls were identified by deriving EHR-based phenotyping algorithms applied to the Partners Healthcare Research Patient Data Registry (RPDR), described in detail previously.[17] Full details of the algorithm are provided in [16]. Samples were whole exome sequenced by the Broad Genomics Platform

and data from control samples were included in Epi25 analyses. Data is hosted on the Terra platform (http://app.terra.bio).

**Dutch controls (PI: Postuma)**

Controls taken from the NESCOG study, described previously.[18] NESCOG contains both a general population and family-based sample of which closely related individuals were excluded. Data were collected on cognitive tasks, behavioral conditions, life events, personality and environmental factors. To correct for undiagnosed attention deficit hyperactivity disorder (ADHD) status, participants scoring over three standard deviations above the mean on the Conners' Adult ADHD Rating Scale (CAARS)[19], or the Attention Problems scale of the Young Adult Self Report (YASR)[20] were excluded. To correct for autism spectrum disorder (ASD) status, participants scoring over three standard deviations above the mean on the Autism Quotient (AQ)[21] were removed. Whole exome sequencing was done by the Broad Genomics Platform and data is available by application to the European Genome Phenome Archive (EGA) under accession number EGAS00001005857.

**NIDDK Inflammatory Bowel Diseases Genetics Consortium (NIDDK IBDGC)**

The NIDDK Inflammatory Bowel Disease Genetics Consortium (IBDGC) was created in 2002 by the National Institute of Diabetes, Digestive and Kidney diseases (NIDDK) to advance knowledge on the inflammatory bowel diseases, specifically Crohn's Disease and Ulcerative Colitis. The Consortium consists of six genetic research centers (GRC) and a data coordinating center (DCC) that prospectively recruits a combination of cases, controls, and trios to gather a large collection of samples and linked phenotype information. DNA samples are used to conduct genetic linkage and association studies. For more information please see https://ibdgc.org/. Control samples from the following cohorts were included in the Epi25 analysis: The University of Pittsburgh School of Medicine (PI: Richard Duerr), The Johns Hopkins Hospital (PI: Steven Brant), The Icahn School of Medicine at Mount Sinai (PI: Judy Cho), and Cedars Sinai (PI: Dermot McGovern, Stephan Targan). All samples were whole exome sequenced by the Broad Genomics Platform and data is available by application to dbGaP/AnVIL under accession number phs0001642.

**Mass General Brigham (MGB) Biobank**

The MGB (formerly Partners) Biobank (https://biobank.massgeneralbrigham.org/), launched in 2010, is a biorepository of consented patients samples at Mass General Brigham (parent organization of Massachusetts General Hospital and Brigham and Women's Hospital). The Biobank has enrolled >100K individuals to study how genes, lifestyle, and other factors affect people's health and contribute to disease. As part of the NHGRI's Centers for Common Disease Genomics, Broad Institute of MIT and Harvard generated genetic data for ~13,500 individuals from the MGB Biobank. Data is available by application to dbGaP/AnVIL under accession number phs0002018.

**Supplementary Acknowledgments**

**Controls locally available at the Broad Institute**

Collection of the Italian controls was supported by RF-2013-02359074; NET-2011-02346784 (Italian Ministry of Health) and samples were sequenced using CCDG funding. UK/Ireland controls 1 were collected with support from the Neuroscience Research Charitable Trust, the Central London NHS (National Health Service) Blood Transfusion Service and the National Institute of Health Research (NIHR) funded Mental Health Research Network. The GPC controls were sequenced with funding from NIH grants MH085548 and MH085542 and the Stanley Center for Psychiatric Research, Broad Institute of MIT and Harvard. We thank the Genomic Psychiatry Cohort (GPC) Consortium teams including: Evelyn J. Bromet, Celia Barreto Carvalho, Eric D. Achtyes, Maria Helena Azevedo, Roman Kotov, Douglas S. Lehrer, Dolores Malaspina, Stephen R. Marder, Helena Medeiros, Christopher P. Morley, Diana O. Perkins, Janet L Sobell, Peter F. Buckley, Fabio Macciardi, Mark H. Rapaport, James A. Knowles, and Ayman H. Fanous. Sample collection of the German controls was supported by the German Research Foundation (DFG TRR-CRC 58, Z02) and whole exome sequencing was funded by the Dalio Foundation. The FINRISK controls were part of the FINRISK studies supported by THL (formerly KTL: National Public Health Institute) through budgetary funds from the government, with additional funding from institutions such as the Academy of Finland, the European Union, ministries and national and international foundations and societies to support specific research purposes. The UK Medical Research Council and Wellcome (Grant ref: 217065/Z/19/Z) and the University of Bristol provide core support for ALSPAC. This publication is the work of the authors who will serve as guarantors for the contents of this paper. We are extremely grateful to all the families who took part in this study, the midwives for their help in recruiting them, and the whole ALSPAC team, which includes interviewers, computer and laboratory technicians, clerical workers, research scientists, volunteers, managers, receptionists and nurses. The ALSPAC samples were sequenced with funding from the Stanley Center for Psychiatric Research, Broad Institute of MIT and Harvard. NJT is a Wellcome Trust Investigator (202802/Z/16/Z), is the PI of the Avon Longitudinal Study of Parents and Children (MRC & WT 217065/Z/19/Z), is supported by the University of Bristol NIHR Biomedical Research Centre (BRC-1215-2001), the MRC Integrative Epidemiology Unit (MC_UU_00011/1) and works within the CRUK Integrative Cancer Epidemiology Programme (C18281/A29019). The collection of the Hong Kong Osteoporosis study samples was funded by the Bone Health Fund and Research Grants Council - Early Career Scheme (Project number: 27100416). Whole exome sequencing was done by the Broad Genomics Platform and supported by the NHGRI CCDG grant (UM1HG008895). USA Controls 1: Sequencing of the USA controls was funded by the Dalio Foundation. USA Controls 2: ICCBD sample and data collection was supported by R01 MH085542. Sequencing of the USA controls was funded by the Dalio Foundation. Dutch controls: Sequencing of the Dutch controls was funded by the Dalio Foundation. NIDDK IBDGC: We thank the National Institute of Diabetes and Digestive and Kidney Diseases (NIDDK) IBD Genetics Consortium (IBDGC) supported by The Helmsley Charitable Trust and the Centers for Common Disease Genomics (NHGRI CCDG). Whole exome sequencing was done by the Broad Genomics Platform and supported by the NHGRI CCDG grant (UM1HG008895). MGB Biobank: We gratefully acknowledge the participants and leadership team of the MGB Biobank, funding support from the NHGRI CCDG (UM1HG008895), and generation of new whole exome sequencing data by the Broad Genomics Platform.

**Cohorts accessed via dbGaP**

Controls from the MIGen Leicester study were obtained through dbGaP accession number phs001000.v1.p1. We thank the Broad Institute for generating high-quality sequence data for the MIGen studies supported by NHGRI funds (grant # U54 HG003067) with Eric Lander as PI. Data from Epi4k were obtained through dbGaP accession numbers phs000654.v3.p1, phs001558.v1.p1, and phs001551.v1.p1. For phs000654.v3.p1 and phs001558.v1.p1, we acknowledge the Epi4K Gene Discovery in Epilepsy study

(NINDS U01-NS077303) and the Epilepsy Genome/Phenome Project (EPGP - NINDS U01-NS053998) For phs001551.v1.p1, we acknowledge the Epilepsy Genetics Initiative, A Signature Program of CURE.
